# MultiAge: A New Multidimensional Biomarker of Biological Age Derived from Comprehensive Phenotypic and Molecular Profiling

**DOI:** 10.64898/2026.06.01.26354629

**Authors:** Valentin Max Vetter, Marit Philine Junge, Ganling Ding, Annemarie Luise Weihs, Johanna Drewelies, Sandra Düzel, Jan Homann, Eva-Marie Maetzel, Dominik Spira, Hans Jörgen Grabe, Eva Grill, Ulman Lindenberger, Matthias Nauck, Graham Pawelec, Anette Peters, Elisabeth Steinhagen-Thiessen, Barbara Thorand, Henry Völzke, Juliane Winkelmann, Klaus Berger, Alexander Teumer, Melanie Waldenberger, Denis Gerstorf, Christina M. Lill, Lars Bertram, Ilja Demuth

## Abstract

**Background:** It is an everyday observation that people of the same chronological age differ with respect to their physical and mental capacity. However, assessing these differences in biological age remains challenging.

**Methods:** Here, we aggregate 89 age-associated variables from the Berlin Aging Study II (BASE-II, n=1,631) to generate MultiAge, a new marker of biological age that summarizes information from ten domains reflecting organ health and global biological age. We then used methylation data obtained from an Illumina MethylationEPIC array and supervised machine learning to translate MultiAge into a DNA methylation signature, MultiAgeEpi (309 CpGs), which was subsequently validated in four independent external validation cohorts (KORA FF4, KORA Age, SHIP-TREND, BiDirect, total n=4,339). MultiAgeEpi results were compared with previously published epigenetic clocks (GrimAge, DunedinPACE, SystemsAge).

**Results:** We report that MultiAgeEpi showed similar, and in several cases, stronger associations with age-associated outcomes such as diabetes, metabolic syndrome, multimorbidity, frailty and mortality (q < 0.05) compared to the other clocks.

**Conclusions:** MultiAge and MultiAgeEpi thus provide a comprehensive assessment of biological age through aggregation of numerous age-associated variables and the use of the high-resolution methylomics data makes transfer of this marker to other cohorts possible.

## Introduction

The effective assessment of biological age has the potential to substantially change how diseases are investigated, prevented and treated (1–4). Generally, phenotypes of aging become clinically measurable after resilience mechanisms, which lose efficacy over the life course, are no longer able to maintain the homeostatic state and fail to adequately respond to stressors (5). Assessing biological age before associated phenotypes manifest clinically allows earlier monitoring of process and enables early and targeted interventions to prevent down-stream manifestations of disease or other adverse age-associated phenotypes. On the other hand, a sensitive monitoring of biological age offers the possibility of trialing interventions aimed at promoting a healthy aging process more effectively (4). While participants would need to be followed for an unfeasibly extended period to assess the impact of these interventions in real-time, measuring a marker of biological age would allow detailed investigations of potential intervention effects long before they become clinically measurable.

However, this approach requires a best-as-possible estimation of the “true” biological age. To achieve this goal, a number of aging biomarkers have been developed and intensively evaluated (4, 6, 7) making use of numerous data types including proteomics, laboratory measurements, telomere length, magnetic resonance imaging, and others. One of the most promising types of biomarkers of aging, epigenetic clocks, are derived from DNA methylation data (6, 8). While the first generation of epigenetic clocks was trained to predict chronological age (9, 10), subsequent iterations used more sophisticated outcomes to train the methylation signatures. However, while the associations with health-associated outcome variables of these second (11, 12) and third generation epigenetic clocks (13) became stronger, they still seem to perform especially well only within certain domains of aging which were often closely related to the manner by which the methylation signatures were constructed (14–16). While these outcome variables often rely on only a few variables, partly due to limited data availability in the cohorts used to develop these epigenetic clocks, other recent approaches make use of a wider variety of variables from different organ systems (17). For example, the SystemsAge clock (17) focuses on the development of methylation signatures for specific organ systems by performing principal component analyses (PCA) on the variables representing the health of each system, using data from the Health and Retirement Study (HRS). This allows a more granular assessment of the individual’s biological age due to a strong focus on the individual organ systems instead of the overall aging process. In a second step, SystemsAge was derived by weighting each epigenetically assessed organ system score in a mortality prediction model based on data from the Framingham Heart Study (FHS). In most cases, SystemsAge as well as the organ system scores showed either similar or stronger associations with the investigated outcome variables in three large external validation cohorts compared to other epigenetic clocks (17). As expected, the system scores are associated more strongly with outcomes closely linked to the variables they were trained on, and they often outperformed SystemsAge score. While these results are highly promising, this strong focus on individual organ systems might come at the expense of a more holistic biological age assessment.

In this study, we aimed to distill a measure of biological age from 89 age-associated variables assessed in >1,600 participants of the Berlin Aging Study II (BASE-II) into one comprehensive measure of overall biological age, the MultiAge, using and comparing five different statistical methods (18–21). By constructing MultiAge in sex-stratified groups and aggregating information without lifespan-weighted variables, our marker captures sex-associated aging differences (2, 22, 23) and prevalent health status to address the healthspan-lifespan gap (3, 24–27), offering a novel tool for studies aiming at minimizing this very feature. To make the MultiAge marker transferable to other cohorts, we derived a DNA methylation signature, MultiAgeEpi, which consists of 309 CpGs covered by both Illumina’s EPIC arrays v1 and v2. MultiAgeEpi was then validated in four independent, external test data sets from Germany (total n=4,339). Our results show that MultiAgeEpi provides a new DNA methylation aging marker that outperforms existing epigenetic clocks for a number of aging-relevant outcomes.

## Methods

### Development of MultiAge and MultiAgeEpi

#### BASE-II study population

MultiAge was developed using data from 1,631 participants (mean age, 68.7; 51.2% women, Table 1) of the baseline assessment of the Berlin Aging Study II (BASE-II) (28). For a flowchart of BASE-II participants, please see Supplementary Figure 4. BASE-II is a longitudinal, observational multi-center study aiming at the identification of factors that promote healthy aging. On average 7.4 years after the baseline assessment, 1,083 participants were followed-up as part of the GendAge study (29). A detailed description of how the investigated variables were measured and processed as well as how missing values were imputed is described in the Supplementary Material. This manuscript was created in accordance with the STROBE guidelines (4).

**Table 1:**
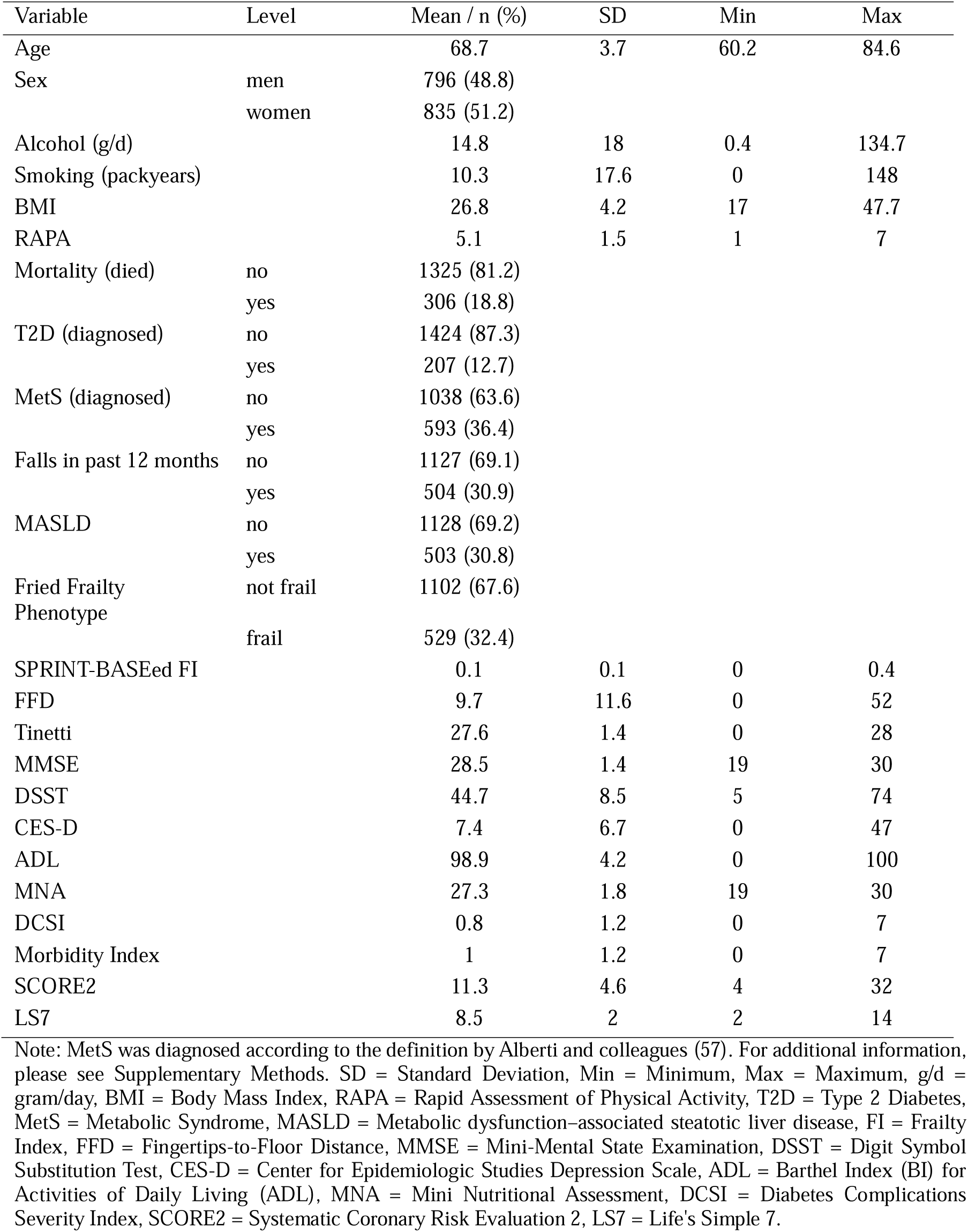
Descriptive statistics of selected BASE-II baseline data used to construct MultiAge and MultiAgeEpi (n=1,631).

| Variable | Level | Mean / n (%) | SD | Min | Max |
| --- | --- | --- | --- | --- | --- |
| Age |  | 68.7 | 3.7 | 60.2 | 84.6 |
| Sex | men | 796 (48.8) |  |  |  |
|  | women | 835 (51.2) |  |  |  |
| Alcohol (g/d) |  | 14.8 | 18 | 0.4 | 134.7 |
| Smoking (packyears) |  | 10.3 | 17.6 | 0 | 148 |
| BMI |  | 26.8 | 4.2 | 17 | 47.7 |
| RAPA |  | 5.1 | 1.5 | 1 | 7 |
| Mortality (died) | no | 1325 (81.2) |  |  |  |
|  | yes | 306 (18.8) |  |  |  |
| T2D (diagnosed) | no | 1424 (87.3) |  |  |  |
|  | yes | 207 (12.7) |  |  |  |
| MetS (diagnosed) | no | 1038 (63.6) |  |  |  |
|  | yes | 593 (36.4) |  |  |  |
| Falls in past 12 months | no | 1127 (69.1) |  |  |  |
|  | yes | 504 (30.9) |  |  |  |
| MASLD | no | 1128 (69.2) |  |  |  |
|  | yes | 503 (30.8) |  |  |  |
| Fried Frailty Phenotype | not frail | 1102 (67.6) |  |  |  |
|  | frail | 529 (32.4) |  |  |  |
| SPRINT-BASEd FI |  | 0.1 | 0.1 | 0 | 0.4 |
| FFD |  | 9.7 | 11.6 | 0 | 52 |
| Tinetti |  | 27.6 | 1.4 | 0 | 28 |
| MMSE |  | 28.5 | 1.4 | 19 | 30 |
| DSST |  | 44.7 | 8.5 | 5 | 74 |
| CES-D |  | 7.4 | 6.7 | 0 | 47 |
| ADL |  | 98.9 | 4.2 | 0 | 100 |
| MNA |  | 27.3 | 1.8 | 19 | 30 |
| DCSI |  | 0.8 | 1.2 | 0 | 7 |
| Morbidity Index |  | 1 | 1.2 | 0 | 7 |
| SCORE2 |  | 11.3 | 4.6 | 4 | 32 |
| LS7 |  | 8.5 | 2 | 2 | 14 |

Note: MetS was diagnosed according to the definition by Alberti and colleagues (57). For additional information, please see Supplementary Methods. SD = Standard Deviation, Min = Minimum, Max = Maximum, g/d = gram/day, BMI = Body Mass Index, RAPA = Rapid Assessment of Physical Activity, T2D = Type 2 Diabetes, MetS = Metabolic Syndrome, MASLD = Metabolic dysfunction-associated steatotic liver disease, FI = Frailty Index, FFD = Fingertips-to-Floor Distance, MMSE = Mini-Mental State Examination, DSST = Digit Symbol Substitution Test, CES-D = Center for Epidemiologic Studies Depression Scale, ADL = Barthel Index (BI) for Activities of Daily Living (ADL), MNA = Mini Nutritional Assessment, DCSI = Diabetes Complications Severity Index, SCORE2 = Systematic Coronary Risk Evaluation 2, LS7 = Life's Simple 7.

#### MultiAge development

Based on previous publications by Lopez-Otin et al. (30), Hägg et al. (31), Lara et al. (32), Margolick and Ferrucci (33), and Engelfriet et al. (34) we assembled a list of biomarkers recommended for biological age assessment which we then reconciled with the available variables in BASE-II. Subsequently, we added additional variables showing well-established associations with age which were available in BASE-II. After quality control and exclusion of highly correlated variables (r > |0.8|, to avoid redundancies), 89 variables were available for further processing (Figure 1).

**Figure 1:**
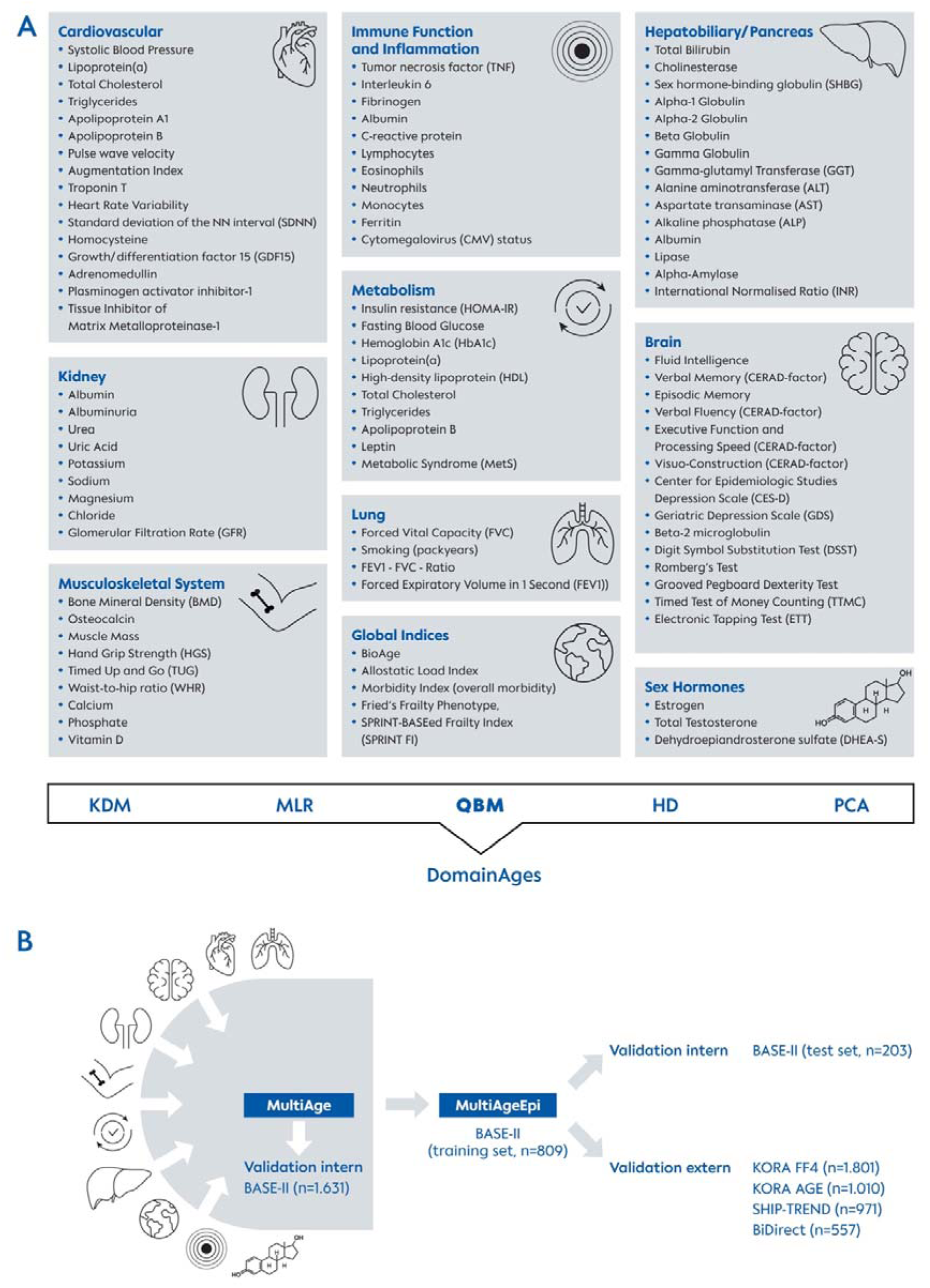
Workflow of the development and construction of DomainAges, MultiAge and MultiAgeEpi using 89 variables from numerous sources and aging domains available in BASE-II (n=1,631). As the first step, a list of biomarkers recommended for biological age assessment was derived from previously published literature. Subsequently, this list was adapted to fit the BASE-II database and extended with variables available in BASE-II that were previously reported to be associated with age in the literature. Variables were then mapped to specific domains reflecting organ health and global biological age and aggregated using five statistical methods established to assess biological age. Comparison of the respective results showed that the quartile-based method (QBM) showed the most strongest consistent association with investigated outcomes (Supplementary Figure 5). Next, the mean across the biological age aggregates in each of the domains was calculated as MultiAge for n=1,631 participants and used to develop a DNA methylation signature in a BASE-II training set (n=809), MultiAgeEpi, which was subsequently internally validated in a BASE-II hold-out test-set (n=203) as well as in four independent external cohorts (KORA FF4, n=1801, KORA Age, n=1010, SHIP-TREND, n=971, BiDirect, n=557). Note: QBM = quartile-based method, KDM = Klemera and Doubal Method, MLR = Multiple Linear Regression, HD = Homeostatic Dysregulation, PCA = Principal Component Analysis.

Development of MultiAge was accomplished in a two-step process. First, individual sex-stratified residuals of a linear regression analysis of continuously scaled variables on chronological age were calculated to determine how each participant differed from the value which would be expected at their age based on the distribution of the sample. By doing so, the resulting value reflects either a “better” (e.g. value >0), “worse” (e.g. value <0) or “equal” (e.g. value ≈ 0) status compared to participants in the same cohort of the same age. Subsequently, all variables that were mapped to one domain (e.g. Brain, Lung, …) were aggregated to respective “DomainAges” (DomainAge (Brain), DomainAge (Lung) etc., Figure 1). To identify the optimal statistical method for variable aggregation, this was done using a quartile-based score method method (QBM), the Klemera and Doubal method (KDM), Multiple Linear Regression (MLR), Homeostatic Dysregulation (HD), and Principal Component Analysis (PCA). Individual description of the specific methods is provided in the Supplementary Methods. Aggregated DomainAges were then compared with respect to their relationship with a panel of 17 age-related outcome variables (almost) all of which were previously used to systematically compare markers of biological age in the BASE-II dataset (35). After adjusting for age, sex, smoking, alcohol consumption, BMI, and physical activity, the quartile-based method showed the strongest associations with 9 of the 17 investigated outcome variables (Supplementary Figure 5). This method, which was previously described by Seeman and colleagues for calculating the Allostatic Load Index (19), awards points to participants based on the distribution of values in the study sample. Participants with values in the highest or lowest quartile indicating impairment or high risk for adverse health events were awarded one point. In the case of binary variables, the outcome reflecting impairment or disease was used to award one point. The average across all variables within one domain was then defined as the respective DomainAge. In a second step, the mean across all DomainAges was calculated to give the final MultiAge value.

#### Development of MultiAgeEpi

DNA methylation data in BASE-II at baseline were available for a subset of n=1,012 participants. An epigenetic signature was trained in an 80% training set (n=809) using supervised machine learning (elastic net regression). In line with the development of previous epigenetic clocks, alpha was set at 0.5 and the best lambda (lambda= 0.0127), a shrinking parameter that controls the strength of regularization (with higher values increasing penalty and reducing model complexity), was identified by 10-fold cross-validation. Quality of model fit was investigated in the remaining 20% hold-out test set of participants which were not part of the training set (Pearsons’r = 0.34, RMSE = 0.09, MAE = 0.07). Detailed information on the pre-processing of BASE-II DNA methylation data and development of MultiAgeEpi is provided in the Supplementary Methods.

### External Validation of MultiAgeEpi

#### KORA FF4

The KORA platform is a long-running research initiative in southern Germany’s Augsburg region (36), featuring multiple baseline surveys: S1 (1984/1985), S2 (1989/1990), S3 (1994/1995), and S4 (1999/2000). As part of the KORA framework, participants undergo different health assessments to observe trends in health conditions, lifestyle factors, and biological indicators at the KORA Study Centre in collaboration with Augsburg University Hospital, Augsburg, Germany. The KORA FF4 study (2013–2014), is the second follow-up of the S4 baseline cohort and participants underwent detailed phenotyping with a focus on cardiometabolic diseases using standardized protocols.

#### KORA Age

The KORA-Age study tracks a group of adults aged 65 to 93 years at baseline in 2008/09, selected from former KORA S1-S4 participants (37–39). The KORA-Age study focused on aged-related conditions such as multimorbidity, frailty, sarcopenia, mental health including cognition and activities of daily living (Ref 38 KORA Age).

#### SHIP-TREND

The SHIP study, based in Western Pomerania (Northeast Germany), investigates the prevalence and incidence of common diseases and their risk factors. For this study, a subsample, SHIP-TREND, which includes 4,420 participants at baseline (2008–2012) and 2,507 at the first follow-up (2016–2019) was used. DNA methylation data was available for 971 participants.

#### BiDirect

The BiDirect Study, conducted in Münster, Germany, investigates the bidirectional relationship between depression and subclinical arteriosclerosis and recruited three distinct cohorts, patients with depression, patients with an acute coronary event and community-dwelling individuals randomly selected in the city registry. All participants were 35 to 65 years at baseline and examined with an identical, complex program four times over 12 years. The present analysis included a subsample of 557 participants drawn from the population-based controls, for which detailed phenotypic and DNA methylation data was available from baseline.

#### Epigenetic clocks assessment

SystemsAge was calculated for all cohorts using the methylCIPHER package as instructed by the authors of the original publication (17). In BASE-II, DunedinPACE was calculated using the package provided by the authors of the original publication (13) and GrimAge was obtained through Steve Horvath’s website (https://horvath.genetics.ucla.edu/html/dnamage/). Detailed information on the assessment of epigenetic clocks in BASE-II was previously published (40, 41). In SHIP-TREND, DunedinPACE (13) and GrimAge were obtained from the biolearn python package (42). In KORA Age and KORA FF4, the methylCIPHER package (17) was used to calculate all clocks used for comparison with MultiAgeEpi in this study. DNA methylation acceleration was calculated for GrimAge and SystemsAge as residuals of a linear regression of epigenetic age on chronological age and leukocyte cell counts.

#### Statistical Analysis

All statistical analyses were conducted using the statistical software package R. Descriptive statistics were obtained through the tableone package. Linear, logistic and Cox regression analyses were conducted to investigate the association with continuously scaled, categorical outcomes and survival. In line with recommendations on the evaluation of biomarkers of aging (43, 44), we provide an unadjusted (model 1), age- and sex-adjusted (model 2), as well as a fully adjusted regression model including age, sex, smoking, alcohol, BMI, and physical activity (model 3). All analyses are conducted in the full dataset as well as in sex-stratified subgroups according to recommendations on the validation of biological age markers (44). This was not done for analyses using the BASE-II hold-out test-set due to its low sample size (n=203). Detailed information regarding the individual steps of the analyses and multiple imputation procedures (if applicable) are detailed in the Supplementary Methods. We emphasize that, by design, MultiAge is independent of chronological age and sex. However, age- and sex-adjusted results are presented for a better comparability with the other investigated epigenetic clocks. To account for multiple testing, p-values were corrected using the Benjamini-Hochberg method for controlling the false discovery rate (FDR) at a threshold of 0.05 and the corresponding q-values are provided. Multiple testing correction was done across all biomarkers, outcomes and across all external validation cohorts as well as for analyses conducted using data from BASE-II. In the Supplementary Tables, original p-values and q-values are reported. An q <0.05 was considered statistically significant.

## Results

### Study population and construction of MultiAge

In this study, we leveraged data from 1,631 participants of the Berlin Aging Study II (BASE-II) between 60 and 85 years of age (mean age, 68.7; 51.2% women, Table 1, Supplementary Tables 1 and 2). Information from 89 age-associated variables matched to ten domains of aging were aggregated to DomainAge using a quartile-based method which awards points to participants within a high risk quartile similar to the procedure described by Seeman and colleagues for calculating the Allostatic Load Index (19). The average across all ten DomainAges was calculated to obtain MultiAge. Because by design, the mean of the whole sample is 0.25 points and differences between individuals are numerically small (Supplementary Figure 1), we standardized DomainAge and MultiAge values prior to statistical analysis to enable an easier interpretation of results.

**Table 2:**
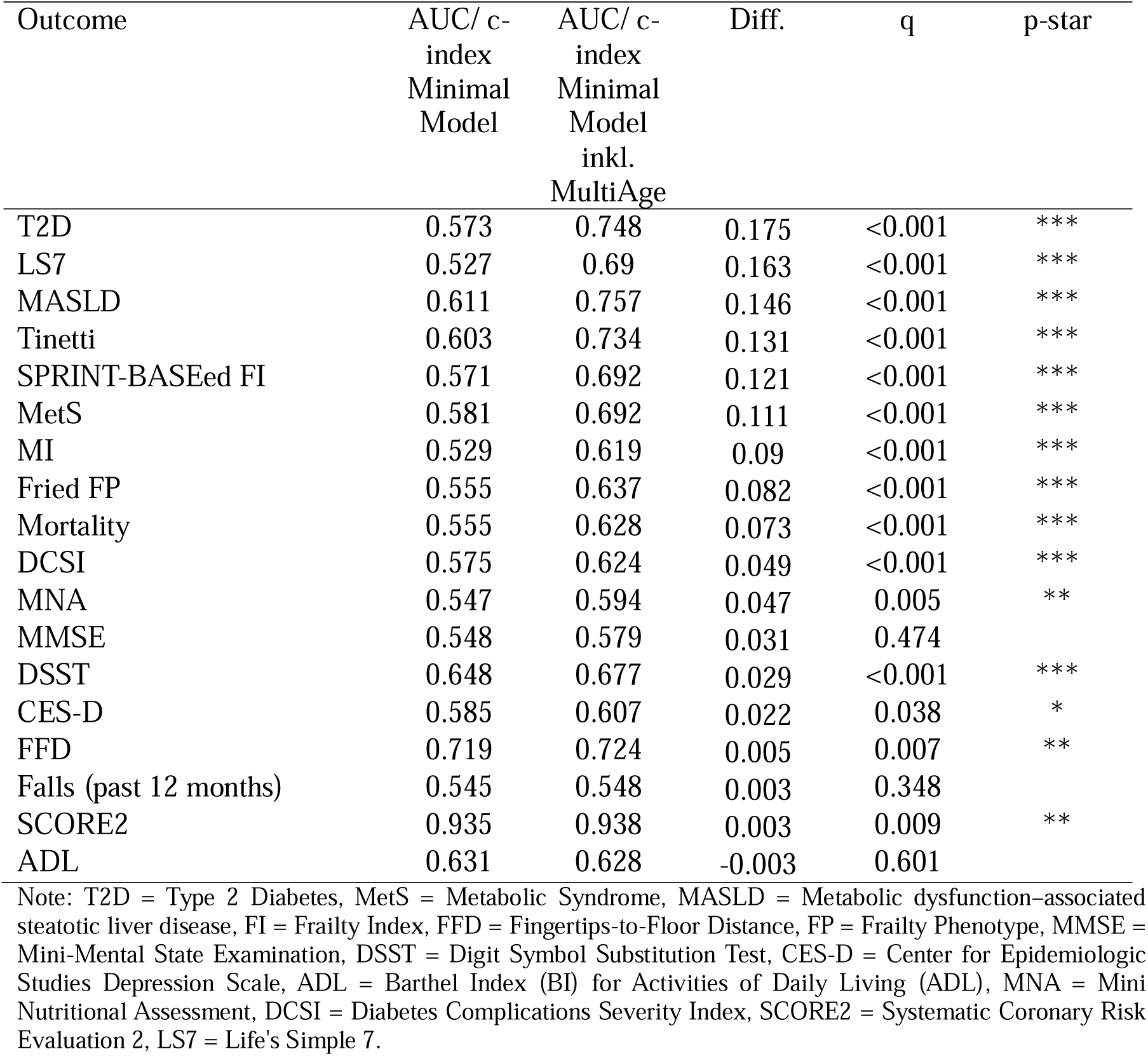
Area Under the Curve (AUC) of a minimal clinical model and a minimal clinical model + MultiAge. Model discrimination was assessed as area under the curve (AUC) from a logistic regression of outcome variables on the minimal clinical model without and with MultiAge. Originally continuously scaled variables were dichotomized based on pre-defined cut-off values. Statistical significance between models was assessed using the log rank test. Comparison of models predicting mortality was done using the c-index of a Cox proportional hazard regression. Statistical significance of models with and without MultiAge was assessed using anova.

| Outcome | AUC/ c-index<br>Minimal<br>Model | AUC/ c-index<br>Minimal<br>Model<br>inkl.<br>MultiAge | Diff. | q | p-star |
| --- | --- | --- | --- | --- | --- |
| T2D | 0.573 | 0.748 | 0.175 | <0.001 | *** |
| LS7 | 0.527 | 0.69 | 0.163 | <0.001 | *** |
| MASLD | 0.611 | 0.757 | 0.146 | <0.001 | *** |
| Tinetti | 0.603 | 0.734 | 0.131 | <0.001 | *** |
| SPRINT-BASEd FI | 0.571 | 0.692 | 0.121 | <0.001 | *** |
| MetS | 0.581 | 0.692 | 0.111 | <0.001 | *** |
| MI | 0.529 | 0.619 | 0.09 | <0.001 | *** |
| Fried FP | 0.555 | 0.637 | 0.082 | <0.001 | *** |
| Mortality | 0.555 | 0.628 | 0.073 | <0.001 | *** |
| DCSI | 0.575 | 0.624 | 0.049 | <0.001 | *** |
| MNA | 0.547 | 0.594 | 0.047 | 0.005 | ** |
| MMSE | 0.548 | 0.579 | 0.031 | 0.474 |  |
| DSST | 0.648 | 0.677 | 0.029 | <0.001 | *** |
| CES-D | 0.585 | 0.607 | 0.022 | 0.038 | * |
| FFD | 0.719 | 0.724 | 0.005 | 0.007 | ** |
| Falls (past 12 months) | 0.545 | 0.548 | 0.003 | 0.348 |  |
| SCORE2 | 0.935 | 0.938 | 0.003 | 0.009 | ** |
| ADL | 0.631 | 0.628 | -0.003 | 0.601 |  |

Note: T2D = Type 2 Diabetes, MetS = Metabolic Syndrome, MASLD = Metabolic dysfunction-associated steatotic liver disease, FI = Frailty Index, FFD = Fingertips-to-Floor Distance, FP = Frailty Phenotype, MMSE = Mini-Mental State Examination, DSST = Digit Symbol Substitution Test, CES-D = Center for Epidemiologic Studies Depression Scale, ADL = Barthel Index (BI) for Activities of Daily Living (ADL), MNA = Mini Nutritional Assessment, DCSI = Diabetes Complications Severity Index, SCORE2 = Systematic Coronary Risk Evaluation 2, LS7 = Life's Simple 7.

### MultiAge in the context of age-associated outcomes

First, we investigated the association of MultiAge with age-associated outcome variables previously used to evaluate biomarkers of aging in BASE-II (15) and mortality. In cross-sectional regression models, MultiAge was statistically significantly associated with all investigated outcomes after adjustment for confounding variables, except falls in the past 12 months (Supplementary Figure 2, Supplementary Table 3 and 4). To account for multiple testing, p-values were adjusted using the Benjamini-Hochberg method (FDR-controlled at 0.05; adjusted p-values are referred to as q throughout the text). In longitudinal analyses investigating the association between MultiAge at baseline and outcome variables assessed on average 7.4 years later, frailty (Fried FP (45, 46), SPRINT-BASEed FI (47, 48)), overall morbditiy (49), nutrition (MNA (50)), cognition (MMSE (51)), cardiovascular health (LS7 (52, 53)), mobility (Tinetti Test (54)), diagnosed type 2 diabetes (T2D), diabetes-associated complications (DCSI (55, 56)) and Metabolic dysfunction–associated steatotic liver disease (MASLD) all showed statistically significant associations (q <0.05) after confounder adjustment (model 3, Figure 2, Supplementary Table 5 and 6). Similarly, in analyses investigating the ability of the marker to predict incident cases at follow-up, MultiAge was statistically significantly associated with incident T2D, impaired mobility, frailty (SPRINT-BASEed FI and Fried FP), cognition (DSST), and all-cause mortality (model 3, Figure 3, Supplementary Table 7 and 8). In line with previous biomarker comparisons in BASE-II (35), the improvement in predicting incident cases of disease or impairment by adding MultiAge to a basic prediction model was calculated. The strongest increase in the AUC for predicting incident cases was found for diagnosed T2D (0.573 vs. 0.748, q <0.001) and LS7 (0.572 vs. 0.690, q <0.001, Table 2). This indicates a possibly substantial benefit of including this marker in models aiming at the prediction of adverse outcomes associated with an unhealthy biological aging process, e.g. in a (pre-)clinical setting.

**Figure 2:**
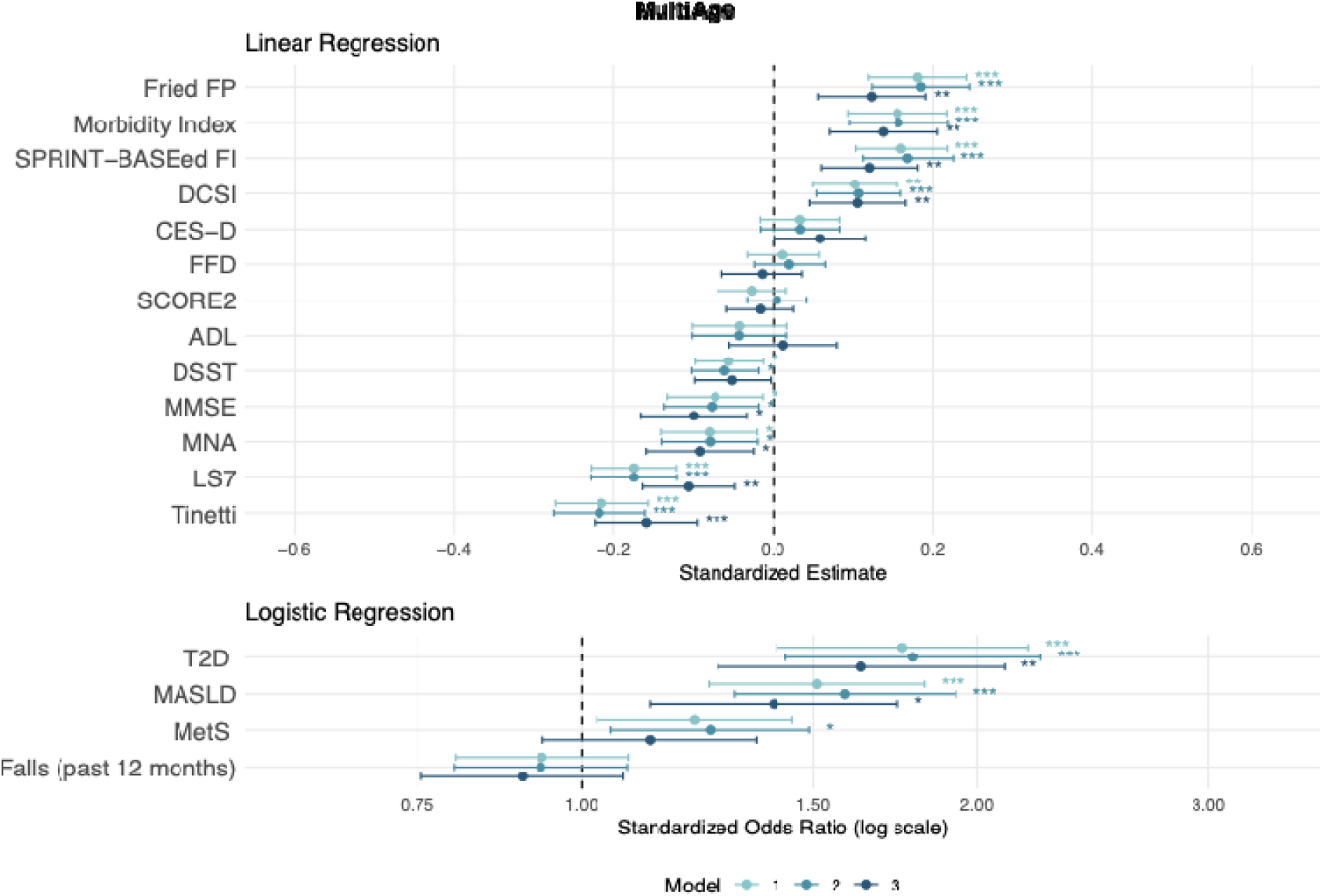
Results from longitudinal regression models of outcomes at T1 on MultiAge at T0 in BASE-II (n=1,072). Results are shown with adjustment for the outcome variable at T0 (model 1), adjusted for the outcome variable at T0, age, and sex (model 2), and adjusted for outcome at T0, age, sex, alcohol intake, smoking, BMI, and physical activity. Forest plots illustrating results from DomainAges are shown in Supplementary Figure 6. Note: * q <0.05, ** q <0.01, *** q <0.001.

**Figure 3:**
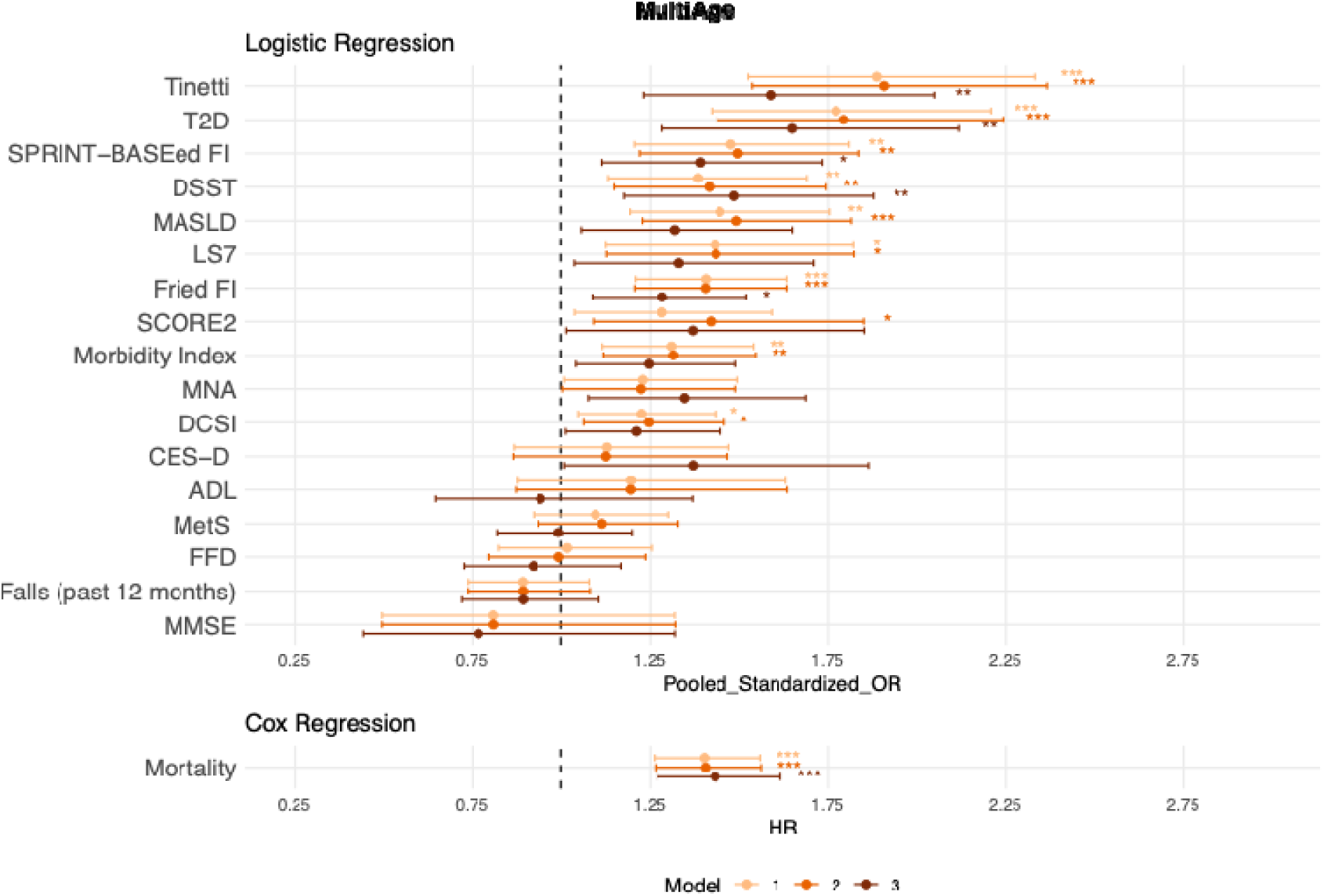
Results from regression models predicting incident cases of disease and impairment over the average 7.4-year follow-up period and mortality in BASE-II. Participants with prevalent disease or impairment in the respective variables were excluded from this analysis (hence the sample size varies between outcome variables). Originally continuously scaled variables were dichotomized based on previously defined cut-off values. Model 1 is unadjusted, Model 2 is adjusted for age and sex, and Model 3 is adjusted for age, sex, alcohol intake, smoking, BMI, and physical activity. Sex-stratified analyses as well as results from DomainAge variables are reported in Supplementary Table 7 and 8. Forest plots illustrating results from DomainAges are shown in Supplementary Figure 7. Note: * q <0.05, ** q <0.01, *** q <0.001.

### Training of MultiAgeEpi in BASE-II

In an effort to make this biomarker transferrable to other datasets, we trained a DNA methylation signature to estimate MultiAge from DNA methylation data. A supervised machine learning approach (elastic net regression) was used to select and weight CpGs from the 850,000 features available in Illumina’s MethylationEPIC array (80% training set, n=809). The methylation signature, MultiAgeEpi, was statistically significantly associated after multiple-testing correction and confounder adjustment with LS7, MI, SPRINT FI, Fried FP, and Metabolic Syndrome (MetS) in a 20% BASE-II hold-out test-set (n=203, Supplementary Table 9). In all cases except SCORE2, stronger associations were found for MultiAge compared to MultiAgeEpi (Supplementary Figure 3). MultiAgeEpi was nominally significantly associated with LS7, SCORE2, MI, MASLD, MetS, and T2D. Even despite the small sample size, the association with MetS remained statistically significant after adjusting for multiple testing (Supplementary Table 10 and 11).

### External Validation of MultiAgeEpi

To assess external validity and transferability of the results from the BASE-II test-set, MultiAgeEpi was investigated in four independent external cohorts: KORA FF4 (n=1801, mean age: 57.8 years, 53% women), KORA Age (n=1010, mean age: 76.4 years, 50% women), SHIP-TREND (n=971, mean age: 50.8 years, 55% women), and BiDirect (n=557, mean age: 51.9 years, 50% women). Descriptive statistics of the external validation sets are shown in Supplementary Tables 12 to 16. Variables from the external cohorts were selected to reflect the best matched phenotypes of aging originally investigated in BASE-II. To evaluate the results in the context of available epigenetic clocks, MultiAgeEpi was compared with results from SystemsAge (17), GrimAge (11), and DunedinPACE (13). The regression models were adjusted for age, sex, smoking, alcohol intake, BMI, and physical activity and results were corrected for multiple testing using FDR adjustment (Supplementary Tables 17 to 27). Among all investigated clocks, MultiAgeEpi showed the strongest association with T2D and MetS in KORA FF4 (Figure 4A, Supplementary Table 19) and effect sizes stronger or similar to the other clocks in KORA Age and SHIP-TREND (Figure 4B and D, Supplementary Table 17, 18, 20, and 21). In BiDirect, a nominally statistically significant association with T2D was found which, however, did not remain significant after multiple testing correction (Figure 4C, Supplementary Tables 24 and 25). Furthermore, statistically significant associations between MultiAgeEpi and multimorbidity were found in two external cohorts, KORA Age and BiDirect. In contrast, no associations were found with depressive symptoms which, however, was true in almost all cohorts for most of the other investigated clocks, as well. A nominally significant association between MultiAgeEpi and cognitive function was found cross-sectionally in KORA Age and BiDirect as well as longitudinally in SHIP-TREND (Figure 4B, C, and F, Supplementary Tables 17, 22, and 24). Across cohorts and compared to SystemsAge, MultiAgeEpi showed consistently stronger and more frequently statistically significant associations with metabolic disease (T2D, MetS) while SystemsAge was seemingly more strongly associated with variables of cognitive function.

**Figure 4:**
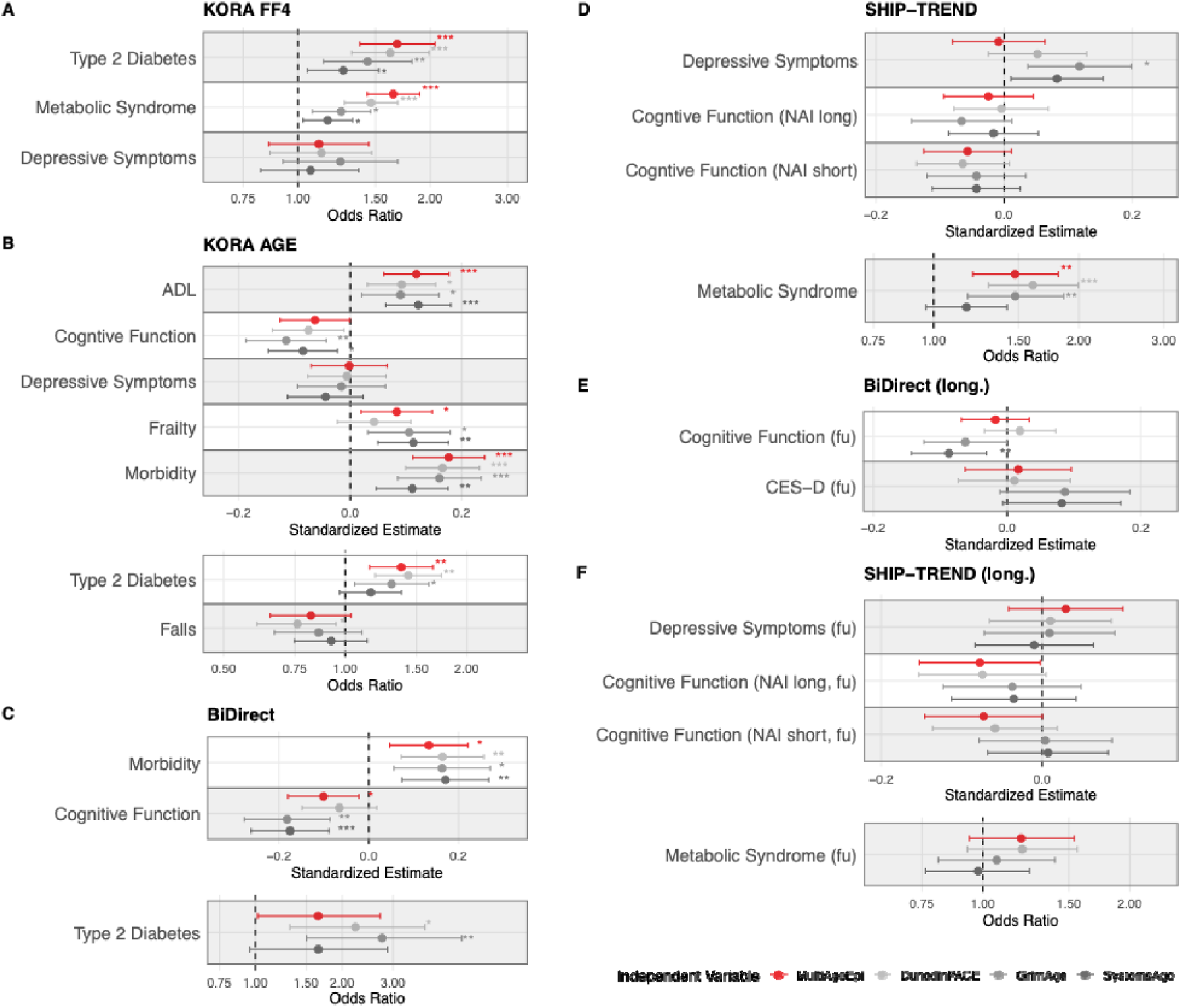
External validation results from MultiAgeEpi in four independent cohorts. Cross-sectional analyses included data from KORA FF4 (A, n=1,801), KORA Age (B, n=1,010), SHIP-TREND (D, n=971), and BiDirect (C, n=557). Additionally, longitudinal analyses were performed using data from BiDirect (E, n=557) and SHIP-TREND (F, n=726). The results of the highest adjusted model (age, sex, alcohol intake, smoking, BMI, and physical activity) are presented here. In longitudinal analyses, the results are additionally adjusted for the outcome at baseline except for CES-D in BiDirect where this information was not available. Note: * q <0.05, ** q <0.01, *** q <0.001.

With respect to mortality, MultiAgeEpi performed in a similar manner to SystemsAge and the investigated epigenetic clocks, GrimAge and DunedinPACE, and was statistically significantly associated with mortality in KORA Age and KORA FF4 (Figure 5, Supplementary Table 27).

**Figure 5:**
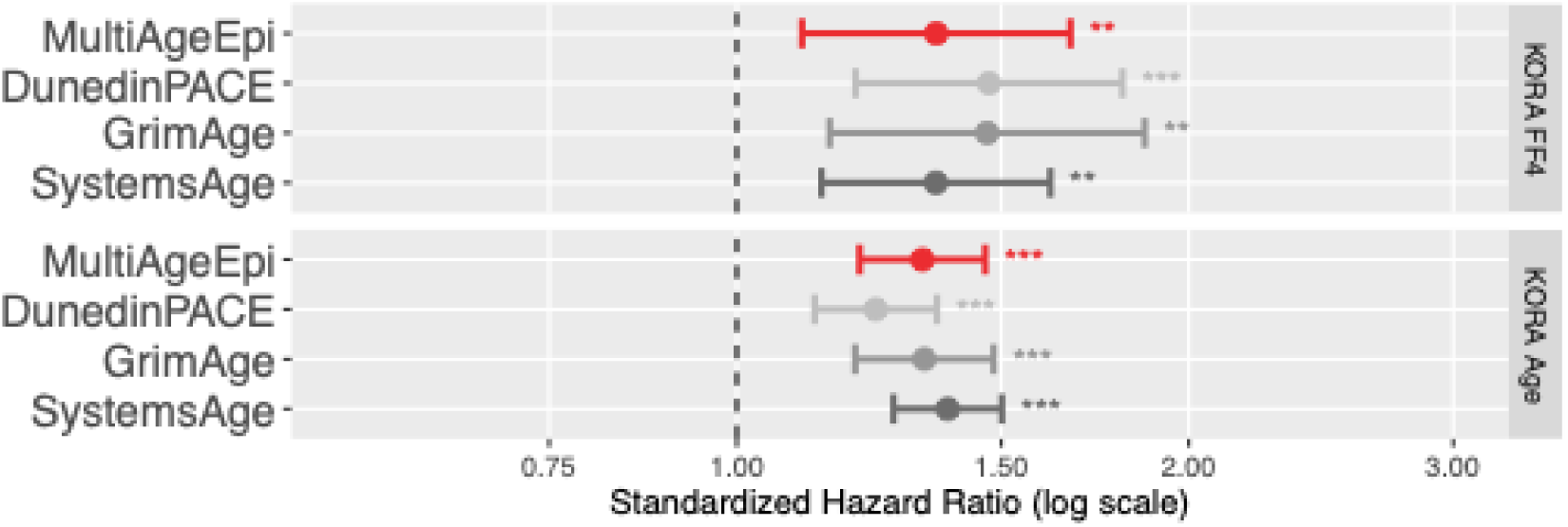
Cox proportional hazard regression models of survival on MultiAgeEpi, SystemsAge, GrimAge, and DunedinPACE in KORA FF4 and KORA Age. Survival data were also available in BiDirect and SHIP-TREND but due to the low number of events, this was not investigated here. The Cox regression was conducted using age as an underlying time scale and adjusted for sex, alcohol, smoking, BMI, and physical activity. Note: * q <0.05, ** q <0.01, *** q <0.001.

In accordance with recommendations for the evaluation of biomarkers of aging (44), we also report the unadjusted (model 1) as well as a age- and sex-adjusted model (model 2) as well as sex-stratified analyses in Supplementary Tables 17 to 27. Compared to the fully adjusted model 3, in most cases, stronger effect sizes were observed in model 1 and 2. A statistically significant association (FDR-adjusted) between MultiAgeEpi and cognitive function was observed in model 1 in SHIP-TREND and KORA Age but was no longer significant after additional confounder adjustment.

## Discussion

In this study, we report on a new, multidimensional biomarker of aging, MultiAge, which was derived from a comprehensive dataset collected from participants of BASE-II. First, we evaluated this newly developed marker cross-sectionally as well as longitudinally in the context of a panel of 17 age-associated outcomes which we previously used to compare established biomarkers of aging in BASE-II (15). Strong and statistically significant associations were found after confounder adjustment and multiple-testing correction with Fried FP, SPRINT-BASEed FI, MI, DCSI, MNA, MMSE, LS7, Tinetti, diagnosed T2D, and MASLD in both cross-sectional and longitudinal analyses. Adding MultiAge to a basic clinical prediction model increased the ability to predict incident cases of disease or impairment over an average follow-up period of 7.4 years by up to 17.5 percentage points (T2D, q <0.001). While previous biomarkers of aging, in most cases, focus on specific aspects of aging (15), MultiAge aggregates comprehensive information regarding biological age in one single value. As such, it was designed to provide a more complete picture of the biological aging status than other available biological age assessments. However, while this marker therefore has potential to improve clinical prediction models, its assessment is laborious, expensive and therefore likely not feasible to be conducted in many other studies or to be translated into a practical/ clinical setting. Therefore, we derived a specific DNA methylation signature, MultiAgeEpi, to facilitate portability of this marker. All results from MultiAgeEpi were compared to the recently published SystemsAge clock, which focuses on the epigenetic representation of biological age in different organ systems and used a smaller set of CpGs (Illumina 450k array). Despite the methodological as well as conceptual differences, MultiAgeEpi performed in a similar manner to SystemsAge as well as the other epigenetic clocks, GrimAge and DunedinPACE, and showed an even stronger association with several of the investigated outcomes (e.g., T2D and MetS in KORA FF4).

In light of the data from the internal validation and the cross-sectional and longitudinal analyses conducted in up to four independent validation cohorts, none of the clocks seems to have a clear advantage over the others. While MultiAgeEpi performs especially well for metabolic diseases (e.g. T2D, MetS), SystemsAge tended to show a stronger association with variables of cognitive function. Future studies investigating these clocks in detail in sufficiently sized longitudinal cohorts with additional outcome variables such as age-associated diseases as well as in the context of their response to potential interventions aimed at promoting healthy aging are needed to better understand possible individual advantages and limitations of these markers.

One of the strengths of MultiAge and MultiAgeEpi is its design based on the comprehensive set of aging-relevant variables collected in BASE-II, which allow for an exceedingly detailed assessment of key aspects of biological age. Also, by training of the DNA methylation signature using the highest-resolution DNAm microarray currently on the market, Illumina’s MethylationEPIC array, state-of-the art methylation data were available which ensures the transferability of results to other cohorts. By restricting the training set to CpGs which are present both in EPIC version 1 and 2, the MultiAgeEpi signature is compatible with both array versions, thus improving applicability across cohorts.

However, this study has a number of limitations. First, compared to other epigenetic clock algorithms, with an n=809 participants, the sample size of the training set used to develop MultiAgeEpi is relatively small. Notwithstanding that, the external validation analyses show highly promising results similar to or at times even exceeding established epigenetic clocks which, while nevertheless illustrating the importance of clinical aging measures, indicates that the biological information of MultiAge is transferred to MultiAgeEpi to a sufficient degree. Second, the age range of the BASE-II cohort (60 to 84 years, Table 1) is comparatively narrow. However, results were replicable in datasets with a wider age range, e.g. KORA FF4 (age range: 38 to 87 years) and SHIP-TREND (age range 20 to 81 years) which increases confidence that the algorithm is applicable to cohorts with different or larger age ranges. Third, the longitudinal analyses in this study were limited to only two cohorts (SHIP-TREND and BiDirect) with restricted availability of outcome variables. Future studies are needed to further evaluate the translational potential of this marker. Fourth, the training sample as well as the four independent replication cohorts include mostly participants of European descent, and all are based in Germany. Future studies need to investigate how the MultiAgeEpi marker performs in different geographical and ethnical contexts. Fifth, no independent evaluation of MultiAge was possible, as the specific variables used for its construction were not fully available in any other cohort. However, by investigating longitudinal regression analyses adjusted for the outcomes at baseline and examining the ability to predict incident cases of age-associated phenotypes, such as T2D and frailty, over a follow-up period of an average of 7.4 years, these results suggest that the MultiAge marker does indeed conserve a comprehensive and close-to-full picture regarding the study participants’ biological age. The transferability to other cohorts was proven using the MultiAgeEpi signature as a proxy for the phenotypically measured MultiAge. It is, however, important to note, that the correlation between MutliAge and MutliAgeEpi was limited suggesting that quite some information of the phenotypic variable was not represented in MultiAgeEpi. Alternative approaches, such as other -omics datasets or a highly informative subset of clinical variables, could be a promising path to increase the preservation of the biological information of MultiAge while simultaneously improving transferability. Finally, we note that there is no gold-standard for how variables for biological age assessment should be selected and how they should be mapped or grouped. By basing our variable mapping on previously published literature, we aimed to incorporate these existing concepts as much as possible but acknowledge that other approaches exist and are equally valid.

In conclusion, our study illustrates that it is possible to conserve information from multiple domains reflecting organ health and global biological age in one variable, MultiAge. This marker showed cross-sectional as well as longitudinal associations with an extensive panel of age-associated outcome variables and was robust against confounder adjustment. The DNA methylation signature trained on this marker, MultiAgeEpi, showed similar and, in several cases, even stronger results compared to other epigenetic clocks in four independent external validation cohorts. Future studies investigating this marker in the context of disease prevention and trialing of interventions aimed at promoting healthy aging will increase our understanding of this markers value in clinical application.

## List of Abbreviations

ADL: Barthel Index (BI) for Activities of Daily Living (ADL)
AUC: Area Under the Curve
BMI: Body Mass Index
CES-D: Center for Epidemiologic Studies Depression Scale
CI: Confidence interval
CpG: Cytosine-phosphate-guanine dinucleotide
DCSI: Diabetes Complications Severity Index
DSST: Digit Symbol Substitution Test
FFD: Fingertips-to-Floor Distance
FI: Frailty Index
FP: Frailty Phenotype
g/d: gram/day
HR: Hazard Ratio
LS7: Life’s Simple 7
MASLD: Metabolic dysfunction–associated steatotic liver disease
Max: Maximum
MetS: Metabolic Syndrome
Min: Minimum
MMSE: Mini-Mental State Examination
MNA: Mini Nutritional Assessment
OR: Odds Ratio
RAPA: Rapid Assessment of Physical Activity
SCORE2: Systematic Coronary Risk Evaluation 2
SD: Standard Deviation
T2D: Type 2 Diabetes

## Declarations

### Ethics approval and consent to participate

#### BASE-II

All participants gave written informed consent. All assessments at baseline and follow-up were conducted in accordance with the Declaration of Helsinki and approved by the Ethics Committee of the Charité—Universitätsmedizin Berlin (approval numbers EA2/029/09, EA2/144/16, and EA2/224/21) and were registered in the German Clinical Trials Registry as DRKS00009277 (BASE-II, registered on August 31, 2015) and DRKS00016157 (GendAge, registered on April 3, 2019).

#### KORA FF4

Ethical approval for the KORA S4/F4/FF4 examinations was granted by the local ethics committee, and written informed consent was obtained from all participants (06068).

### KORA Age

Written informed consent was obtained from all participants, and ethical approval was granted by the local ethics committee (08064).

#### SHIP-TREND

The study was allowed under the recommendations of the Declaration of Helsinki. The medical ethics committee of the University of Greifswald approved the study protocol, and oral and written informed consents were obtained from each of the study participants.

### BiDirect

Written informed consent was obtained from all participants, and ethical approval was granted by the local ethics committee (2009-391-f-S).

## Availability of data and materials

### BASE-II

Due to concerns for participant privacy, BASE-II data are available only upon reasonable request. Additional information can be found on the BASE-II website: https://www.base2.mpg.de/7549/data-documentation. Selected CpGs as well as the weights needed to calculate MultiAgeEpi are available in Supplementary Table 30.

### KORA FF4 and KORA Age

The data from this study are not publicly available due to data protection regulations and restrictions imposed by the Ethics Committee of the Bavarian Chamber of Physicians to protect participant privacy. However, data can be accessed upon request through project agreements with KORA (https://helmholtz-muenchen.managed-otrs.com/external).

### SHIP-TREND

The data of the SHIP-TREND study cannot be made publicly available due to the informed consent of the study participants, but it can be accessed through a data application form available at https://fvcm.med.uni-greifswald.de/ for researchers who meet the criteria for access to confidential data.

### BiDirect

Due to concerns for participant privacy, BiDirect data are available only upon reasonable request. Additional information can be found on the BiDirect website: https://www.medizin.uni-muenster.de/epi/forschung/projekte/bidirect.html

## Funding

### BASE-II

This work was supported by grants from the Deutsche Forschungsgemeinschaft (project number 460683900 to ID and LB), the ERC (as part of the “Lifebrain” project to LB and CML), the Cure Alzheimer’s Fund (as part of the “CIR-CUITS” consortium to LB), and a grant from the EU Joint Programme – Neurodegenerative Disease Research (JPND2021-650-289, coordinator: CML). This article uses data from the Berlin Aging Study II (BASE-II). BASE-II was supported by the German Federal Ministry of Education and Research under grant numbers #01UW0808; #16SV5536K, #16SV5537, #16SV5538, #16SV5837, #01GL1716A, and #01GL1716B. C.M. L. was supported by the Heisenberg program of the DFG (DFG; LI 2654/4-1).

### SHIP-TREND

SHIP is part of the Community Medicine Research Net of the University Medicine Greifswald, Germany, which is funded by the Federal State of Mecklenburg-West Pomerania, and the network ‘Greifswald Approach to Individualized Medicine (GANI_MED)’ funded by the Federal Ministry of Education and Research (grant 03IS2061A). DNA methylation data have been supported by the DZHK (grants 81X3400104, 81X2400157). A.T. has been funded by the Deutsche Forschungsgemeinschaft (DFG, German Research Foundation) – 542489987.

### KORA FF4 and KORA Age

The KORA study was initiated and financed by the Helmholtz Zentrum München – German Research Center for Environmental Health, which is funded by the German Federal Ministry of Research, Technology and Space (BMFTR) and by the State of Bavaria. Data collection in the KORA study is done in cooperation with the University Hospital of Augsburg. The KORA-Age project was financed by the German Federal Ministry of Research, Technology and Space (BMFTR FKZ 01ET0713 and 01ET1003A) as part of the ‘Health in old age’ program.

### BiDirect

The BiDirect Study was funded by grants from the German Federal Ministry of Education and Research (BMBF, grants: FKZ-01ER0816 and FKZ-01ER1506) to the University of Münster.

## Author contributions

Conceptualization: V.M.V, L.B., I.D.; Data curation: V.M.V., I.D.; Formal analysis: V.M.V, M.P.J., G.D., A.L.W., J.H.,; Investigation: V.M.V, M.P.J., J.D., S.D., J.H., E.-M. M., D.S., C.M.L., L.B., I.D; Methodology: V.M.V, C.M.L., L.B., I.D.; Project administration: V.M.V, I.D.; Resources: H.J.G., E,G., U.L., M.N., G.P., A.P., E.S.-T., B.T., H.V., J.W., K.B. A.T., M.W., C.L., L.B., I.D.; Supervision: V.M.V., H.J.G., A.T., M.W., C.M.L., L.B., I.D.; Visualization: V.M.V; Writing - original draft: V.M.V, I.D.; and Writing - review & editing: all authors.

## Data Availability

BASE-II
Due to concerns for participant privacy, BASE-II data are available only upon reasonable request. Additional information can be found on the BASE-II website: https://www.base2.mpg.de/7549/data-documentation. Selected CpGs as well as the weights needed to calculate MultiAgeEpi are available in Supplementary Table 30.
KORA FF4 and KORA Age
The data from this study are not publicly available due to data protection regulations and restrictions imposed by the Ethics Committee of the Bavarian Chamber of Physicians to protect participant privacy. However, data can be accessed upon request through project agreements with KORA (https://helmholtz-muenchen.managed-otrs.com/external).
SHIP-TREND
The data of the SHIP-TREND study cannot be made publicly available due to the informed consent of the study participants, but it can be accessed through a data application form available at https://fvcm.med.uni-greifswald.de/ for researchers who meet the criteria for access to confidential data.
BiDirect
Due to concerns for participant privacy, BiDirect data are available only upon reasonable request. Additional information can be found on the BiDirect website: https://www.medizin.uni-muenster.de/epi/forschung/projekte/bidirect.html

## Acknowledgements

Not applicable.

## Supplementary Figures

**Supplementary Figure 1: Histogram of MultiAge in the sample of BASE-II participants used to develop this marker of biological age (n=1,631).**

**Supplementary Figure 2: Cross-sectional results from linear and logistic regression models of the panel of age-associated outcomes variables on MultiAge and DomainAge variables in BASE-II (n=1,631) at baseline.** Model 1 is unadjusted, Model 2 is adjusted for age and sex, and Model 3 is adjusted for age, sex, alcohol, smoking, BMI, and physical activity. Note: * q <0.05, ** q <0.01, *** q <0.001, T2D = Type 2 Diabetes, MetS = Metabolic Syndrome, MASLD = Metabolic dysfunction–associated steatotic liver disease, FI = Frailty Index, FFD = Fingertips-to-Floor Distance, MMSE = Mini-Mental State Examination, DSST = Digit Symbol Substitution Test, CES-D = Center for Epidemiologic Studies Depression Scale, ADL = Barthel Index (BI) for Activities of Daily Living (ADL), MNA = Mini Nutritional Assessment, DCSI = Diabetes Complications Severity Index, SCORE2 = Systematic Coronary Risk Evaluation 2, LS7 = Life’s Simple 7.

**Supplementary Figure 3: Regression analyses comparing the association of MultiAge and MultiAgeEpi with the panel of outcome variables in the BASE-II hold-out test set.** The investigated test-set was separated from the training set in an 80:20-split and was therefore not part of the the data used to train MultiAgeEpi. The presented results are adjusted for age, sex, alcohol, smoking, BMI, and physical activity. Note: * q <0.05, ** q <0.01, *** q <0.001.

**Supplementary Figure 4: Flow chart illustrating the sample sizes of the BASE-II dataset used to develop MultiAge and train and test MultiAgeEpi.** Of the full n=2,171 people who participated in the medical part of BASE-II, n=500 were not analyzed as they belonged to the younger control group and n=40 had in more than 50% of the variables selected for MultiAge construction missings.

**Supplementary Figure 5: Results of linear and logistic regression analyses of age-associated outcomes on MultiAge aggregated using five statistical methods.** All results are adjusted for age, sex, smoking, alcohol, BMI, and physical activity. Note: T2D = Type 2 Diabetes, MetS = Metabolic Syndrome, MASLD = Metabolic dysfunction–associated steatotic liver disease, FI = Frailty Index, FFD = Fingertips-to-Floor Distance, MMSE = Mini-Mental State Examination, DSST = Digit Symbol Substitution Test, CES-D = Center for Epidemiologic Studies Depression Scale, ADL = Barthel Index (BI) for Activities of Daily Living (ADL), MNA = Mini Nutritional Assessment, DCSI = Diabetes Complications Severity Index, SCORE2 = Systematic Coronary Risk Evaluation 2, LS7 = Life’s Simple 7.

**Supplementary Figure 6: Results from longitudinal regression models of outcomes at T1 on DomainAge variables at T0 in BASE-II (n=1,072).** Results are shown with adjustment for the outcome variable at T0 (model 1), adjusted for the outcome variable at T0, age, and sex (model 2), and adjusted for outcome at T0 age, sex, alcohol, smoking, BMI, and physical activity (model 3). Associations with MultiAge is shown in Figure 2 in the main text. Note: * q <0.05, ** q <0.01, *** q <0.001.

**Supplementary Figure 7: Results from regression models using the DomainAge variables to predict incident cases of disease or impairment over the on-average 7.4 year follow-up period and survival.** Participants with prevalent disease or impairment in the respective variables were excluded from this analysis (therefore, the sample size varies between outcome variables). Model 1 is unadjusted, Model 2 is adjusted for age and sex, and Model 3 is adjusted for age, sex, alcohol, smoking, BMI, and physical activity. Sex-stratified analyses as well as results from DomainAge variables are reported in Supplementary Table 7 and 8. The results using MultiAge instead of the DomainAge variables are visualized in Figure 3 of the main text. Note: * q <0.05, ** q <0.01, *** q <0.001.

## References

1. Kusters CD, Horvath S. Quantification of epigenetic aging in public health. Annual review of public health. 2024;46.

2. Herzog C, Poganik JR, Barzilai N, Basisty N, Beerman I, Belsky DW, et al. Biomarkers of Aging–NIA Joint Symposium 2024: New Insights Into Aging Biomarkers. Aging Cell. 2025:e70124.

3. Kennedy BK, Berger SL, Brunet A, Campisi J, Cuervo AM, Epel ES, et al. Geroscience: Linking Aging to Chronic Disease. Cell. 2014;159(4):709–13.

4. Lohman T, Bains G, Berk L, Lohman E. Predictors of Biological Age: The Implications for Wellness and Aging Research. Gerontology and Geriatric Medicine. 2021;7:23337214211046419.

5. Ferrucci L, Levine ME, Kuo P-L, Simonsick EM. Time and the Metrics of Aging. Circulation Research. 2018;123(7):740–4.

6. Jylhava J, Pedersen NL, Hagg S. Biological Age Predictors. EBioMedicine. 2017;21:29–36.

7. Silva N, Rajado AT, Esteves F, Brito D, Apolónio J, Roberto VP, et al. Measuring healthy ageing: current and future tools. Biogerontology. 2023;24(6):845–66.

8. Lowsky DJ, Olshansky SJ, Bhattacharya J, Goldman DP. Heterogeneity in Healthy Aging. The Journals of Gerontology: Series A. 2013;69(6):640–9.

9. Horvath S. DNA methylation age of human tissues and cell types. Genome biology. 2013;14(10):R115.

10. Hannum G, Guinney J, Zhao L, Zhang L, Hughes G, Sadda S, et al. Genome-wide methylation profiles reveal quantitative views of human aging rates. Molecular cell. 2013;49(2):359–67.

11. Lu AT, Quach A, Wilson JG, Reiner AP, Aviv A, Raj K, et al. DNA methylation GrimAge strongly predicts lifespan and healthspan. Aging (Albany NY). 2019;11(2):303.

12. Levine ME, Lu AT, Quach A, Chen BH, Assimes TL, Bandinelli S, et al. An epigenetic biomarker of aging for lifespan and healthspan. Aging (Albany NY). 2018;10(4):573.

13. Belsky DW, Caspi A, Corcoran DL, Sugden K, Poulton R, Arseneault L, et al. DunedinPACE, a DNA methylation biomarker of the pace of aging. eLife. 2022;11:e73420.

14. Mavrommatis C, Belsky DW, Ying K, Moqri M, Campbell A, Richmond A, et al. An unbiased comparison of 14 epigenetic clocks in relation to 174 incident disease outcomes. Nature Communications. 2025;16(1):11164.

15. Vetter VM, Drewelies J, Homann J, Düzel S, Deecke L, Jawinski P, et al. Comprehensive Comparison of Sixteen Markers of Biological Aging: Cross-Sectional and Longitudinal Results from the Berlin Aging Study II (BASE-II). medRxiv. 2025:2025.04.09.25325514.

16. Vetter VM, Junge MP, Drevon CA, Gundersen TE, Homann J, Lill CM, et al. Comparing fourteen consensus biomarkers of aging: epigenetic pace of aging as the strongest predictor of mortality in BASE-II. Biomarker Research. 2026.

17. Sehgal R, Markov Y, Qin C, Meer M, Hadley C, Shadyab AH, et al. Systems Age: A single blood methylation test to quantify aging heterogeneity across 11 physiological systems. Nature Aging. 2025;5(9):1880–96.

18. Bafei SEC, Shen C. Biomarkers selection and mathematical modeling in biological age estimation. npj Aging. 2023;9(1):13.

19. Seeman TE, Singer BH, Rowe JW, Horwitz RI, McEwen BS. Price of adaptation—allostatic load and its health consequences: MacArthur studies of successful aging. Archives of internal medicine. 1997;157(19):2259–68.

20. Klemera P, Doubal S. A new approach to the concept and computation of biological age. Mechanisms of Ageing and Development. 2006;127(3):240–8.

21. Nakamura E, Miyao K, Ozeki T. Assessment of biological age by principal component analysis. Mechanisms of Ageing and Development. 1988;46(1):1–18.

22. Carmel S. Health and well-being in late life: Gender differences worldwide. Frontiers in medicine. 2019;6:218.

23. Hägg S, Jylhävä J. Sex differences in biological aging with a focus on human studies. eLife. 2021;10:e63425.

24. Garmany A, Yamada S, Terzic A. Longevity leap: mind the healthspan gap. npj Regenerative Medicine. 2021;6(1):57.

25. Partridge L, Deelen J, Slagboom PE. Facing up to the global challenges of ageing. Nature. 2018;561(7721):45–56.

26. Jagger C, Gillies C, Moscone F, Cambois E, Van Oyen H, Nusselder W, et al. Inequalities in healthy life years in the 25 countries of the European Union in 2005: a cross-national meta-regression analysis. The Lancet. 2008;372(9656):2124–31.

27. Goldman DP, Cutler D, Rowe JW, Michaud P-C, Sullivan J, Peneva D, et al. Substantial health and economic returns from delayed aging may warrant a new focus for medical research. Health affairs. 2013;32(10):1698–705.

28. Bertram L, Bockenhoff A, Demuth I, Duzel S, Eckardt R, Li SC, et al. Cohort profile: The Berlin Aging Study II (BASE-II). International journal of epidemiology. 2014;43(3):703–12.

29. Demuth I, Banszerus V, Drewelies J, Düzel S, Seeland U, Spira D, et al. Cohort profile: follow-up of a Berlin Aging Study II (BASE-II) subsample as part of the GendAge study. BMJ Open. 2021;11(6):e045576.

30. Lopez-Otin C, Blasco MA, Partridge L, Serrano M, Kroemer G. The hallmarks of aging. Cell. 2013;153(6):1194–217.

31. Hägg S, Belsky DW, Cohen AA. Developments in molecular epidemiology of aging. Emerging Topics in Life Sciences. 2019;3(4):411–21.

32. Lara J, Cooper R, Nissan J, Ginty AT, Khaw K-T, Deary IJ, et al. A proposed panel of biomarkers of healthy ageing. BMC medicine. 2015;13(1):1–8.

33. Margolick JB, Ferrucci L. Accelerating aging research: how can we measure the rate of biologic aging? Experimental gerontology. 2015;64:78–80.

34. Engelfriet PM, Jansen EHJM, Picavet HSJ, Dollé MET. Biochemical Markers of Aging for Longitudinal Studies in Humans. Epidemiologic Reviews. 2013;35(1):132–51.

35. Vetter VM, Drewelies J, Homann J, Düzel S, Deecke L, Jawinski P, et al. Comprehensive cross-sectional and longitudinal comparison of sixteen markers of biological aging from the Berlin Aging Study II. Communications Medicine. 2026;6(1):168.

36. Linkohr B, Heier M, Gieger C, Thorand B, Grallert H, Holle R, et al. Cohort Profile: Cooperative Health Research in the Region of Augsburg (KORA) 1984-2024. Int J Epidemiol. 2025;54(6).

37. Holle R, Happich M, Löwel H, Wichmann H-E, Group MKS. KORA-a research platform for population based health research. Das Gesundheitswesen. 2005;67(S 01):19–25.

38. Peters A, Döring A, Ladwig K, Meisinger C, Linkohr B, Autenrieth C, et al. Multimorbidity and successful aging: the population-based KORA-Age study. Zeitschrift fur Gerontologie und Geriatrie. 2011;44:41–54.

39. Steinbeisser K, Grill E, Holle R, Peters A, Seidl H. Determinants for utilization and transitions of long-term care in adults 65+ in Germany: results from the longitudinal KORA-Age study. BMC geriatrics. 2018;18(1):172.

40. Vetter VM, Spira D, Banszerus VL, Demuth I. Epigenetic clock and leukocyte telomere length are associated with vitamin D status, but not with functional assessments and frailty in the Berlin Aging Study II. The Journals of Gerontology: Series A. 2020.

41. Vetter VM, Kalies CH, Sommerer Y, Spira D, Drewelies J, Regitz-Zagrosek V, et al. Relationship Between 5 Epigenetic Clocks, Telomere Length, and Functional Capacity Assessed in Older Adults: Cross-Sectional and Longitudinal Analyses. The Journals of Gerontology: Series A. 2022;77(9):1724–33.

42. Ying K, Paulson S, Eames A, Tyshkovskiy A, Li S, Eynon N, et al. A unified framework for systematic curation and evaluation of aging biomarkers. Nature Aging. 2025;5(11):2323–39.

43. Moqri M, Herzog C, Poganik JR, Justice J, Belsky DW, Higgins-Chen A, et al. Biomarkers of aging for the identification and evaluation of longevity interventions. Cell. 2023;186(18):3758–75.

44. Moqri M, Herzog C, Poganik JR, Ying K, Justice JN, Belsky DW, et al. Validation of biomarkers of aging. Nature Medicine 2024.

45. Spira D, Buchmann N, König M, Rosada A, Steinhagen-Thiessen E, Demuth I, et al. Sex-specific differences in the association of vitamin D with low lean mass and frailty–Results from the Berlin Aging Study II. Nutrition. 2018.

46. Fried LP, Tangen CM, Walston J, Newman AB, Hirsch C, Gottdiener J, et al. Frailty in older adults: evidence for a phenotype. The Journals of Gerontology Series A: Biological Sciences and Medical Sciences. 2001;56(3):M146–M57.

47. Pajewski NM, Williamson JD, Applegate WB, Berlowitz DR, Bolin LP, Chertow GM, et al. Characterizing Frailty Status in the Systolic Blood Pressure Intervention Trial. The Journals of Gerontology: Series A. 2016;71(5):649–55.

48. Vetter VM, Drewelies J, Düzel S, Homann J, Meyer-Arndt L, Braun J, et al. Change in body weight of older adults before and during the COVID-19 pandemic: Longitudinal results from the Berlin Aging Study II. The Journal of nutrition, health and aging. 2024;28(4):100206.

49. Meyer A, Salewsky B, Spira D, Steinhagen-Thiessen E, Norman K, Demuth I. Leukocyte telomere length is related to appendicular lean mass: cross-sectional data from the Berlin Aging Study II (BASE-II). Am J Clin Nutr. 2016;103(1):178–83.

50. Vellas B, Guigoz Y, Garry PJ, Nourhashemi F, Bennahum D, Lauque S, et al. The Mini Nutritional Assessment (MNA) and its use in grading the nutritional state of elderly patients. Nutrition. 1999;15(2):116–22.

51. Folstein MF, Folstein SE, McHugh PR. “Mini-mental state”. A practical method for grading the cognitive state of patients for the clinician. J Psychiatr Res. 1975;12(3):189–98.

52. Lloyd-Jones DM, Hong Y, Labarthe D, Mozaffarian D, Appel LJ, Van Horn L, et al. Defining and setting national goals for cardiovascular health promotion and disease reduction: the American Heart Association’s strategic Impact Goal through 2020 and beyond. Circulation. 2010;121(4):586–613.

53. König M, Drewelies J, Norman K, Spira D, Buchmann N, Hülür G, et al. Historical trends in modifiable indicators of cardiovascular health and self-rated health among older adults: Cohort differences over 20 years between the Berlin Aging Study (BASE) and the Berlin Aging Study II (BASE-II). PLoS one. 2018;13(1):e0191699.

54. Tinetti ME, Williams TF, Mayewski R. Fall risk index for elderly patients based on number of chronic disabilities. The American journal of medicine. 1986;80(3):429–34.

55. Young BA, Lin E, Von Korff M, Simon G, Ciechanowski P, Ludman EJ, et al. Diabetes complications severity index and risk of mortality, hospitalization, and healthcare utilization. The American journal of managed care. 2008;14(1):15.

56. Spieker J, Vetter VM, Spira D, Steinhagen-Thiessen E, Regitz-Zagrosek V, Buchmann N, et al. Diabetes Type 2 in the Berlin Aging Study II: Prevalence, Incidence and Severity Over up to Ten Years of Follow-up. PREPRINT (Version 3) available at Research Square. 2021.

57. Alberti KG, Eckel RH, Grundy SM, Zimmet PZ, Cleeman JI, Donato KA, et al. Harmonizing the metabolic syndrome: a joint interim statement of the International Diabetes Federation Task Force on Epidemiology and Prevention; National Heart, Lung, and Blood Institute; American Heart Association; World Heart Federation; International Atherosclerosis Society; and International Association for the Study of Obesity. Circulation. 2009;120(16):1640–5.

