## Supplementary Material for "MultiAge: A New Multidimensional Biomarker of Biological Age Derived from Comprehensive Phenotypic and Molecular Profiling"

#### Supplementary Methods

### BASE-II

#### Development of MultiAge

##### Study Population

The observational, longitudinal Berlin Aging Study II (BASE-II) (1) consists of 1,671 older participants between 60 and 80 years and a younger cohort of 500 participants between 20 and 35 years (not analyzed in this study). Participants were recruited though the Max Planck Institute for Human Development's participant pool in Berlin and via advertisements in public transportation networks and local newspapers. Following up on the baseline assessment (T0, 2009 – 2014), 1,083 participants of the older group were re-assessed on average 7.4 years later (SD: 1.5 years, range: 3.9 to 10.4 years) as part of the GendAge study (2018 – 2020) (2). Between assessments, 588 participants dropped out. Of those, 126 were confirmed to have died based on the mortality update (updated twice per year). Reasons for dropout among the other 462 participants are not available. It was previously shown that differences between participants who dropped-out and those who were followed-up were small (3).

##### Data cleaning

The distribution of all variables was checked visually. The log-transformed version of variables was used in subsequent analyses if the transformed variable fit the normal distribution better than the original value. If no normal distribution was observed after transformation the respective variable was flagged and excluded from all statistical procedures (i.e. variable aggregation steps) which prerequisite normal distribution. Variables with constant values or very few values indicating suboptimal values were excluded, because they are not expected to add information to the aggregated value due to their small variance. Participants for which more than 50% of the required values were missing were excluded from the imputation and all subsequent analyses. This led to the exclusion of n=40 participants from the dataset resulting in n=1,671 – 40 = 1,631 participants who were investigated. For 11 of the excluded 40 participants, Illumina methylation data were available at baseline effectively reducing the dataset used for training and testing the DNA methylation algorithm by 11 participants.

##### Missing values

Of all observed values, 9.3% were missing. Missing data was visualized using the vis_miss function (visdat package) (4). The connectedness of missing values with the observed data (influx) as well as the connectedness of observed data with missing values (outflux) were calculated using the *flux* function (mice package (5)). Influx represents the ability of a variable to be imputed from the other, observed values. Outflux represents how suitable a variable is to impute other variables missing values. Generally, low influx and high outflux values were observed indicating a high degree of connection of missing data with available data (influx) and high degree of connection between observed data and missing values (outflux).

#### Imputation

In the BASE-II data used to develop MultiAge and MultiAgeEpi (as well as all DomainAge variables), sex-stratified multiple imputation was done using the *mice* function (mice package (5)) using all data available at baseline after exclusion of participants with more than 50% missing values (n=1,631). Using the *quickpred* function (minuc=0.1, mincor=0.075), a prediction matrix with a median of 29 (men) and 31 (women) predictors per variable was build. Each prediction model was forced to include chronological age (m=10, maxit = 20, method = "pmm"). Convergence of the mice algorithm was checked visually by plotting mean and SD over the imputation iterations. Subsequently, the distribution of imputed values in the first imputed dataset in relation to measured (i.e., non-missing) values were visually observed in histograms. Additionally, all imputed values were examined with respect to unequal distribution in context of observed values in scatter plots against chronological age. Imputation as well as all checks on the imputed data were done stratified by sex.

#### Variable processing

To assess whether each participant showed "good" or "bad" values in each of the available variables in comparison to peers of the same age and sex, we calculated sex-stratified residuals from a linear regression of each variable on chronological age. These residuals reflect how far each individual differs from the value that would be expected for the respective variable based on their chronological age and the distribution of values in the whole dataset.

#### Multicollinearity within each domain of aging

To avoid inclusion of variables in one domain which correlated strongly, correlation between variables within one domain was checked. Pearson correlation coefficients in all imputed datasets were calculated and subsequently pooled using the *mcombine.cor* function (miceadds package). If two variables presented a correlation coefficient higher than |0.8|, one was excluded from subsequent aggregation of variables to domain ages. This procedure led to modifications to the following domains:

- Cardiovascular System (excluded: HDL, LDL)
- Metabolism (excluded: LDL, Apolipoprotein A1)
- Musculoskeletal System (excluded: BMI, body weight)
- Brain (excluded: working memory)

#### Aggregation Methods

##### *Quartile-based method (QBM)*

For all variables, the high-risk group was defined based on literature search and subject matter knowledge. This was done independently by two scientists and results were compared subsequently. Discrepancies were then discussed and a consensus was reached after further literature search. Grouping into high-risk groups was done by either specifying the highest quartile or the lowest quartile. For variables which indicate adverse state when values are either too high or too low, participants who fell on the 87.5$^{th}$ or higher or on the 12.5$^{th}$ or lower percentile were given a point. If no cut-off based on the statistical distribution seems suitable, a clinically relevant cut-off was chosen. If the variable was binary (e.g. diagnosed metabolic syndrome), participants with the diagnosis were given a point.

The points were given for each imputed dataset individually, resulting in m=10 datasets with either 0=”not impaired” or 1=”impaired“ point for each variable in the dataset. Then, the m=10 datasets were combined into an array using the *abind* function (abind package) and the mean and SD of the value of each cell was calculated across all 10 datasets. In other words, each of the 10 available values for each participant for each variable was used to calculate the mean and SD. It is important to note, that although only the values of 0 and 1 were assigned within each of the m=10 imputed datasets, due to the averaging of the assigned points across all m=10 datasets, values between 0 and 1 are possible for variables that had a missing value for the respective participant before multiple imputation. This, however, allows to assign more nuanced values for missing values in which it is less obvious if the participant would be best assigned to a high-risk group after multiple imputations. Subsequently, the DomainAge was calculated as average across all individual variables within the respective domain.

##### *Klemera-Doubal Methode (KDM)*

The method to calculate biological age by Klemera and Doubal (6) uses chronological age not as an outcome but as a predictor and thereby makes use of the reverse regression technique proposed by Hochschild (7). Here, the *kdm_calc* function from the “bioage” package by Kwon and Belsky was used (8). As the KDM makes use of multiple linear regression models, their assumptions should be met for the association between the phenotypes and chronological age. Therefore, the raw phenotype values instead of the residuals were used to calculate biological age.

##### *Multiple Linear Regression (MLR)*

Individual linear regression models with chronological age as dependent variable and variables of each aging domain were calculated using R’s *lm* function. As a linear association between the dependent and independent variables is assumed, the original raw values of the phenotypes were used instead of the residuals. The weights of the resulting model were then used to estimate the outcome in the same data using the *predict* function. It was noted, that a systematic error is observable at the regression edges that leads to an underestimation of age for older participants and an overestimation of age for the younger participants when estimated using linear regression models (9, 10). To correct for this distortion, Dubina and colleagues provided a correction method to avoid this systematic error (10). For each of the imputed datasets, individual DomainAge variables were calculated based on the MLR method. For the final analysis, the average across all calculated DomainAge variables across all imputed datasets was calculated to get one final dataset. To transform the results to a scale comparable to those of the other markers, the age acceleration was calculated, similarly to the procedure described for the epigenetic age acceleration, as residuals of linear regression analysis on chronological age.

##### *Homeostatic Dysregulation (HD)*

To calculate the HD, the *hd_calc* function from the BioAge package (8) was used. The z-transformed residuals were used as input and the BASE-II dataset was used as reference population. The multidimensional distance of the variable values of each individual from the mean of the complete dataset was assessed. As the z-transformed residuals are used as input, this corresponds to the age-adjusted sex-specific mean of the group which is, in our view, the optimal reference group in light of a missing external reference dataset from the same source population and with the same phenotypes available.

##### *Principal Components Analysis (PCA)*

The use of a PCA to construct biological age was first proposed by Nakamura and colleagues in 1988 (11) and they recommended to use the first PC as biological estimate. To allow a more intuitive interpretation of the result, the transformation to the scale of chronological age was done. Later, Jia and colleagues (12) added to this method by presenting a formula which standardizes the phenotype values and by suggesting to correct the biological age estimated by using the method developed by Dubina and colleagues (10). In this study, the PCA on the correlation matrix was calculated using R's *princomp* function. Subsequently, the score as well as the loading factors were extracted to calculate the biological age variables based on the methods outlined by Nakamura et al. (11) and Jia et al. (12). Following the procedure described by Nakamura et al. (11), the crude PCA biological age variables were scaled to have a mean of 0 and a SD of 1. Then the following formula was used to project the crude results on the same scale as chronological age: $BA = (BS \times SD_{cAge}) + Mean_{cAge}$. To transform the values to a scale comparable with the other markers, the age acceleration was calculated as residuals from a linear regression analysis on chronological age.

#### Training and internal validation of MultiAgeEpi

##### Pre-processing of methylation data

Methylation data in this study was measured using the Illumina methylation array EPIC version 1. In summary, CpG probes were removed if ≥1% of all investigated samples had a detection *p*-value above 0.05 or if the bead count was below 3 in more than 5% of samples. The efficiency of bisulfite conversion was evaluated using *bscon* (bigmelon package), and samples with conversion rates below 80% were excluded from further analysis. Following outlier removal, the remaining samples were reloaded and normalized using the *dasen* function. We then used the *qual* function to assess shifts in beta values caused by normalization, excluding any samples with absolute deviations ≥0.1 in beta values post-normalization. The dataset, now excluding flagged CpG probes, was reloaded, re-normalized, and subsequently used for DNA methylation age estimation and training of MultiAgeEpi. To facilitate the estimation of the MultiAgeEpi in methylation data obtained from both the Illumina methylation EPIC array version 1 and the newer version 2, we excluded all CpGs from our dataset which are not present in the EPIC version 2. The list of CpGs that was excluded was downloaded from the Illumina website on February 4$^{th}$, 2025. Before QC, 865,918 CpG positions were available. After QC, our methylation dataset contained information on 763,923 CpGs. From EPIC version 1 to EPIC version 2, 143,462 CpG positions were excluded of which 64,606 were part of our methylation data set after QC and therefore resulted in our final methylation dataset containing 699,317 CpGs. After restricting the DNA methylation dataset to the CpGs present on EPICv1 and EPICv2, it was merged with the DomainAge and MultiAge variables and a 80:20 training-test set split was done using R's sample function.

#### Elastic-Net Regression

The 80% training set was used to train an elastic net regression model with DomainAge and MultiAge as outcome. To do so, an alpha of 0.5 was used as it was done for the development of previous epigenetic clocks by Belsky and colleagues (13) and Horvath(14). A tuning grid with alpha = 0.5 and lambda between 0 and 1 in 20 log(10) steps was defined and used to identify the optimal lambda using a 10-fold cross-validation within the training set using the *train* function from the caret package. The best lambda was then used to calculate the final model using the *glmnet* function in the training data. To evaluate the model fit, 20% test previously separated from the training set was used (hold-put test-set). Root-Mean-Squared-Error, Mean Absolute Error (MAE), $R^2$, and Pearson's correlation coefficients were then used to assess model fit.

#### Statistical Analyses

*Evaluation of phenotypic MultiAge and epigenetic MultiAgeEpi*

To analyze MultiAge and the DomainAge aggregates in their phenotypic as well as epigenetic form, linear and logistic regression as well as Cox regression analyses were conducted. Missing values were imputed using the same procedure as described above using all dependent and independent variables. Phenotypes of interest were dichotomized as described in ref. (3) if they were originally on a continuous scale. To allow easy comparison between outcomes on different scales, the complete dataset was normalized prior to regression modelling using the *umx_scale* function after transforming the mids object to a list using the *complete* function. For the calculation of incident cases all participants with a prevalent outcome of interest at T0 were excluded from the analyses. ROC statistics from logistic regressions models were calculated using the pROC package for each imputed dataset and subsequently pooled using the *pool_auc* function (psfmi package). Difference between AUCs was assessed using the *lrtest* function (lmtest package). Difference between Cox proportional hazard models was done using the *anova* function. To visualize the result tables and allow a more intuitive interpretation, forest plots were drawn using ggplot2.

Association with mortality was done by calculating Cox Proportional Hazard regression models on chronological age as underlying time-scale using the *coxph* function. To investigate predictive ability of the models, the c-index was reported. To assess statistical significance of differences in the c-index between the clinical comparison model and the respective model including the biomarkers, the *anova* function from the survival package was used. Multiple testing correction was done by controlling the False Discovery Rate (FDR) using the Benjamini-Hochberg approach. Statistical significance was defined at FDR-adjusted p-value <0.05.

### BASE-II baseline variables

| **Variables** | **Description** |
| --- | --- |

| | |
|---|---|
| Barthel Index (BI) for Activities of Daily Living (ADL) | The Barthel Index (15), is a 10-item assessment that measures an individual's ability to independently carry out everyday activities. The instrument includes questions regarding personal hygiene, continence, dressing, eating, and mobility. |
| Adrenomedullin (ADM) | Because directly measured adrenomedullin was not available in BASE-II, DNA methylation–based Adrenomedullin (DNAmADM) was derived from Illumina MethylationEPIC array data using the method described by Lu and colleagues (16). |
| Alanine aminotransferase (ALT) | Alanine aminotransferase was measured using an assay in accordance with the recommendations of the International Federation of Clinical Chemistry and Laboratory Medicine (IFCC). |
| Albumin | Albumin was measured via an immunoturbidimetric assay (Roche Diagnostics, cobas, Rotkreuz, Switzerland). |
| Albuminuria | See Albumin. |
| Alcohol | Alcohol consumption in gram per day (g/d) was calculated from the food-frequency questionnaire from the European Prospective Investigation into Cancer and Nutrition (17). |
| Alkaline phosphatase (ALP) | Alkaline Phosphatase was measured using a colorimetric assay in accordance with a standardized method (Roche Diagnostics, cobas, Rotkreuz, Switzerland). |
| Allostatic Load Index (ALI) | ALI was calculated using the "Group ALI" approach (18) which is the most used version of the ALI (19). Six groups of variables were considered: neuroendocrine (cortisol, dehydroepiandrosterone sulfate (DHEA-S)), inflammation (C-reactive protein), metabolic (high-density lipoprotein cholesterol (HDL-C), low-density lipoprotein cholesterol (LDL-C), triglycerides (TG), glycosylates hemoglobin (HbA1c), fasting glucose), renal (creatinine), cardiovascular (systolic blood pressure (SBP), diastolic blood pressure (DBP), resting heart rate (RHR)), and anthropometric (waist-hip-ratio (WHR), body mass index (BMI)). Participants who were in the sex-stratified high-risk quartile (>75$^{th}$ percentile or <25$^{th}$ percentile) were awarded one point resulting in a possible value between 0 and 14 points for each participant. To assess possible dysregulation in one of the listed variables which however would be masked by a successful pharmacological therapy (20, 21), participants were given one point if the following medication intake was reported: LDL-C (statins, cholesterol absorption inhibitors, niacin or bile |

| | |
|---|---|
| | sequestrants), TG (fibrates), HbA1c and fasting glucose (antidiabetic medication), SBP (antihypertensive medication), and RHR (beta-blockers, calcium channel blockers, cardiac glycosides, amiodaron). The individual variables and corresponding medication were chosen based on previous publications The contributing parameters and considered medications were chosen in accordance with the publications by McCrory et al. and Seeman et al. (18, 20, 21). |
| Alpha-1 Globulin | Alpha-1-Globuin, Alpha-2-Globulin, Beta-Globulin, and Gamma-Globulin were measured via electrophoresis using the CAPILLARYS system (Sebia, Mainz, Germany). |
| Alpha-2 Globulin | See Alpha-1-Globulin. |
| Alpha-Amylase | Alpha-Amylase was measured using an enzymatic colorimetric assay which was designed following the recommendation by The International Federation of Clinical Chemistry and Laboratory Medicine (IFCC) (Roche Diagnostics, cobas, Rotkreuz, Switzerland). |
| Apolipoprotein A1 | Apolipoprotein A1 was measured using an immunoturbidimetric assay (Tina-quant Apolipoprotein A-1 ver.2, Roche Diagnostics, Rotkreuz Switzerland). |
| Apolipoprotein B | Apolipoprotein B was measured using an immunoturbidimetric assay (Tina-quant Apolipoprotein B ver.2, Roche Diagnostics, Rotkreuz Switzerland). |
| Aspartate transaminase (AST) | Aspartate transaminase was measured using an assay in accordance with the recommendations of the International Federation of Clinical Chemistry and Laboratory Medicine (IFCC). |
| Augmentation Index (AIx) | Pulse wave analysis (PWA) was conducted using the Mobil-O-Graph device (I.E.M. Germany) with participants seated in a calm, relaxed setting, in a room kept as quiet as possible, in accordance with international guidelines. The central augmentation index (AIx) was calculated as the difference between the second and first systolic peaks of the central arterial waveform, expressed as a percentage of pulse pressure. It reflects wave reflection characteristics and arterial stiffness, being primarily driven by large artery stiffness while also affected by peripheral resistance, body height, heart rate, and sex. Additional information can be found elsewhere (22). |

| | |
|---|---|
| Beta-2 microglobulin | Because directly measured Beta-2 microglobulin was not available in BASE-II, DNA methylation-based Beta-2 microglobulin (DNAmB2M) was derived from Illumina MethylationEPIC array data using the method described by Lu and colleagues (16). |
| Beta Globulin | See Alpha-1 Globulin. |
| Total Bilirubin | Total Bilirubin was measured using the colorimetric Diazo-Method (Roche Diagnostics, cobas, Rotkreuz, Switzerland). |
| BioAge | BioAge is a composite marker that includes the following 12 weighted variables which can be obtained from a routine laboratory report: zinc, sodium, chloride, uric acid, albumin, alpha-1 globulin, alpha-2 globulin, HbA1c, hemoglobin, leukocytes, lymphocytes, and creatinine. This marker was developed in BASE, a completely independent sample assessed prior to the BASE-II participants analyzed in this study. (23) |
| Body Mass Index (BMI) | BMI was calculated using bodyweight and height ($kg/m^2$) measured with the 763 seca measuring station (SECA, Hamburg, Germany) |
| Body Weight | Body weight measured in kilogram with the 763 seca measuring station (SECA, Hamburg, Germany) |
| Bone Mineral Density (BMD) | Bone Mineral Density was measured from data obtained through dual-energy X-ray absorptiometry (Hologic QDR Discovery; Hologic Inc., Bedford, MA). The measurement was performed by a trained technician and femoral neck bone mineral density was used for further analyses. |
| Calcium | Calcium was measured by allowing calcium ions to react with 5-nitro-5'-methyl-BAPTA (NM-BAPTA) under alkaline conditions to form a complex, which then undergoes a subsequent reaction with EDTA. The change in absorbance is directly proportional to the calcium concentration and is measured photometrically (Roche Diagnostics GmbH, Rotkreuz, Switzerland). |
| Center for Epidemiologic Studies Depression Scale (CES-D) | The Center for Epidemiological Studies Depression Scale (CES-D) is a 20-item questionnaire which covers, among others, the frequency of experiencing a depressed mood, feelings of guilt/worthlessness, feelings of helplessness/hopefulness, and psychomotor retardation. Each of the 20 items is scored with a point between 1 and 4, resulting in maximum of 60 points (24). |

| | |
|---|---|
| Chloride | See Potassium |
| Cholinesterase (ChE) | Cholinesterase was measured using a colorimetric assay (Roche Diagnostics, cobas, Rotkreuz, Switzerland). |
| Chronological Age | Chronological age was assessed as time since birth. |
| Cytomegalovirus (CMV) | Anti-CMV IgG antibody levels were determined in plasma samples from BASE-II participants using an enzyme immunoassay based CMV IgG kit (Omega Diagnostic Group, Edinburgh, UK). |
| C-reactive protein (CRP) | C-reactive protein was measured via immunoturbidimetry (Roche Diagnostics, cobas, Rotkreuz, Switzerland). |
| Diabetes Complications Severity Index (DCSI) | Young and colleagues (25) published the Diabetes Complications Severity Index (DCSI) as a tool to objectify the long-term effects of diabetes on different organ systems. Specifically, information regarding retinopathy, nephropathy, neuropathy, cerebrovascular disease, cardiovascular disease, peripheral vascular disease, and metabolic complications is aggregated in dependence of the severity (0 =”no abnormality” and 2 =”severe abnormality”) to score the final DCSI-value. Details on how DCSI was calculated in BASE-II, including slight adaptations to the original algorithm, are reported elsewhere (26). |
| Dehydroepiandrosterone sulfate (DHEA-S) | Dehydroepiandrosterone sulfate was measured in a standard hospital laboratory. Towards the end of the study, a change from Radioimmunoassay, TKDS Coat-A-Count kits (Siemens Healthcare Diagnostics Ltd., Germany) to Electrochemiluminescence immunoassays (ECLIA), Elecsys and cobas e (Roche Diagnostics GmbH, Rotkreuz, Switzerland) occurred. However, both methods provided highly comparable results and further information regarding the switch in methods can be found in the Supplementary Table of ref. (27). |
| Digit Symbol Substitution Test (DSST) | In the Digit Symbol Substitution Test (28), participants match symbols to corresponding numbers on a single worksheet using a reference key displayed at the top of the page. Cognitive performance was then assessed by counting the number of correctly completed symbol–number pairings. |
| Education | Education was assessed as the number of years spent in formal schooling. |

| | |
|---|---|
| Electronic Tapping Test (ETT) | Participants were asked to place the hand and arm on the table and tap as fast as they could with their index finger. This test was repeated 5 times. If the difference between tests was larger than 5, the test was repeated until 5 tries had a difference of 5 or lower. The mean of all tries was calculated for each hand, and the higher value was used for subsequent analyses. |
| Eosinophils | See Lymphocytes |
| Episodic Memory | Episodic Memory was assessed as latent factor from a confirmatory factor analysis. Details on how this variable was assessed were previously published (29). |
| Estrogen | Estrogen was measured in a standard hospital laboratory using the same methods described for Total Testosterone. |
| Executive Function and Processing Speed (CERAD-factor) | See Verbal Memory (CERAD-factor). |
| Falls in past 12 months | Participants were asked in one-to-one interviews whether they had fallen within the 12 months preceding the assessment. |
| Fasting Blood Glucose | Fasting blood glucose was measured using Enzymatic reference method with hexokinase (Roche Diagnostics GmbH, Rotkreuz, Switzerland). |
| Ferritin | Ferritin was measured using a particle enhanced immunoturbidimetric assay (Roche Diagnostics, cobas, Rotkreuz, Switzerland). |
| Forced Expiratory Volume in 1 Second (FEV1) | Please see FVC. |
| FEV1/FVC ratio | Please see FVC |
| Fingertips-to-Floor Distance (FFD) | Participants were instructed to bend at the waist and attempt to touch the floor with their fingertips while keeping their legs straight and their feet together. The vertical distance from the fingertips to the floor was recorded in centimeters and served as an indicator of the participants' mobility (30). |
| Fibrinogen | Fibrinogen was measured using a modified version of the method described by Clauss. Citrated plasma is clotted using an excess amount of thrombin, and the resulting clotting time primarily |

| | |
|---|---|
| | reflects the fibrinogen concentration in the sample (Siemens Healthcare, Munich, Germany). |
| Fluid Intelligence | Fluid intelligence was assessed as latent factor from a confirmatory factor analysis. Details on how this variable was assessed were previously published (29). |
| Fried's Frailty Phenotype (FP) | The frailty phenotype according to the definition of Fried and colleagues (31) incorporates information on unintended weight loss, exhaustion, weakness, slow walking speed, and low physical activity. Sex-stratified cut-offs are used to award a point for each of the variables resulting in a possible range of the final index between 0 and 5 points. Further information on how the Fried Frailty Phenotype was assessed in BASE-II was described in detail before (32). |
| Forced Vital Capacity (FVC) | Lung function in BASE-II was assessed using an EasyOne™ spirometer (ndd Medizintechnik AG, Zurich, Switzerland). To ensure high data quality, the assessment was done in accordance with national and international guidelines (e.g. “Deutsche Atemwegsliga” and “Deutsche Gesellschaft für Pneumologie”). Participants were asked to perform the assessment at least twice and the highest value from all tests was used for subsequent analyses. |
| Gamma Globulin | See Alpha-1-Globulin. |
| Gamma-glutamyl Transferase (GGT) | Gamma-glutamyl Transferase was measured using an enzymatic colorimetric assay (Roche Diagnostics, cobas, Rotkreuz, Switzerland). |
| Growth/differentiation factor 15 (GDF15) | Because directly measured GDF15 was not available in BASE-II, DNA methylation–based GDF15 (DNAmGDF15) was derived from Illumina MethylationEPIC array data using the method described by Lu and colleagues (16). |
| Genetic ancestry | Genetic ancestry was assessed as the first four principal components (PC1-PC4) from a principal component analysis (PCA) on genome-wide single nucleotide polymorphism genotyping data (33). |
| Geriatric Depression Scale (GDS) | The Geriatric Depression Scale (GDS) (34) is a screening instrument which was designed to detect depressive symptoms in older people. In this study the 15-question version of the GDS was used. |

| | |
|---|---|
| Glomerular Filtration Rate (GFR) | The Glomerular Filtration Rate was calculated using the sex-specific FAS-formula including creatinine and age. Further information on the GFR in BASE-II was described in detail before (35). |
| Grooved Pegboard Dexterity Test | The Grooved Pegboard Dexterity Test evaluates fine motor skills as well as visual-motor coordination. Participants are asked to put kegs in the correct rotation into holes in board in front of them. The time needed to complete the task (25 pegs) is documented for each hand individually. In this study, the lower value (=better result) was used for subsequent analyses. |
| Hand Grip Strength (HGS) | Hand Grip Strength was measured using a Smedley Dynamometer (Scandidact, Denmark). Participants were asked to stand comfortably with adducted and neutrally rotated shoulders, elbow flexed at 90°, and forearms and wrists in neutral position. Three contractions were performed with each hand, and the maximal Hand Grip Strength was used for further analyses. |
| Hemoglobin A1c (HbA1c) | Hemoglobin A1c was measured using high performance liquid chromatography (HPLC) using the VARIANT II TURBO HbA1c Kit – 2.0 (Bio-Rad Laboratories, Inc., Hercules, CA, USA). |
| High-density lipoprotein (HDL) | High-density lipoprotein was measured via homogeneous enzymatic colorimetric test (Roche Diagnostics, Rotkreuz, Switzerland). |
| Heart Rate Variability (HRV) | Hear Rate Variability was obtained from an 2-hour electrocardiogram (ECG) using the CardioMem® CM 3000-12 BT (Getemed, Teltow, Germany). |
| Homocysteine | Homocysteine was assessed photometrically using an enzyme-cycling assay. |
| Interleukin 6 (IL-6) | IL-6 levels were quantified using BD Biosciences' high-sensitivity Cytometric Bead Array flex kit, with an additional dilution of the standard, measured in triplicate, to enhance the precision of the standard curves. To ensure consistent flow cytometer (BD LSR-II) performance, tracking beads and cytometer settings were regularly monitored. |
| International Normalized Ratio (INR) | The International Normalized Ratio was calculated by comparing a patient's prothrombin time to that of a standard control sample and then raising this ratio to the power of the testing system's ISI value. The prothrombin time was determined via |

| | |
|---|---|
| | electromechanical clotting test (Siemens Healthcare, Munich, Germany). |
| Insulin resistance | Insulin resistance was assessed using the Homeostasis Model Assessment (HOMA) Index. Insulin was measured using electrochemiluminescence immunoassay “ECLIA” (Insulin Elecsys 2010, Roche Diagnostics, cobas, Rotkreuz, Switzerland) |
| Low-density lipoprotein (LDL) | Low-density lipoprotein was measured via homogeneous enzymatic colorimetric test (Roche Diagnostics, Rotkreuz, Switzerland). |
| Leptin | Because directly measured Leptin was not available in BASE-II, DNA methylation–based Leptin (DNAmLeptin) was derived from Illumina MethylationEPIC array data using the method described by Lu and colleagues (16). |
| Lipase | Lipase was measured in a standard hospital laboratory. |
| Lipoprotein(a) (Lp(a)) | Concentration of Lp(a) in blood samples was measured using immunoturbidimetry (Tina-quant lipoprotein[a] assay, LPALX, Roche Diagnostics, Rotkreuz, Switzerland). |
| Life's Simple 7 (LS7) | The Life’s Simple 7 score developed by the American Heart Association (AHA) (36) was slightly adapted to be able to be calculated in BASE-II (37). The score incorporates information on the participant’s BMI, blood pressure, total cholesterol, HbA1c, diet, smoking and physical activity. |
| Lymphocytes | Cell counts were determined using flow cytometry (MVZ Labor 28 GmbH, Berlin, Germany). |
| Magnesium | Magnesium concentration was assessed by photometrically measuring the decrease of xylidyl blue extinction (Roche Diagnostics, Rotkreuz, Switzerland). |
| Metabolic dysfunction–associated steatotic liver disease (MASLD) | Participants were diagnosed with MASLD if their fatty liver index (38) was $\geq 60$, and they met at least one of the five cardiometabolic criteria listed in (39). While the diagnosis should be only given if alcohol consumption was less than 10 g/day for women and less than 20 g/day for men, we calculated and investigated this variable for all participants (with available information on the criteria in question). To assess whether this changes the reported effect sizes, a sensitivity analysis with the subgroup of participants below the cut-off values for alcohol intake was calculated and results are shown for MultiAge cross- |

| | |
|---|---|
| | sectionally (Supplementary Table 28) and longitudinally (Supplementary Table 29). With respect to MultiAge, only small changes in the OR were found cross-sectionally, but longitudinally, a substantially stronger association was observable in the restricted subgroup (OR = 1.4 vs. OR = 1.7, model 3). |
| Metabolic Syndrome (MetS) | The Metabolic Syndrome was diagnosed in BASE-II according to the definition by Alberti and colleagues (40). Therefore, MetS was defined as present if at least three of the following five criteria were fulfilled:<br><br>- *Abdominal obesity:* waist circumference ≥94 cm in men and ≥80 cm in women<br>- *High triglycerides:* triglycerides ≥150 mg/dl<br>- *Low HDL-C:* HDL-C <40 mg/dl in men; <50 mg/dl in women<br>- *High blood pressure:* systolic blood pressure ≥130 and/or diastolic blood pressure ≥85 mmHg or prevalent hypertension or use of antihypertensive medication in participants with known hypertension<br>- *Insulin resistance:* fasting glucose ≥100 mg/dL, prevalent T2D or use of antidiabetic medication in participants with T2D.<br><br>Further information regarding the definition and assessment of Metabolic Syndrome in BASE-II can be found in ref. (41). |
| Mini-Mental State Examination (MMSE) | The Mini-Mental State Examination (MMSE) (42) is a brief, interviewer-administered screening instrument used to assess and monitor cognitive function. It consists of a series of simple questions and tasks completed by the participant which are designed to evaluate several cognitive domains, including orientation, attention, learning ability, calculation, delayed memory recall, and visuoconstructive skills. An unauthorized version of the German MMSE was used by the study team without permission and has been rectified with PAR. The MMSE is a copyrighted instrument and may not be used or reproduced in whole or in part, in any form or language, or by any means without written permission of PAR (www.parinc.com). |
| Mini Nutritional Assessment (MNA) | The Mini Nutritional Assessment (MNA) (43) is an 18-item assessment that aims to identify malnutrition or patients who are at risk for malnutrition. Between 0 and 3 points are given per item which results in maximum of 30 achievable points. Values of 24 |

| | |
|---|---|
| | points or higher indicate that no increased risk for malnutrition was detected. |
| Monocytes | See Lymphocytes |
| Morbidity Index (MI) | In this study, a modified version (44) of the Charlson Morbidity Index (45) first published in 1987 was used. The index was designed to give an overall representation of the individual morbidity burden by incorporating information of the prevalence as well as the severity of the following disease: myocardial infarction, congestive heart failure, peripheral vascular disease, cerebrovascular disease, dementia, chronic pulmonary disease, connective tissue disease, mild liver disease, diabetes, diabetes with end-organ damage, renal disease, hemiplegia, lymphoma, leukemia, any tumor, and moderate to severe liver disease. A detailed description on how the Morbidity Index was calculated in BASE-II was previously published (44). |
| Mortality | Mortality data for BASE-II participants is routinely updated through the Berlin city registry. Mortality data used in this study was last updated in in May 2025. |
| Muscle Mass | Muscle mass, defined as absolute appendicular lean mass (ALM), was determined by summing the lean mass of all four limbs from DEXA data (please see “Bone Mineral Density (BMD)” for additional information). The BMI standardized ALM was used in this study. Further information about muscle mass in BASE-II can be found in ref. (32). |
| Monocytes s | See Lymphocytes |
| Osteocalcin | Osteocalcin was assessed by radioimmunoassay for quantitative determination of intact osteocalcin in serum samples (BRAHMS, Henningsdorf, Germany) |
| Phosphate | Inorganic phosphate reacts with ammonium molybdate to form an ammonium phosphomolybdate complex, which is quantified photometrically in the ultraviolet range (PHOS, Roche Diagnostics GmbH, Rotkreuz, Switzerland). |
| Physical Activity (PA) | Physical activity was measured using the Rapid Assessment of Physical Activity (RAPA) questionnaire developed by Topolski and colleagues (46). The questionnaire consists of seven questions regarding the frequency and intensity of physical |

| | |
|---|---|
| | activity and is scored by identifying the highest ranked affirmed item. |
| Plasminogen activator inhibitor-1 (PAI-1) | Because directly measured Plasminogen activator inhibitor-1 was not available in BASE-II, DNA methylation–based PAI1 (DNAmPAI1) was derived from Illumina MethylationEPIC array data using the method described by Lu and colleagues (16). |
| Potassium | Potassium, sodium, and chloride were measured with an ion-selective electrode in automatically diluted serum/plasm (ISE indirect NA-k_Cl for Gen.2, Roche Diagnostics, Rotkreuz, Switzerland). |
| Pulse wave velocity (PWV) | The pulse wave analysis (PWA) was conducted as described for the “Augmentation Index (AIx)” above. Oscillometric recordings obtained from an upper-arm cuff were used to analyze the brachial artery pressure waveform. Pulse wave velocity (PWV) was calculated from the time interval between the forward pulse wave and its reflected counterpart, using an estimated aortic path length based on body height divided by half the reflection time. Additional information can be found elsewhere (22). |
| Romberg’s Test | The neurological function for balance was tested using Romberg’s test. Participants were asked by the examiner to stand with their feet together and put their hands by their sides. Then, the participant is asked to close their eyes, and the examiner observes the participant for one full minute. Swaying, unsteadiness and the tendency to fall is documented as well as whether the participant is not able to perform the test or if they require constant support. |
| Systematic Coronary Risk Evaluation 2 (SCORE2) | Systematic Coronary Risk Evaluation 2 (SCORE2) (47) and SCORE2-OP (for participants >70 years, (48), hereafter referred to as SCORE2) are assessments designed to estimate the 10-year risk of cardiovascular disease for patients in Europe. Although the original authors recommend excluding participants with diagnosed diabetes mellitus, myocardial infarction, or stroke from SCORE2 calculations, we imputed missing values for these individuals to ensure comparability across outcomes. While this approach deviates from recommendations on how to apply this score in a clinical context, we maintain that SCORE2 remains informative for assessing participants’ cardiovascular health within the scope of this study. |
| Sex | Sex was assessed in one-on-one interviews with study personnel. |

| | |
|---|---|
| Sex hormone-binding globulin (SHBG) | SHBG was determined via electrochemiluminescence immunoassay "ECLIA" (Roche Diagnostics, cobas, Rotkreuz, Switzerland). |
| Smoking (packyears) | Pack-years were assessed by trained study personnel in one-on-one interviews. |
| Sodium | See Potassium |
| SPRINT-BASEed Frailty Index (SPRINT FI) | An adapted version of the frailty index developed in the Systolic Blood Pressure Intervention Trial (SPRINT) by Pajewski and colleagues (49) was calculated in BASE-II to offer an alternative quantification of frailty compared to Fried's Frailty Phenotype. The SPRINT-BASEed frailty index utilizes 31 of the 37 items described in the original publication. In addition, grip strength was included resulting in a total of 32 items. If no information was available for more than 2 items, no index was calculated for the respective participant. More details on how this index was calculated in BASE-II are described in ref. (50). |
| Standard deviation of the NN interval (SDNN) | A 2-hour electrocardiogram (ECG) was conducted using the CardioMem® CM 3000-12 BT (Getemed, Teltow, Germany). The ECG was evaluated using CardioDay software, Version 2.2.1. The standard deviation of all normal-to-normal (NN) intervals within the measurement period, expressed in milliseconds, was calculated from the long-term ECG described under pNN50. |
| Systolic Blood Pressure (SBP) | Blood pressure was measured by study personnel in a seated position on the right and left arm after a resting period of at least 5 minutes. The mean systolic blood pressure measured from the left and right arm was used for further analyses. To reduce variability between study personnel, blood pressure was measured using an electronic sphygmomanometer (boso-medicus memory, Jung Willingen, Germany). |
| Type 2 diabetes (diagnosed) | Type 2 diabetes was diagnosed based on American Diabetes Association (ADA) guidelines(51) if at least one of the following criteria was fulfilled:<br>- Anamnestic history of T2D (self-report)<br>- Antidiabetic medication<br>- Fasting plasma glucose ≥126 mg/dL<br>- 2 h plasma glucose during 75 g- OGTT ≥200 mg/dL<br>- HbA1c ≥ 48 mmol/mol [6.5%] |

| | |
|---|---|
| Timed Test of Money Counting (TTMC) | The Timed Test of Money Counting (TTMC) by Nikolaus and colleagues (52) tests cognitive capacity as well as manual skill by counting the seconds a person's needs to take a specified amount of money out of a purse and count it. |
| Timed Up and Go (TUG) | The Timed Up and Go test was performed according to the original publication (53). The time in seconds it took participants to get up from a standard chair, walk a distance of 3 meters, turn and return to chair and sit down was measured in seconds. |
| Tinetti Test | The Tinetti Test (54) is a standardized clinical tool used to assess fall risk in older adults and consists of two components: balance (part 1) and gait (part 2). The first part includes tasks such as standing without support, turning, and maintaining postural stability in response to a light external push. For the second part, participants were asked to walk and length and height of steps as well as continuity and symmetry of gait was scored by the examiner. |
| Tissue Inhibitor of Matrix Metalloproteinase-1 (TIMP-1) | Because directly measured TIMP1 was not available in BASE-II, DNA methylation–based TIMP1 (DNAmTIMP1) was derived from Illumina MethylationEPIC array data using the method described by Lu and colleagues (16). |
| Tumor necrosis factor (TNF) | Serum concentrations of tumor necrosis factor were quantified using the high-sensitivity CBA Flex system (BD, Franklin Lakes, USA) in accordance with the manufacturer's protocol. To improve the precision of the standard curves, an additional dilution of the standard was included and measured in triplicate. Data acquisition was performed on a BD LSR-II flow cytometer, with instrument performance monitored using BD CS&T beads. |
| Total Cholesterol | Total cholesterol was measured using an enzymatic colorimetric assay (Roche Diagnostics GmbH, Rotkreuz, Switzerland). |
| Total Testosterone | Total Testosterone was measured in a standard hospital laboratory. Samples processed prior to September 2013 were measured with Fluorimmunoassay, AutoDELFIA system (Perkin Elmer Inc., Waltham, MA). Blood samples taken at a later time were analyzed by Electrochemiluminescence immunoassays (ECLIA), Elecsys and cobas e (Roche Diagnostics GmbH, Rotkreuz, Switzerland). |

| | |
|---|---|
| Triglycerides | Triglycerides were measured via homogeneous enzymatic colorimetric test (Triglyceride GPO-PAP, Roche Diagnostics, Rotkreuz, Switzerland). |
| Troponin T | High-sensitivity cardiac troponin T (hs-cTnT) was measured using an immunoassay (Elecsys Troponin Ths, Roche Diagnostics, Rotkreuz Switzerland). |
| Urea | Urea was measured via kinetic UV-test (UREA/BUN, Roche Diagnostics, Rotkreuz, Switzerland). |
| Uric Acid | Uric acid was measured using an enzymatic colorimetric test (Uric Acid ver.2, Roche Diagnostics, cobas, Rotkreuz, Switzerland). |
| Verbal Fluency (CERAD-factor) | See Verbal Memory (CERAD-factor). |
| Verbal Memory (CERAD-factor) | Cognitive performance was assessed in BASE-II using the CERAD-Plus test battery (Consortium to Establish a Registry for Alzheimer's Disease) (55). By applying a confirmatory factor analysis (CFA) on the CERAD-Plus variables, four latent factors representing verbal memory, verbal fluency, executive function and processing speed, visuo-construction were generated. Further details can be found elsewhere (56). |
| Visuo-Construction (CERAD-factor) | See Verbal Memory (CERAD-factor). |
| Vitamin D | Levels of 25-hydroxyvitamin D serum (25(OH)D) were measured in an accredited standard laboratory. A change in methodology occurred in the laboratory during the study period. The following two chemiluminescence immunoassays (CLIA) were used to measure 25(OH)D in BASE-II: IDS-iSYS 25-hydroxyvitamin (IDS-iSYS 25OHD, Immunodiagnostic Systems, UK) and LIAISON 25 OH Vitamin D TOTAL Assay (DiaSorin, Italy). |
| Waist-to-hip ratio (WHR) | Waist and hip circumference were measured in light cloths in centimeters using a non-elastic tape measure by study personnel. |
| Working Memory | Working Memory was assessed as latent factor from a confirmatory factor analysis. Details on how this variable was assessed were previously published (29). |

Continuously scaled variables which were part of the outcome panel to evaluate the biomarkers of aging in BASE-II were dichotomized as described before (57) to be able to analyze them in logistic regression analyses as well as examine incident cases at follow-up.

#### KORA FF4

##### Study population

The KORA platform is a long-running research initiative in southern Germany's Augsburg region (58), featuring multiple baseline surveys: S1 (1984/1985), S2 (1989/1990), S3 (1994/1995), and S4 (1999/2000). As part of the KORA framework, participants undergo different health assessments to observe trends in health conditions, lifestyle factors, and biological indicators at the KORA Study Centre in collaboration with Augsburg University Hospital, Augsburg, Germany. The KORA FF4 study (2013–2014), is the second follow-up of the S4 baseline cohort and participants underwent detailed phenotyping with a focus on cardiometabolic diseases using standardized protocols.

Ethical approval for the KORA S4/F4/FF4 examinations was granted by the local ethics committee, and written informed consent was obtained from all participants (06068).

##### DNA methylation measurement

Genomic DNA (750 ng) from 1920 individuals was bisulfite converted using the EZ-96 DNA Methylation Kit (Zymo Research, Orange, CA, USA) in two batches (N=488, N=1440), with 8 individuals subsequently withdrawing consent for their data to be used (2 from batch 1, 6 from batch 2). Subsequent methylation analysis was performed on an Illumina (San Diego, CA, USA) iScan platform using the Infinium MethylationEPIC BeadChip according to standard protocols provided by Illumina. GenomeStudio software version 2011.1 with Methylation Module version 1.9.0 was used for initial quality control of assay performance and for generation of methylation data export files.

Further quality control and preprocessing of the data were performed in R v3.5.1 with the package minfi v1.28.3 (59) and following primarily the CPACOR pipeline (60). Probes with detection p-values >0.01 were set to missing.

Before normalization, forty samples were removed: 2 showed a mismatch between reported sex and that predicted by minfi; 33 had median intensity <50% of the experiment-wide mean, or <2000 arbitrary units; and 9 (overlap of 4 with previous) had >5% missing values on the autosomes. A total of 59631 probes were removed (some overlapping multiple categories): cross-reactive probes as given in published lists (N=44493) (61, 62); probes with SNPs with minor allele frequency >5% at the CG position (N=11370) or the single base extension (N=5597) as given by minfi; and 5786 with >5% missing values. A total of 806228 probes remained for analysis. Probes from the X chromosome (N=17743, following quality control) and the Y chromosome (N=379) were excluded from the analysis.

Quantile normalization (QN) was then performed separately on the signal intensities. The transformed intensities were then used to generate methylation beta values, a measure from 0 to 1 indicating the percentage of cells methylated at a given locus. As methylation levels in blood can be strongly influenced by leukocyte composition, the white blood cell type proportions were calculated using the method of Houseman et al (63) as implemented in minfi.

#### Variables

##### Type 2 Diabetes

Known type 2 diabetes was defined based on self-report, physician confirmation, or current use of glucose-lowering medication. Participants without a prior diagnosis underwent standardized testing: a 2-hour OGTT and fasting glucose after at least 8 hours of fasting. Diabetes was defined as known T2D or by meeting any of the following criteria: fasting plasma glucose ≥126 mg/dL (≥7.0 mmol/L) or 2-hour OGTT glucose ≥200 mg/dL (≥11.1 mmol/L). Glucose concentrations were determined either the GLU assay or the GLUC3 assay(64).

##### Metabolic Syndrome

Metabolic syndrome (MetS) was defined according to the harmonized criteria proposed by Alberti et al., requiring the presence of at least three of the following five components: (1) waist circumference ≥94 cm in men or ≥80 cm in women; (2) fasting triglycerides ≥150 mg/dL or treatment for elevated triglycerides; (3) HDL cholesterol <40 mg/dL in men or <50 mg/dL in women or treatment affecting HDL (e.g., fibrates); (4) systolic blood pressure ≥130 mmHg or diastolic blood pressure ≥85 mmHg or use of antihypertensive medication; and (5) fasting glucose ≥100 mg/dL or use of antidiabetic medication (65). The MetS score was calculated as the sum of fulfilled components, ranging from 0 to 5.

##### Depressive Symptoms

Mental health depression was assessed by applying the Brief Patient Health Questionnaire (PHQ-D) short form (66), a nine-symptom checklist scoring each depressive symptom from 0 (“not at all“) to 3 (“nearly every day“) leading to a range of 0–27. This variable is categorized by three types: no depressive disorder, mild or subthreshold depressive disorder and marked to severe depressive disorder.

#### KORA AGE

##### Study population

The KORA-Age study tracks a group of adults aged 65 to 93 years at baseline in 2008/09, selected from former KORA S1-S4 participants (67-69). The KORA-Age study focused on aged-related conditions such as multimorbidity, frailty, sarcopenia, mental health including cognition and activities of daily living (69). Written informed consent was obtained from all participants, and ethical approval was granted by the local ethics committee (08064).

##### DNA methylation measurement

Genomic DNA (750 ng) from N=1026 individuals was bisulfite converted using the EZ-96 DNA Methylation Kit (Zymo Research, Orange, CA, USA). Subsequent methylation analysis was performed on an Illumina (San Diego, CA, USA) iScan platform using the Infinium MethylationEPIC BeadChip according to standard protocols provided by Illumina. GenomeStudio software version 2011.1 with Methylation Module version 1.9.0 was used for initial quality control of assay performance and for generation of methylation data export files. Further quality control and preprocessing of the data were performed in R v3.5.1 (https://www.R-project.org/), with the package minfi v1.28.3 (59) and following primarily the CPACOR pipeline (60). Probes with detection p-values >1e-16 were set to missing.

Before normalization, 16 samples were removed: 3 showed a mismatch between reported sex and that predicted by minfi; 1 failed the intensity quality control implemented in minfo (getQC, cut-off 10.5); 7 had >5% missing values on the autosomes; and 5 failed 2 or more of these filters. A total of 70190 probes were removed (some overlapping multiple categories): cross-reactive probes as given in published lists (N=44493) (61, 62); probes with SNPs with minor allele frequency >5% at the CG position (N=11370) or the single base extension (N=5597) as given by minfi; and 18091 with >5% missing values (autosomes only). Probes from the X chromosome (N=17743, following quality control) and the Y chromosome (N=379) were excluded from the analysis.

Quantile normalization (QN) was then performed separately on the signal intensities. The transformed intensities were then used to generate methylation beta values, a measure from 0 to 1 indicating the percentage of cells methylated at a given locus.

As methylation levels in blood can be strongly influenced by leukocyte composition, the white blood cell type proportions were calculated using the method of Houseman et al (63) as implemented in minfi.

##### Variables

###### ADL

Disability was assessed via telephone interview using the Health Assessment Questionnaire Disability Index (HAQ-DI) (73). This instrument comprises 20 items across eight domains (dressing and grooming, hygiene, arising, reach, eating, grip, walking, and usual daily activities), each rated from 0 (no difficulty) to 3 (unable to perform). Domain scores are defined by the highest item score within each domain, and the overall HAQ-DI score is calculated as the mean of the eight domain scores. A score of 0 indicates no disability, whereas a score of 3 reflects severe disability.

##### Cognitive Function

Cognitive status was assessed using the German version of the modified Telephone Interview for Cognitive Status (TICS-m), with adjustment for years of education (74). The instrument was administered according to published, standardized procedures (74, 75) and includes two additional tasks: immediate and delayed verbal recall. The TICS-m comprises the following components: (i) orientation (name, date, age, and phone number; 9 points); (ii) backward counting (2 points); (iii) a 10-word learning task with subsequent delayed recall (20 points); (iv) serial sevens (5 points); (v) responsive naming (4 points); (vi) repetition (2 points); (vii) knowledge of the current German President and Chancellor (4 points); (viii) finger tapping (2 points); and (ix) word opposites (2 points). These items cover four cognitive domains: orientation; memory (registration, recent memory, and delayed recall); attention/calculation; and language (semantic memory, comprehension, and repetition). The total TICS-m score ranges from 0 to 50.

##### Depressive Symptoms

Depressive symptoms were assessed using the 15-item Geriatric Depression Scale (GDS-15) developed by Sheikh and Yesavage (76). This short form provides a reliable indication of depressive symptomatology, even in individuals with mild to moderate cognitive impairment. Scores range from 0 to 15, with 0–5 indicating no depression, 6–10 mild to moderate depression, and 11–15 severe depression.

##### Frailty

Frailty was defined according to the five criteria proposed by Fried et al. (77): (1) weight loss, (2) exhaustion, (3) physical inactivity, (4) low walking speed, and (5) weakness. The criteria were operationalized as follows (78):

- Weight loss: Participants reporting an unintentional weight loss of more than 5 kg in the past six months met this criterion.
- Exhaustion: Participants were asked about their level of energy and activity during the previous two weeks. Those responding “never” to feeling energetic and active were classified as exhausted.
- Physical inactivity: Physical activity was assessed based on sports participation (including cycling) in summer and winter, as well as walking duration on working days. The criterion was met if no sports were performed in at least one season and walking time was less than 30 minutes per working day.
- Low walking speed: Walking speed was measured using the Timed Up and Go (TUG) test (79). Participants in the highest quintile of completion time, stratified by sex and standing height, fulfilled this criterion.

- Weakness: Grip strength was assessed as the mean of three measurements using a JAMAR handheld dynamometer (Saehan Corp., Masan, Korea). Participants in the lowest quintile, stratified by sex and body mass index (BMI), met this criterion.

Participants meeting ≥3 criteria were classified as frail, those with 1–2 criteria as prefrail, and those with none as non-frail. Due to data availability, the applied definitions differ slightly from the original criteria proposed by Fried et al. (77).

##### Multimorbidity

Multimorbidity (MM) was defined as the co-occurrence of two or more chronic conditions within one individual (80). A total of 14 major chronic diseases were considered: hypertension, eye disease, heart disease, diabetes, joint disease, lung disease, gastrointestinal disease, stroke, cancer, kidney disease, liver disease, neurological disease, depression, and anxiety.

Hypertension, diabetes, cancer (diagnosed within the past three years), stroke, and heart disease (myocardial infarction and coronary artery disease) were assessed via self-report, based on whether participants currently had the condition. All other diseases were identified through a telephone interview using items derived from the Charlson Comorbidity Index (81). Participants reported the presence of kidney, liver, and lung diseases (e.g., asthma, chronic bronchitis, emphysema), inflammatory joint conditions (arthritis or rheumatism), gastrointestinal diseases (e.g., colitis, gallbladder, gastric, or ulcer disease), heart conditions (e.g., congestive heart failure or angina), and eye diseases (e.g., cataract, glaucoma, macular degeneration, or diabetic retinopathy). Neurological disorders were assessed by self-report (e.g., epilepsy, Parkinson’s disease, multiple sclerosis).

Depression and anxiety were evaluated using the Geriatric Depression Scale (76) and the Generalized Anxiety Disorder-7 (82), respectively, with scores >10 indicating the presence of depression or anxiety.

##### Type 2 Diabetes

Due to lack of OGTT, diabetes mellitus was defined as a self-reported physician diagnosis, use of anti-diabetic medication, and/or an HbA1c level ≥ 6.5% (48 mmol/mol). Participants were classified as having type 2 diabetes (T2D) if the diagnosis was reported after the age of 25 years (83). Individuals with diabetes other than type 2 were excluded from the analyses.

##### Falls

Socio-demographic characteristics and lifestyle factors were collected through face-to-face interviews. History of falls was assessed using items from the National Health and Nutrition Examination Survey (84). Responses to the question “Did you fall in the previous year?” (“yes, once,” “yes, more than once,” or “no”) were dichotomized into “at least one fall” and “no falls.”

#### SHIP-TREND

##### Study population

The Study of Health in Pomerania (SHIP) is a population-based project conducted in Western Pomerania, a region in Northeast Germany, assessing the prevalence and incidence of common diseases and their risk factors (85). For the current analyses, data from the SHIP-TREND sample, comprising N=4420 participants at baseline (assessed between 2008 and 2012) and N=2507 participants at the first follow-up (assessed between 2016 and 2019), was used. DNA methylation data was available for n=971 participants at baseline. Age, sex, smoking and alcohol habits, physical activities, BMI, current medication, cognitive health and depressive symptoms were documented during a computer-assisted face-to-face interview. Blood samples were extracted on the same day.

##### DNA methylation measurement

DNA was extracted from blood samples of n=508 SHIP-TREND baseline participants to assess DNA methylation using the Illumina HumanMethylationEPIC BeadChip array. Samples were randomly selected based on availability of multiple omics data, excluding type II diabetes, and enriched for prevalent myocardial infarction. The samples were taken between 07:00 AM and 04:00 PM, and serum aliquots were prepared for immediate analysis and for storage at -80 °C in the Integrated Research Biobank (Liconic, Liechtenstein). Processing of the DNA samples was performed at the Helmholtz Zentrum München. Preparation and normalization of the array data was performed according to the CPACOR workflow (60) using the software package R (www.r-project.org). The array idat files were processed using the minfi package. Probes that had a detection p-value above background (sum of per-array methylated and unmethylated intensity values based p-value ≥1E-16) were set to missing. Methylation beta values were calculated as proportion of methylated intensity value on the sum of methylated+unmethylated+100 intensities. Arrays with observed technical problems (±4SD outside control probe intensity mean) during steps like bisulfite conversion, hybridization or extension, as well as arrays with mismatch between sex of the proband and sex determined by the chr X and Y probe intensities were removed from subsequent analyses. Additionally, only arrays with a call rate ≥ 95% were processed further resulting in 495 samples with methylation data on 865,859 sites available for subsequent analyses.

DNA methylation of additional 480 samples of the SHIP-TREND baseline cohort was assessed using the HumanMethylationEPICv2 BeadChip array and processed using the same workflow as before, resulting in 476 samples which passed final quality control. For this array type, the call rate of the final samples was >85%. Technical replicate probes were merged using the *sesame* R package.

To account for potential confounding effects due to blood cell composition, blood cell subtypes were estimated using the method of Houseman et al. (63).

CpG sites with missing values in more than 20% of the samples were excluded from the dataset. Subsequently, missing values were imputed as the mean beta of all available methylation betas for this CpG site. Lastly, participants with missing covariates were excluded, leaving N=958 participants at baseline and N=471 at follow-up that could be included in the present analyses.

### Variables

#### Metabolic Syndrome (MetS)

MetS was defined as present if at least three of the following criteria were fulfilled:

- waist circumference ≥94cm in men and ≥80cm in women
- current antidiabetic therapy (ATC A10) or serum glucose level ≥8mmol/ l (144mg/dl) for participants who fasted for <8h and ≥6.1 mmol/ l (110mg/dl) for participants who fasted for ≥8h
- HDL-C <1.03mmol/ l (<40mg/dl) in men and <1.29mmol/ l (<50mg/dl) in women for participants who fasted for <8h + HDL-C <1.03mmol/ l (<40mg/dl) in men and <1.3mmol/ l (<50mg/dl) in women for participants who fasted for ≥8h
- current lipid-lowering therapy (Fibrate (ATC C10AB) or Niacine (ATC C10AD)) or triglycerides ≥2.3mmol /l (>204mg/ dl) for participants who fasted for <8h and ≥1.7mmol /l (>150mg/ dl) for participants who fasted for ≥8h
- blood pressure ≥130/85mmHg or current antihypertensive therapy (ATC C02A*, C03C*, C03E*, C09BA*, C07A*, C08C*, C08DA*, C09AA*, C09CA*)

#### Cognitive Function

Cognitive health has been measured with the Nuremberg age inventory (NAI). In the test for immediate word recall (NAI short), a list of eight words was read to the participant who afterwards had to repeat all words that he/she remembered from the list. The „NAI short“ score is the number of words that were correctly recalled, i.e. a large score represents good cognitive health. After 20min, a list of 16 words was presented to the participant, eight of which were from the list before and eight were not. The participant had to decide whether each word had been in the list or not. In this test of delayed word recognition (NAI long), the score is the number of correctly recognized words minus the number of words that were recognized although they had not been in the list, i.e. a large score represents good cognitive health.

#### Depressive Symptoms

Depressive symptoms within the last two weeks have been assessed with the patient health questionnaire 9 (PHQ-9). It consists of nine questions addressing aspects such as mood, sleep, appetite, concentration and suicidal thoughts, each assessed on a scale from zero to three. The PHQ-9 score is the sum of all nine single items, i.e. a small score reflects little depressive symptomatic.

#### Mortality

The mortality data investigated in this study was last updated in January 2023. Age at death was calculated by adding the time to death to the age at baseline assessment.

#### BiDirect

##### Study population

The BiDirect Study, conducted in Münster, Germany, investigates the bidirectional relationship between depression and subclinical arteriosclerosis. BiDirect recruited three distinct cohorts, i) patients with acute clinically diagnosed depression (n=1004), ii) patients with established cardiovascular disease (CVD) (n=348), and iii) a control sample of randomly drawn individuals from the city registry of Münster, Germany (n=966). All participants were 35 to 65 years at baseline and examined with an identical, complex program four times over 10 years. The present analysis included a subsample of 557 participants from the baseline population-based cohort, for which DNA methylation data was available.

##### DNA methylation measurement

DNAm levels were measured from whole blood samples in two batches using the Infinium MethylationEPIC v1.0 or v2.0 BeadChip (Illumina Inc. San Diego, USA) arrays. The raw intensity data was generated at Life&Brain GmbH (Bonn, Germany) and pre-processed in-house. Quality control and normalization procedures were performed using the minfi (59) and bigmelon (86) R packages (R Foundation for Statistical Computing, Vienna, Austria, 2018. URL https://www.R-project.org/). These steps included: 1) data conversion, 2) calculation of detection p-values, 3) removal of probes that a) failed detection, b) located on the sex chromosomes, c) contained polymorphisms, and/or d) are flagged as cross-reactive, as well as 4) the removal of outlier samples (outlyx function of bigmelon), 5) calculation of normalized beta values using the dasen method (87) and 6) transformation of normalized betas to M-values for use in statistical analyses.

##### Variables

Computer assisted face-to-face interviews with all BiDirect participants were carried out by trained and certified study nurses. Information about sociodemographic factors, comorbidities including diabetes, cardiovascular and renal diseases, stroke, hypertension as well as lifestyle and risk factors was collected during the interviews.

##### Cognitive Function

A neuropsychological test battery was assessed by trained study nurses from each participant in every examination wave. It included the animal-naming-test, a 12-item emotional word memory list, a Color–Word Interference Test (CWIT), the Trail-Making-Test (TMT) A+B and the Purdue Pegboard Test. From the results of the test battery a global cognition score based on principal component analysis of the z-standardized test scores (using the general population cohort as the reference), was calculated (details are described in Bonberg N. et al: doi: 10.3389/fnagi.2022.804842).

##### Depressive Symptoms

The 20-item Center for Epidemiologic Studies Depression Scale (CES-D) was used in each examination wave in addition to the M.I.N.I. vs.5.0.0 Neuropsychiatric Interview (German version) modul ‘Major Depression’. The established cut-off of $\geq$16 points was used for the CES-D to categorize moderate to severe current depressive symptom severity.

#### References:

1. Bertram L, Bockenhoff A, Demuth I, Duzel S, Eckardt R, Li SC, et al. Cohort profile: The Berlin Aging Study II (BASE-II). International journal of epidemiology. 2014;43(3):703-12.
2. Demuth I, Banszerus V, Drewelies J, Düzel S, Seeland U, Spira D, et al. Cohort profile: follow-up of a Berlin Aging Study II (BASE-II) subsample as part of the GendAge study. BMJ Open. 2021;11(6):e045576.
3. Vetter VM, Drewelies J, Homann J, Düzel S, Deecke L, Jawinski P, et al. Comprehensive Comparison of Sixteen Markers of Biological Aging: Cross-Sectional and Longitudinal Results from the Berlin Aging Study II (BASE-II). medRxiv. 2025:2025.04.09.25325514.
4. Tierney N. visdat: Visualising whole data frames. The Journal of Open Source Software. 2017;2(16):355.
5. van Buuren S, Groothuis-Oudshoorn K. mice: Multivariate Imputation by Chained Equations in R. Journal of Statistical Software. 2011;45(3):1 - 67.
6. Klemera P, Doubal S. A new approach to the concept and computation of biological age. Mechanisms of Ageing and Development. 2006;127(3):240-8.
7. Hochschild R. Improving the precision of biological age determinations. Part 2: automatic human tests, age norms and variability. Experimental gerontology. 1989;24(4):301-16.
8. Kwon D, Belsky DW. A toolkit for quantification of biological age from blood chemistry and organ function test data: BioAge. Geroscience. 2021;43(6):2795-808.
9. Bafei SEC, Shen C. Biomarkers selection and mathematical modeling in biological age estimation. npj Aging. 2023;9(1):13.
10. Dubina TL, Mints AY, Zhuk EV. Biological age and its estimation. III. Introduction of a correction to the multiple regression model of biological age and assessment of biological age in cross-sectional and longitudinal studies. Experimental Gerontology. 1984;19(2):133-43.
11. Nakamura E, Miyao K, Ozeki T. Assessment of biological age by principal component analysis. Mechanisms of Ageing and Development. 1988;46(1):1-18.
12. Jia L, Zhang W, Jia R, Zhang H, Chen X. Construction formula of biological age using the principal component analysis. BioMed research international. 2016;2016(1):4697017.
13. Belsky DW, Caspi A, Corcoran DL, Sugden K, Poulton R, Arseneault L, et al. DunedinPACE, a DNA methylation biomarker of the pace of aging. eLife. 2022;11:e73420.
14. Horvath S. DNA methylation age of human tissues and cell types. Genome biology. 2013;14(10):R115.
15. Mahoney FI, Barthel DW. FUNCTIONAL EVALUATION: THE BARTHEL INDEX. Md State Med J. 1965;14:61-5.
16. Lu AT, Quach A, Wilson JG, Reiner AP, Aviv A, Raj K, et al. DNA methylation GrimAge strongly predicts lifespan and healthspan. Aging (Albany NY). 2019;11(2):303.
17. Nöthlings U, Hoffmann K, Bergmann MM, Boeing H. Fitting portion sizes in a self-administered food frequency questionnaire. The Journal of nutrition. 2007;137(12):2781-6.
18. Seeman TE, Singer BH, Rowe JW, Horwitz RI, McEwen BS. Price of adaptation—allostatic load and its health consequences: MacArthur studies of successful aging. Archives of internal medicine. 1997;157(19):2259-68.
19. Juster R-P, McEwen BS, Lupien SJ. Allostatic load biomarkers of chronic stress and impact on health and cognition. Neuroscience & Biobehavioral Reviews. 2010;35(1):2-16.

20. Seeman M, Merkin SS, Karlamangla A, Koretz B, Seeman T. Social status and biological dysregulation: The "status syndrome" and allostatic load. Social Science & Medicine. 2014;118:143-51.
21. McCrory C, Fiorito G, Cheallaigh CN, Polidoro S, Karisola P, Alenius H, et al. How does socio-economic position (SEP) get biologically embedded? A comparison of allostatic load and the epigenetic clock (s). Psychoneuroendocrinology. 2019;104:64-73.
22. Seeland U, Demuth I, Regitz-Zagrosek V, Steinhagen-Thiessen E, König M. Sex differences in arterial wave reflection and the role of exogenous and endogenous sex hormones: results of the Berlin Aging Study II. Journal of hypertension. 2020;38(6):1040-6.
23. Drewelies J, Hueluer G, Duezel S, Vetter VM, Pawelec G, Steinhagen-Thiessen E, et al. Using blood test parameters to define biological age among older adults: association with morbidity and mortality independent of chronological age validated in two separate birth cohorts. GeroScience. 2022.
24. Radloff LS. The CES-D Scale:A Self-Report Depression Scale for Research in the General Population. Applied Psychological Measurement. 1977;1(3):385-401.
25. Young BA, Lin E, Von Korff M, Simon G, Ciechanowski P, Ludman EJ, et al. Diabetes complications severity index and risk of mortality, hospitalization, and healthcare utilization. The American journal of managed care. 2008;14(1):15.
26. Spieker J, Vetter VM, Spira D, Steinhagen-Thiessen E, Regitz-Zagrosek V, Buchmann N, et al. Diabetes Type 2 in the Berlin Aging Study II: Prevalence, Incidence and Severity Over up to Ten Years of Follow-up. PREPRINT (Version 3) available at Research Square. 2021.
27. Schmid H, Vetter VM, Homann J, Bahr V, Lill CM, Regitz-Zagrosek V, et al. Cross-sectional and Longitudinal Relationship Between Sex Hormones and 6 Epigenetic Clocks in Older Adults: Results of the Berlin Aging Study II. The Journals of Gerontology, Series A: Biological Sciences and Medical Sciences. 2025;80(7):glaf106.
28. San Antonio TPC. Wechsler Adult Intelligence Scale - Revised. 1981.
29. Düzel S, Voelkle MC, Düzel E, Gerstorf D, Drewelies J, Steinhagen-Thiessen E, et al. The Subjective Health Horizon Questionnaire (SHH-Q): assessing future time perspectives for facets of an active lifestyle. Gerontology. 2016;62(3):345-53.
30. Perret C, Poiraudeau S, Fermanian J, Colau MM, Benhamou MA, Revel M. Validity, reliability, and responsiveness of the fingertip-to-floor test. Arch Phys Med Rehabil. 2001;82(11):1566-70.
31. Fried LP, Tangen CM, Walston J, Newman AB, Hirsch C, Gottdiener J, et al. Frailty in older adults: evidence for a phenotype. The Journals of Gerontology Series A: Biological Sciences and Medical Sciences. 2001;56(3):M146-M57.
32. Spira D, Buchmann N, König M, Rosada A, Steinhagen-Thiessen E, Demuth I, et al. Sex-specific differences in the association of vitamin D with low lean mass and frailty–Results from the Berlin Aging Study II. Nutrition. 2018.
33. Hong S, Dobricic V, Ohlei O, Bos I, Vos SJB, Prokopenko D, et al. TMEM106B and CPOX are genetic determinants of cerebrospinal fluid Alzheimer's disease biomarker levels. Alzheimers Dement. 2021;17(10):1628-40.
34. Yesavage JA, Brink TL, Rose TL, Lum O, Huang V, Adey M, et al. Development and validation of a geriatric depression screening scale: a preliminary report. J Psychiatr Res. 1982;17(1):37-49.
35. König M, Gollasch M, Demuth I, Steinhagen-Thiessen E. Prevalence of Impaired Kidney Function in the German Elderly: Results from the Berlin Aging Study II (BASE-II). Gerontology. 2017;63(3):201-9.
36. Lloyd-Jones DM, Hong Y, Labarthe D, Mozaffarian D, Appel LJ, Van Horn L, et al. Defining and setting national goals for cardiovascular health promotion and disease reduction:

the American Heart Association's strategic Impact Goal through 2020 and beyond. Circulation. 2010;121(4):586-613.
37. König M, Drewelies J, Norman K, Spira D, Buchmann N, Hülür G, et al. Historical trends in modifiable indicators of cardiovascular health and self-rated health among older adults: Cohort differences over 20 years between the Berlin Aging Study (BASE) and the Berlin Aging Study II (BASE-II). PLoS one. 2018;13(1):e0191699.
38. Bedogni G, Bellentani S, Miglioli L, Masutti F, Passalacqua M, Castiglione A, et al. The Fatty Liver Index: a simple and accurate predictor of hepatic steatosis in the general population. BMC Gastroenterol. 2006;6:33.
39. Rinella ME, Lazarus JV, Ratziu V, Francque SM, Sanyal AJ, Kanwal F, et al. A multisociety Delphi consensus statement on new fatty liver disease nomenclature. Hepatology. 2023;78(6):1966-86.
40. Alberti KG, Eckel RH, Grundy SM, Zimmet PZ, Cleeman JI, Donato KA, et al. Harmonizing the metabolic syndrome: a joint interim statement of the International Diabetes Federation Task Force on Epidemiology and Prevention; National Heart, Lung, and Blood Institute; American Heart Association; World Heart Federation; International Atherosclerosis Society; and International Association for the Study of Obesity. Circulation. 2009;120(16):1640-5.
41. Demuth I, Vetter VM, Homann J, Junge MP, Regitz-Zagrosek V, Gerstorf D, et al. DunedinPACE Predicts Incident Metabolic Syndrome: Cross-sectional and Longitudinal Data from the Berlin Aging Study II (BASE-II). The Journals of Gerontology: Series A. 2025;80(9).
42. Folstein MF, Folstein SE, McHugh PR. "Mini-mental state". A practical method for grading the cognitive state of patients for the clinician. J Psychiatr Res. 1975;12(3):189-98.
43. Vellas B, Guigoz Y, Garry PJ, Nourhashemi F, Bennahum D, Lauque S, et al. The Mini Nutritional Assessment (MNA) and its use in grading the nutritional state of elderly patients. Nutrition. 1999;15(2):116-22.
44. Meyer A, Salewsky B, Spira D, Steinhagen-Thiessen E, Norman K, Demuth I. Leukocyte telomere length is related to appendicular lean mass: cross-sectional data from the Berlin Aging Study II (BASE-II). Am J Clin Nutr. 2016;103(1):178-83.
45. Charlson ME, Pompei P, Ales KL, MacKenzie CR. A new method of classifying prognostic comorbidity in longitudinal studies: development and validation. J Chronic Dis. 1987;40(5):373-83.
46. Topolski TD, LoGerfo J, Patrick DL, Williams B, Walwick J, Patrick MB. The Rapid Assessment of Physical Activity (RAPA) among older adults. Preventing chronic disease. 2006;3(4):A118-A.
47. Collaboration ECR, group Sw. SCORE2 risk prediction algorithms: new models to estimate 10-year risk of cardiovascular disease in Europe. European Heart Journal. 2021;42(25):2439-54.
48. SCORE2-OP risk prediction algorithms: estimating incident cardiovascular event risk in older persons in four geographical risk regions. European Heart Journal. 2021;42(25):2455-67.
49. Pajewski NM, Williamson JD, Applegate WB, Berlowitz DR, Bolin LP, Chertow GM, et al. Characterizing Frailty Status in the Systolic Blood Pressure Intervention Trial. The Journals of Gerontology: Series A. 2016;71(5):649-55.
50. Vetter VM, Drewelies J, Düzel S, Homann J, Meyer-Arndt L, Braun J, et al. Change in body weight of older adults before and during the COVID-19 pandemic: Longitudinal results from the Berlin Aging Study II. The Journal of nutrition, health and aging. 2024;28(4):100206.
51. Association AD. 2. Classification and diagnosis of diabetes: standards of medical care in diabetes—2019. Diabetes care. 2019;42(Supplement 1):S13-S28.

52. NIKOLAUS T, BACH M, SPECHT-LEIBLE N, OSTER P, SCHLIERF G. The Timed Test of Money Counting: A Short Physical Performance Test for Manual Dexterity and Cognitive Capacity. Age and Ageing. 1995;24(3):257-8.
53. Podsiadlo D, Richardson S. The timed "Up & Go": a test of basic functional mobility for frail elderly persons. Journal of the American Geriatrics Society. 1991;39(2):142-8.
54. Tinetti ME, Williams TF, Mayewski R. Fall risk index for elderly patients based on number of chronic disabilities. The American journal of medicine. 1986;80(3):429-34.
55. Morris JC, Heyman A, Mohs RC, Hughes JP, van Belle G, Fillenbaum G, et al. The Consortium to Establish a Registry for Alzheimer's Disease (CERAD). Part I. Clinical and neuropsychological assessment of Alzheimer's disease. Neurology. 1989;39(9):1159-65.
56. Röhr F, Bucholtz N, Toepfer S, Norman K, Spira D, Steinhagen-Thiessen E, et al. Relationship between Lipoprotein (a) and cognitive function–Results from the Berlin Aging Study ii. Scientific Reports. 2020;10(1):10636.
57. Vetter VM, Drewelies J, Homann J, Düzel S, Deecke L, Jawinski P, et al. Comprehensive cross-sectional and longitudinal comparison of sixteen markers of biological aging from the Berlin Aging Study II. Communications Medicine. 2026;6(1):168.
58. Linkohr B, Heier M, Gieger C, Thorand B, Grallert H, Holle R, et al. Cohort Profile: Cooperative Health Research in the Region of Augsburg (KORA) 1984-2024. Int J Epidemiol. 2025;54(6).
59. Aryee MJ, Jaffe AE, Corrada-Bravo H, Ladd-Acosta C, Feinberg AP, Hansen KD, et al. Minfi: a flexible and comprehensive Bioconductor package for the analysis of Infinium DNA methylation microarrays. Bioinformatics. 2014;30(10):1363-9.
60. Lehne B, Drong AW, Loh M, Zhang W, Scott WR, Tan ST, et al. A coherent approach for analysis of the Illumina HumanMethylation450 BeadChip improves data quality and performance in epigenome-wide association studies. Genome Biol. 2015;16(1):37.
61. McCartney DL, Walker RM, Morris SW, McIntosh AM, Porteous DJ, Evans KL. Identification of polymorphic and off-target probe binding sites on the Illumina Infinium MethylationEPIC BeadChip. Genom Data. 2016;9:22-4.
62. Pidsley R, Zotenko E, Peters TJ, Lawrence MG, Risbridger GP, Molloy P, et al. Critical evaluation of the Illumina MethylationEPIC BeadChip microarray for whole-genome DNA methylation profiling. Genome Biol. 2016;17(1):208.
63. Houseman EA, Accomando WP, Koestler DC, Christensen BC, Marsit CJ, Nelson HH, et al. DNA methylation arrays as surrogate measures of cell mixture distribution. BMC Bioinformatics. 2012;13:86.
64. Laxy M, Knoll G, Schunk M, Meisinger C, Huth C, Holle R. Quality of Diabetes Care in Germany Improved from 2000 to 2007 to 2014, but Improvements Diminished since 2007. Evidence from the Population-Based KORA Studies. PLoS One. 2016;11(10):e0164704.
65. Alberti KG, Eckel RH, Grundy SM, Zimmet PZ, Cleeman JI, Donato KA, et al. Harmonizing the metabolic syndrome: a joint interim statement of the International Diabetes Federation Task Force on Epidemiology and Prevention; National Heart, Lung, and Blood Institute; American Heart Association; World Heart Federation; International Atherosclerosis Society; and International Association for the Study of Obesity. Circulation. 2009;120(16):1640-5.
66. Spitzer RL, Kroenke K, Williams JB. Validation and utility of a self-report version of PRIME-MD: the PHQ primary care study. Primary Care Evaluation of Mental Disorders. Patient Health Questionnaire. Jama. 1999;282(18):1737-44.
67. Holle R, Happich M, Löwel H, Wichmann HE. KORA--a research platform for population based health research. Gesundheitswesen. 2005;67 Suppl 1:S19-25.
68. Steinbeisser K, Grill E, Holle R, Peters A, Seidl H. Determinants for utilization and transitions of long-term care in adults 65+ in Germany: results from the longitudinal KORA-Age study. BMC Geriatr. 2018;18(1):172.

69. Peters A, Döring A, Ladwig KH, Meisinger C, Linkohr B, Autenrieth C, et al. [Multimorbidity and successful aging: the population-based KORA-Age study]. Z Gerontol Geriatr. 2011;44 Suppl 2:41-54.
70. Holle R, Happich M, Löwel H, Wichmann H-E, Group MKS. KORA-a research platform for population based health research. Das Gesundheitswesen. 2005;67(S 01):19-25.
71. Peters A, Döring A, Ladwig K, Meisinger C, Linkohr B, Autenrieth C, et al. Multimorbidity and successful aging: the population-based KORA-Age study. Zeitschrift fur Gerontologie und Geriatrie. 2011;44:41-54.
72. Steinbeisser K, Grill E, Holle R, Peters A, Seidl H. Determinants for utilization and transitions of long-term care in adults 65+ in Germany: results from the longitudinal KORA-Age study. BMC geriatrics. 2018;18(1):172.
73. Fries JF, Spitz PW, Young DY. The dimensions of health outcomes: the health assessment questionnaire, disability and pain scales. J Rheumatol. 1982;9(5):789-93.
74. Lacruz M, Emeny R, Bickel H, Linkohr B, Ladwig K. Feasibility, internal consistency and covariates of TICS-m (telephone interview for cognitive status-modified) in a population-based sample: findings from the KORA-Age study. Int J Geriatr Psychiatry. 2013;28(9):971-8.
75. Plassman BL, Newman TT, Welsh KA, Helms M, Breitner J. Application in epidemiological and longitudinal studies. Neuropsychiatry, Neuropsychology & Behavioral Neurology. 1994;7(3):235-41.
76. Sheikh JI, Yesavage JA. A knowledge assessment test for geriatric psychiatry. Hosp Community Psychiatry. 1985;36(11):1160-6.
77. Fried LP, Tangen CM, Walston J, Newman AB, Hirsch C, Gottdiener J, et al. Frailty in older adults: evidence for a phenotype. J Gerontol A Biol Sci Med Sci. 2001;56(3):M146-56.
78. Johar H, Emeny RT, Bidlingmaier M, Reincke M, Thorand B, Peters A, et al. Blunted diurnal cortisol pattern is associated with frailty: a cross-sectional study of 745 participants aged 65 to 90 years. J Clin Endocrinol Metab. 2014;99(3):E464-8.
79. Podsiadlo D, Richardson S. The timed "Up & Go": a test of basic functional mobility for frail elderly persons. J Am Geriatr Soc. 1991;39(2):142-8.
80. Kirchberger I, Meisinger C, Heier M, Zimmermann AK, Thorand B, Autenrieth CS, et al. Patterns of multimorbidity in the aged population. Results from the KORA-Age study. PLoS One. 2012;7(1):e30556.
81. Chaudhry S, Jin L, Meltzer D. Use of a self-report-generated Charlson Comorbidity Index for predicting mortality. Med Care. 2005;43(6):607-15.
82. Spitzer RL, Kroenke K, Williams JB, Löwe B. A brief measure for assessing generalized anxiety disorder: the GAD-7. Arch Intern Med. 2006;166(10):1092-7.
83. Ferrari U, Then C, Rottenkolber M, Selte C, Seissler J, Conzade R, et al. Longitudinal association of type 2 diabetes and insulin therapy with muscle parameters in the KORA-Age study. Acta Diabetol. 2020;57(9):1057-63.
84. USNCfH S. National Health and Nutrition Examination Survey Questionnaire. Hyattsville: National Center for Health Statistics.
85. Völzke H, Schössow J, Schmidt CO, Jürgens C, Richter A, Werner A, et al. Cohort Profile Update: The Study of Health in Pomerania (SHIP). Int J Epidemiol. 2022;51(6):e372-e83.
86. Gorrie-Stone TJ, Smart MC, Saffari A, Malki K, Hannon E, Burrage J, et al. Bigmelon: tools for analysing large DNA methylation datasets. Bioinformatics. 2019;35(6):981-6.
87. Pidsley R, CC YW, Volta M, Lunnon K, Mill J, Schalkwyk LC. A data-driven approach to preprocessing Illumina 450K methylation array data. BMC Genomics. 2013;14:293.
