## Supplementary figures and images for "MultiAge: A New Multidimensional Biomarker of Biological Age Derived from Comprehensive Phenotypic and Molecular Profiling"

### Supplementary Figure 1

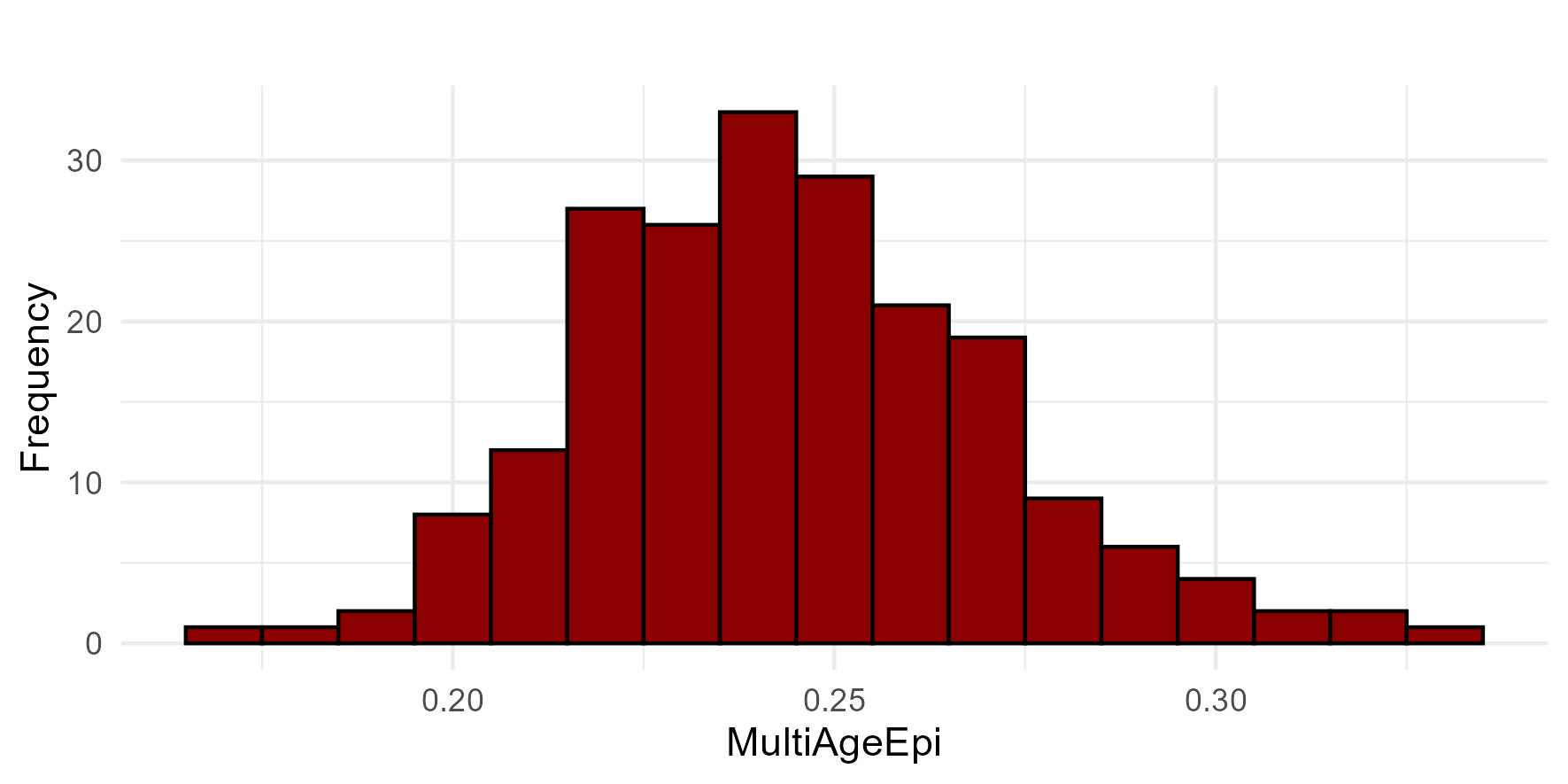

### Supplementary Figure 3

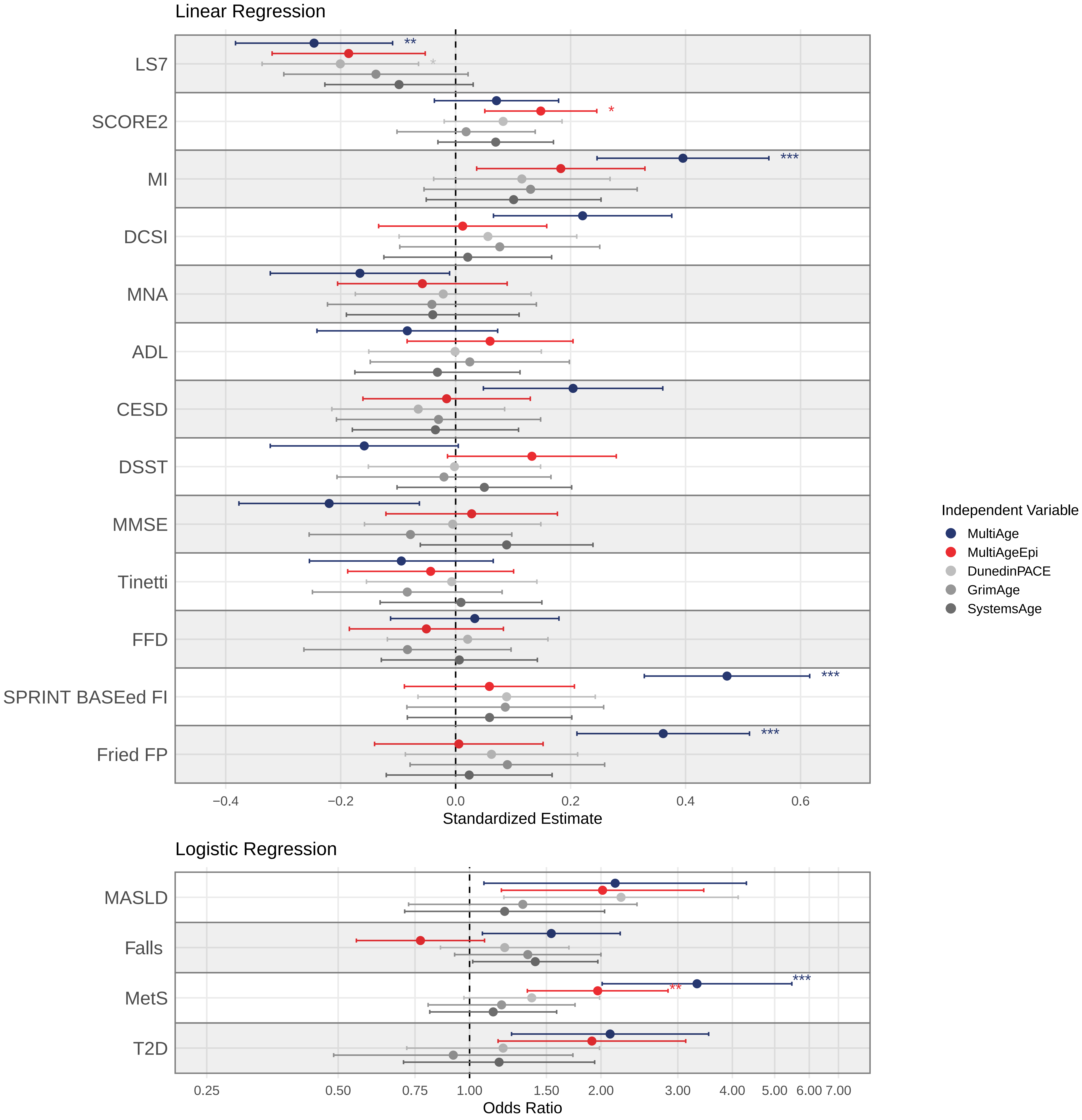

### Supplementary Figure 5

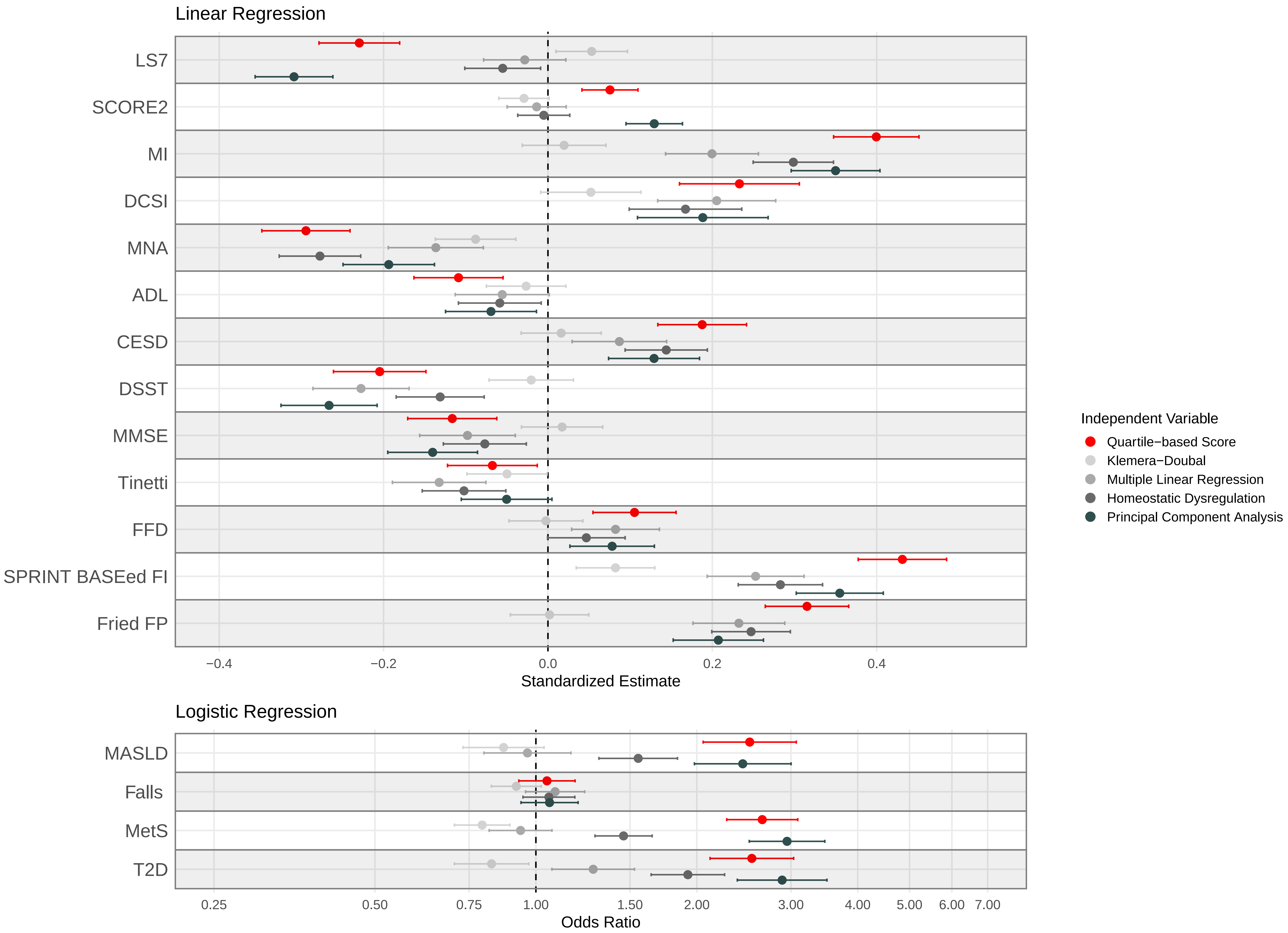

### Supplementary Figure 6

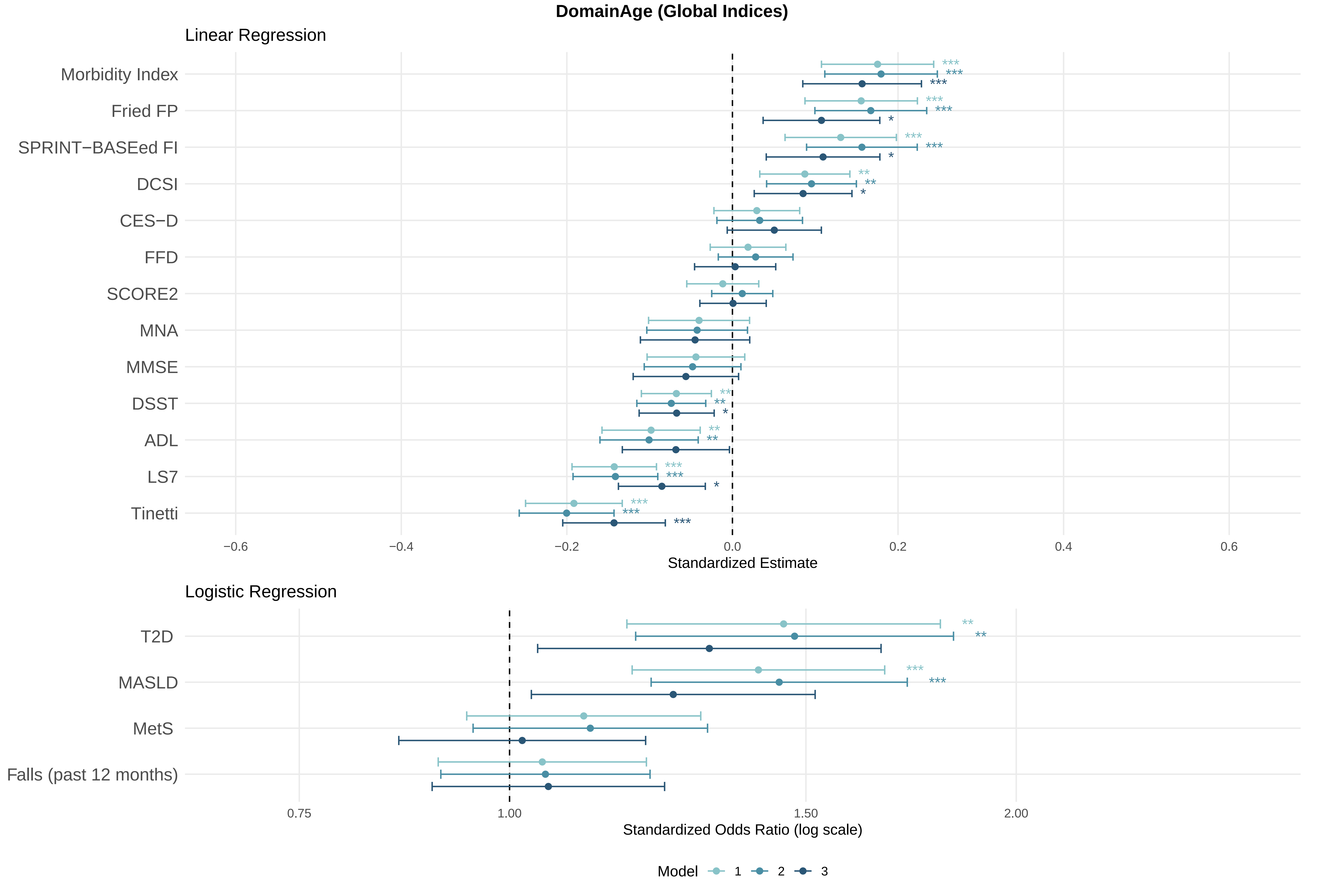

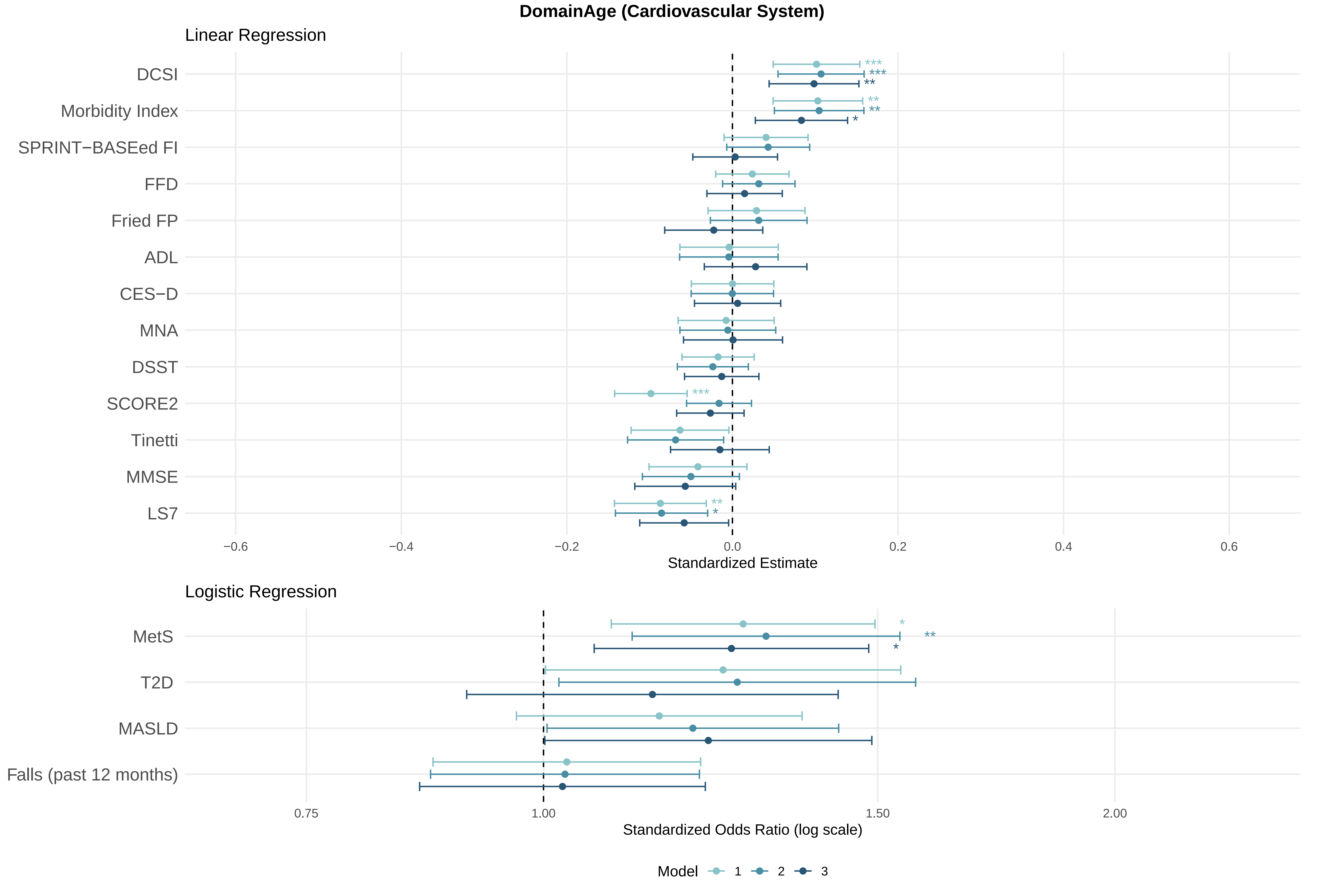

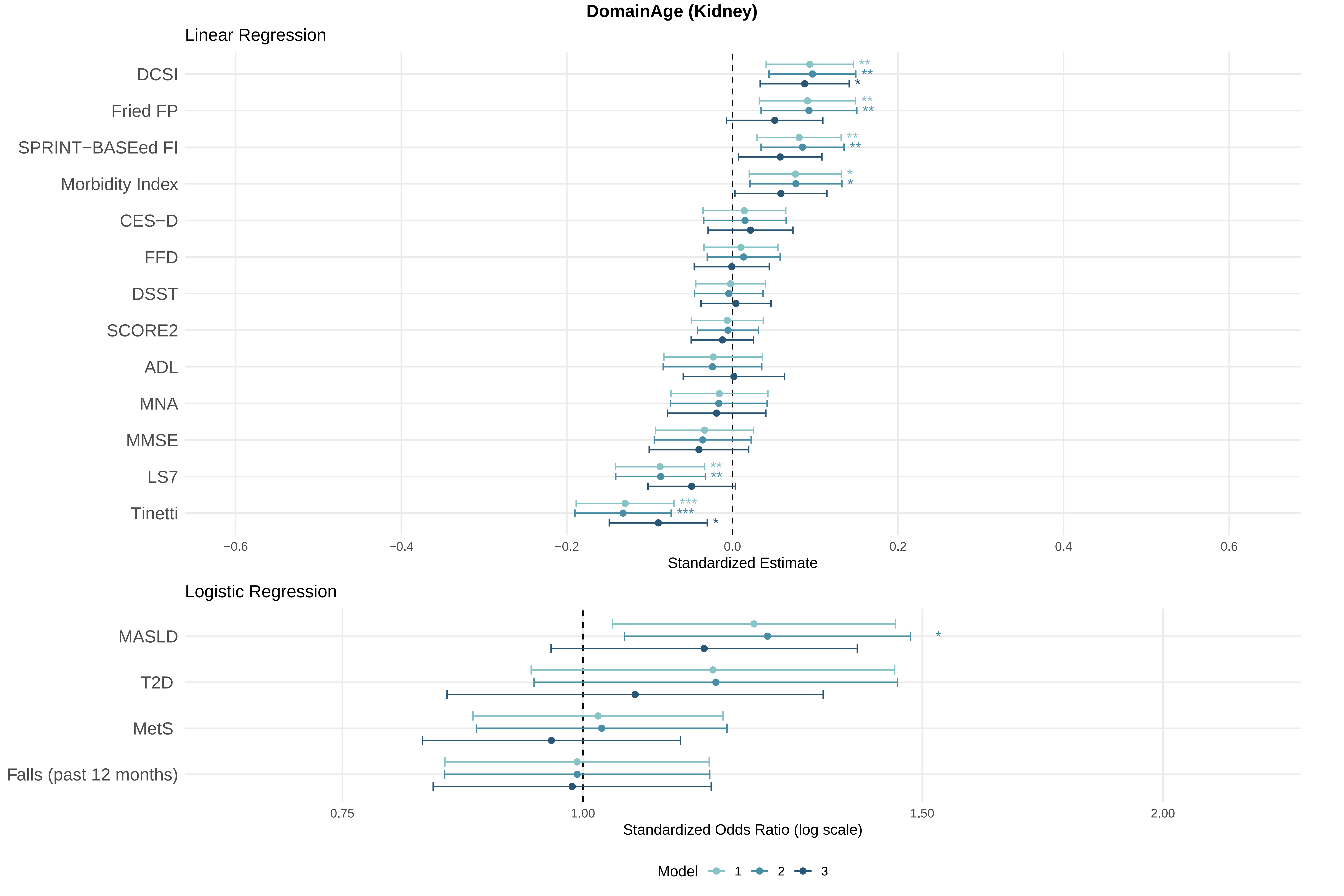

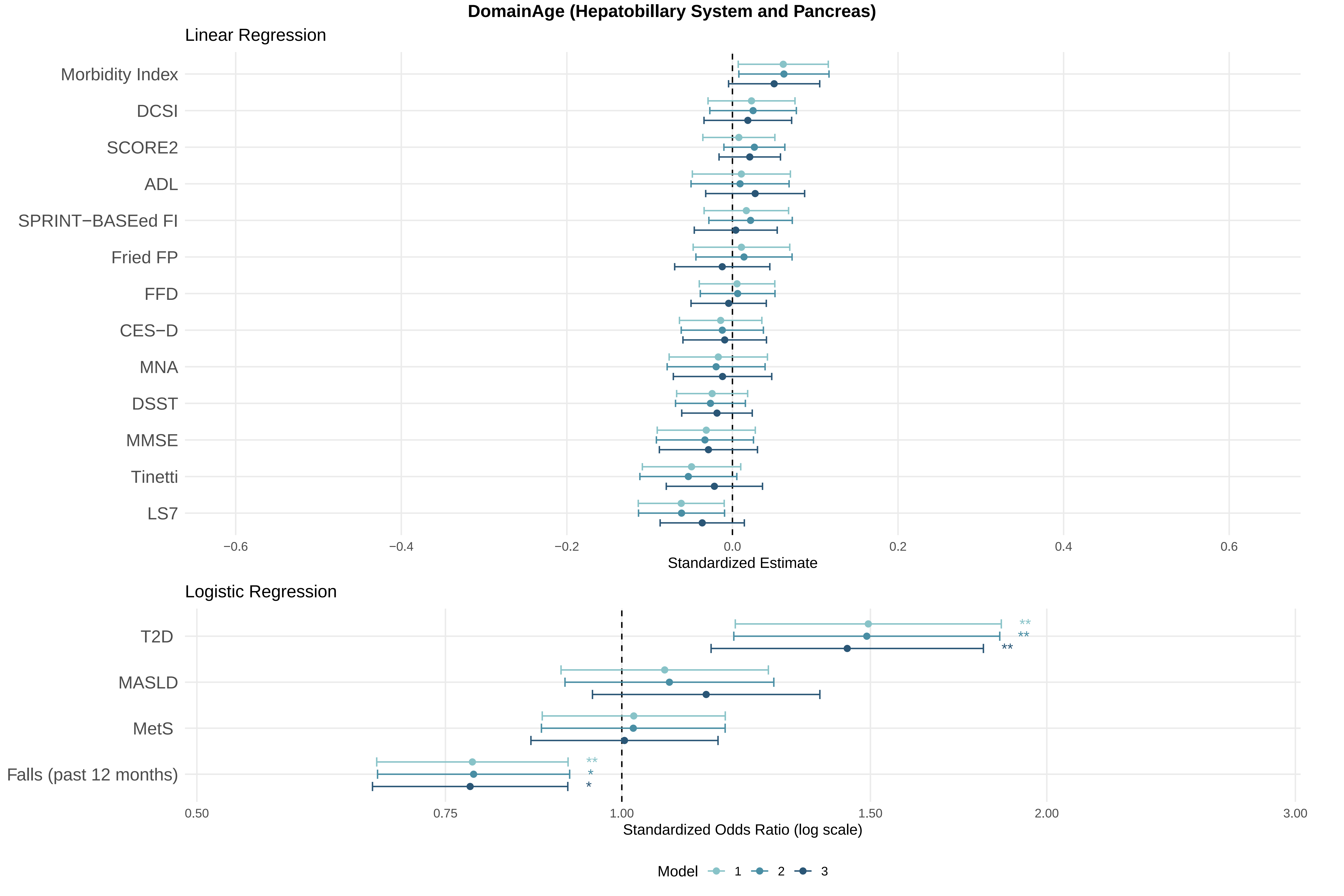

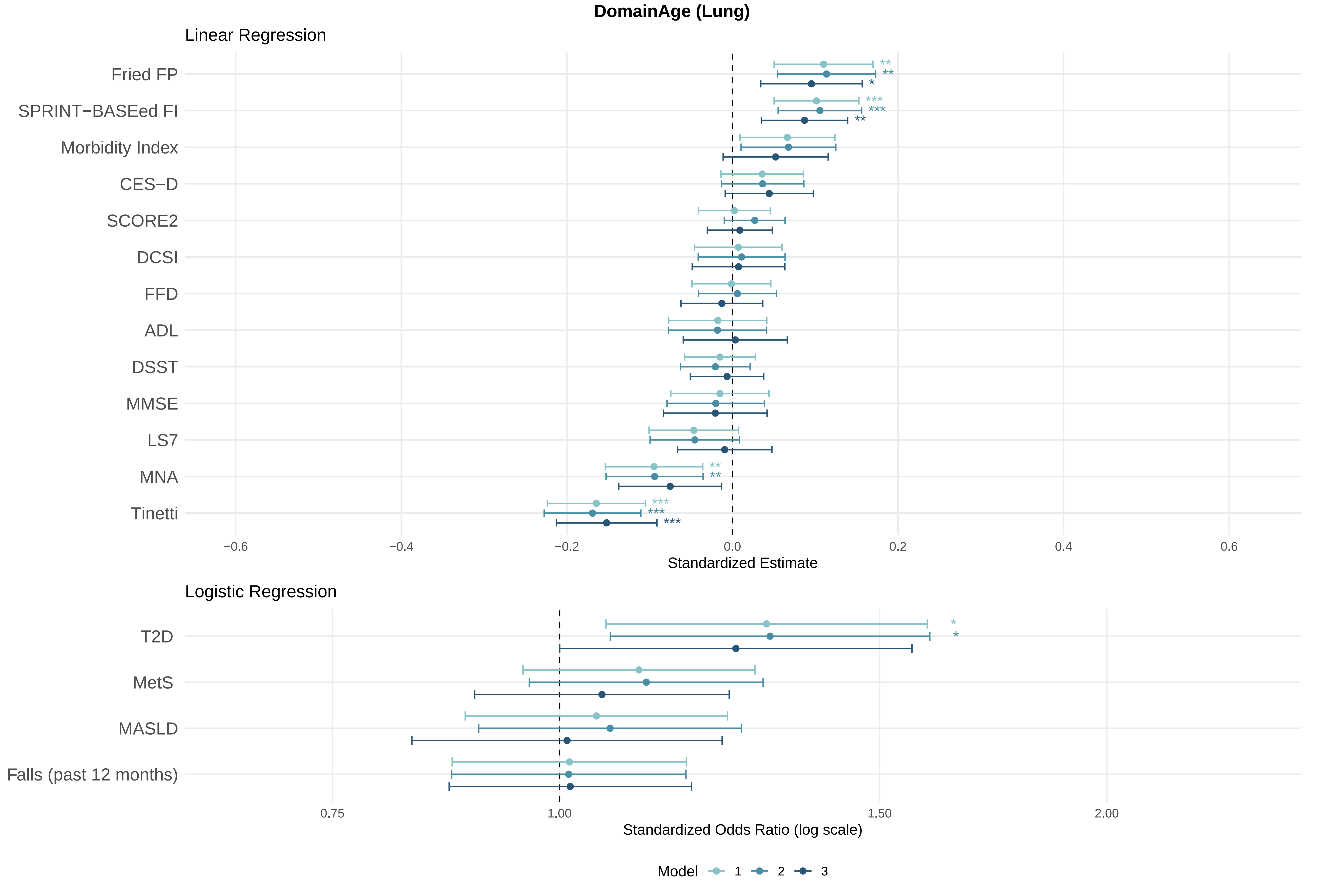

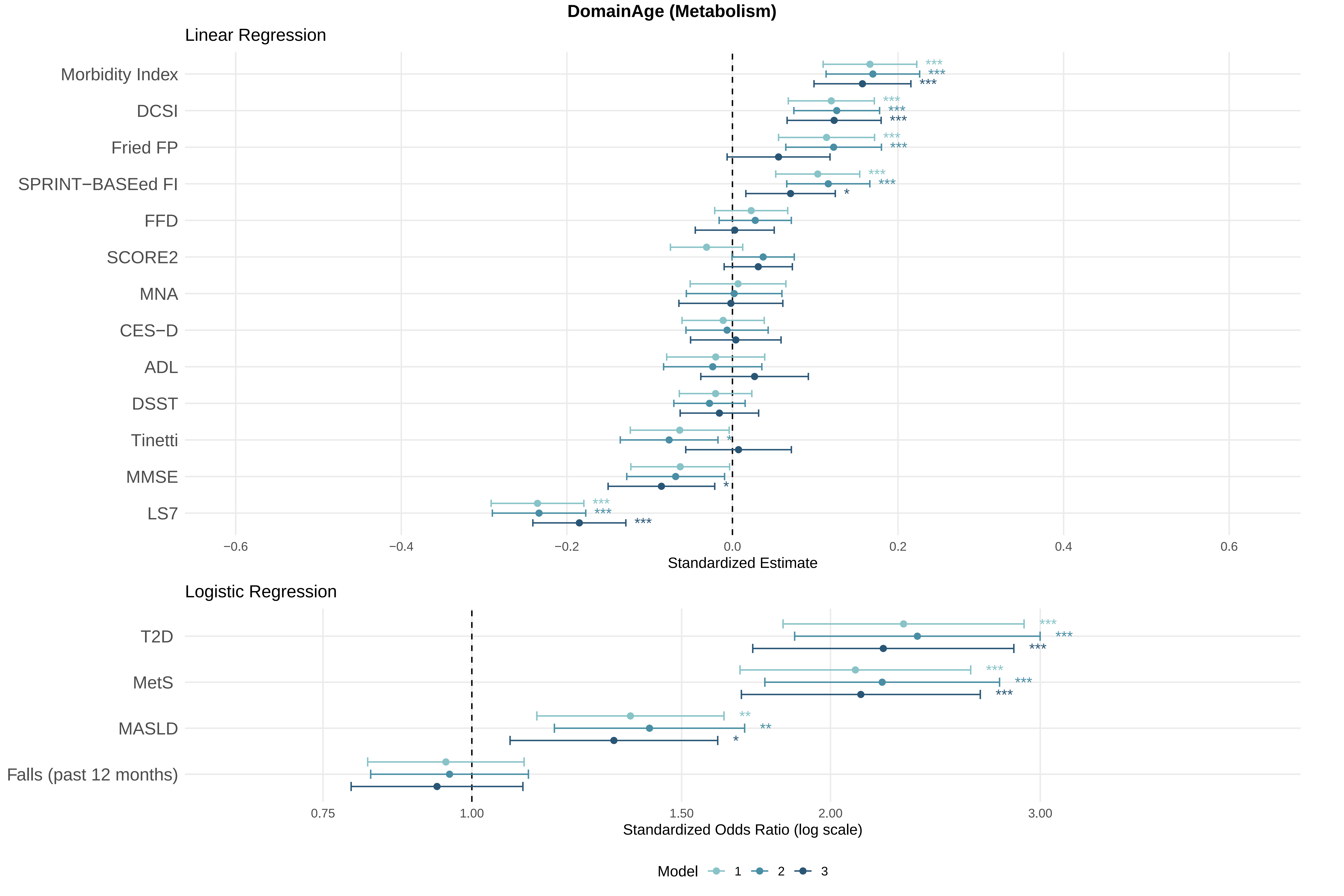

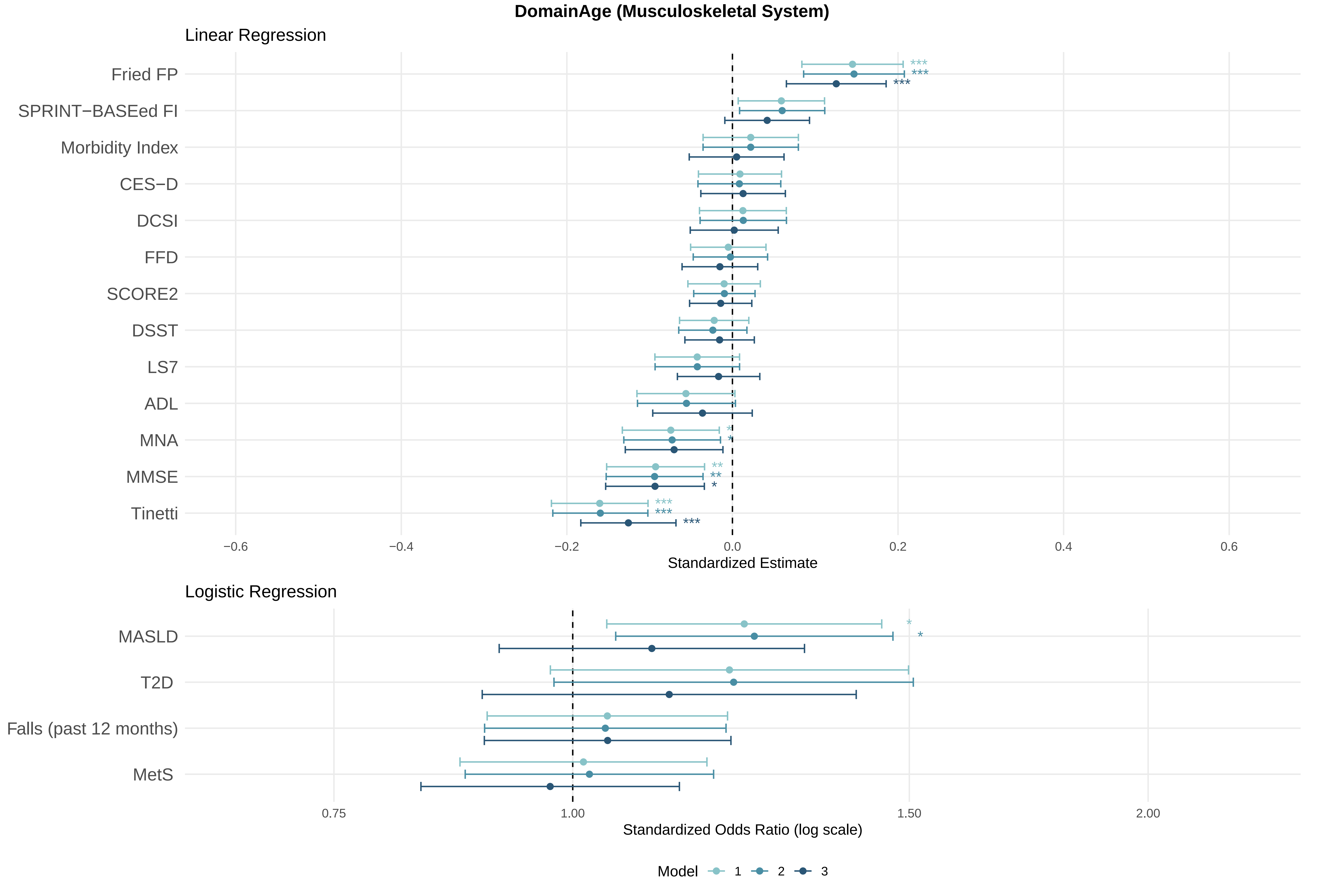

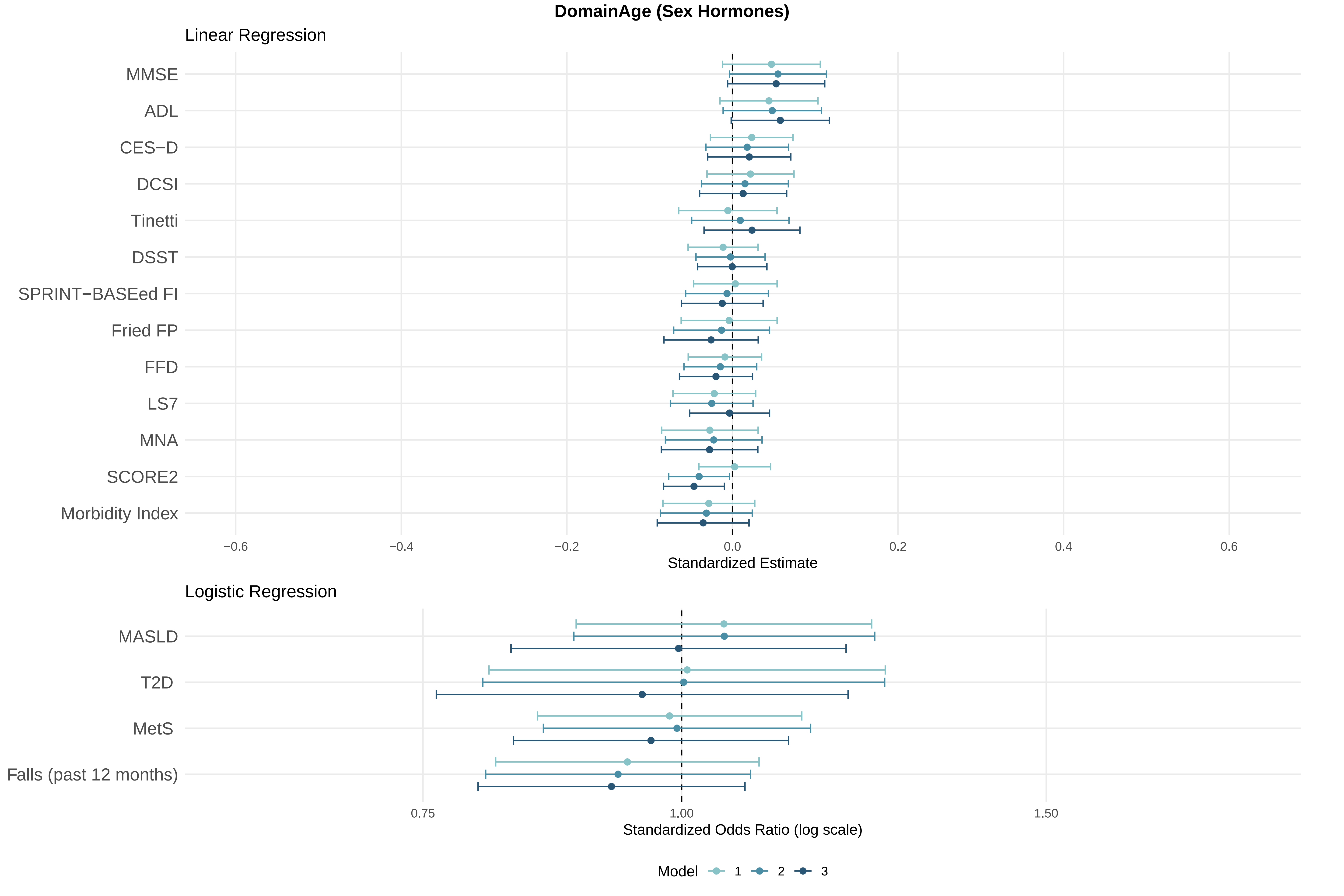

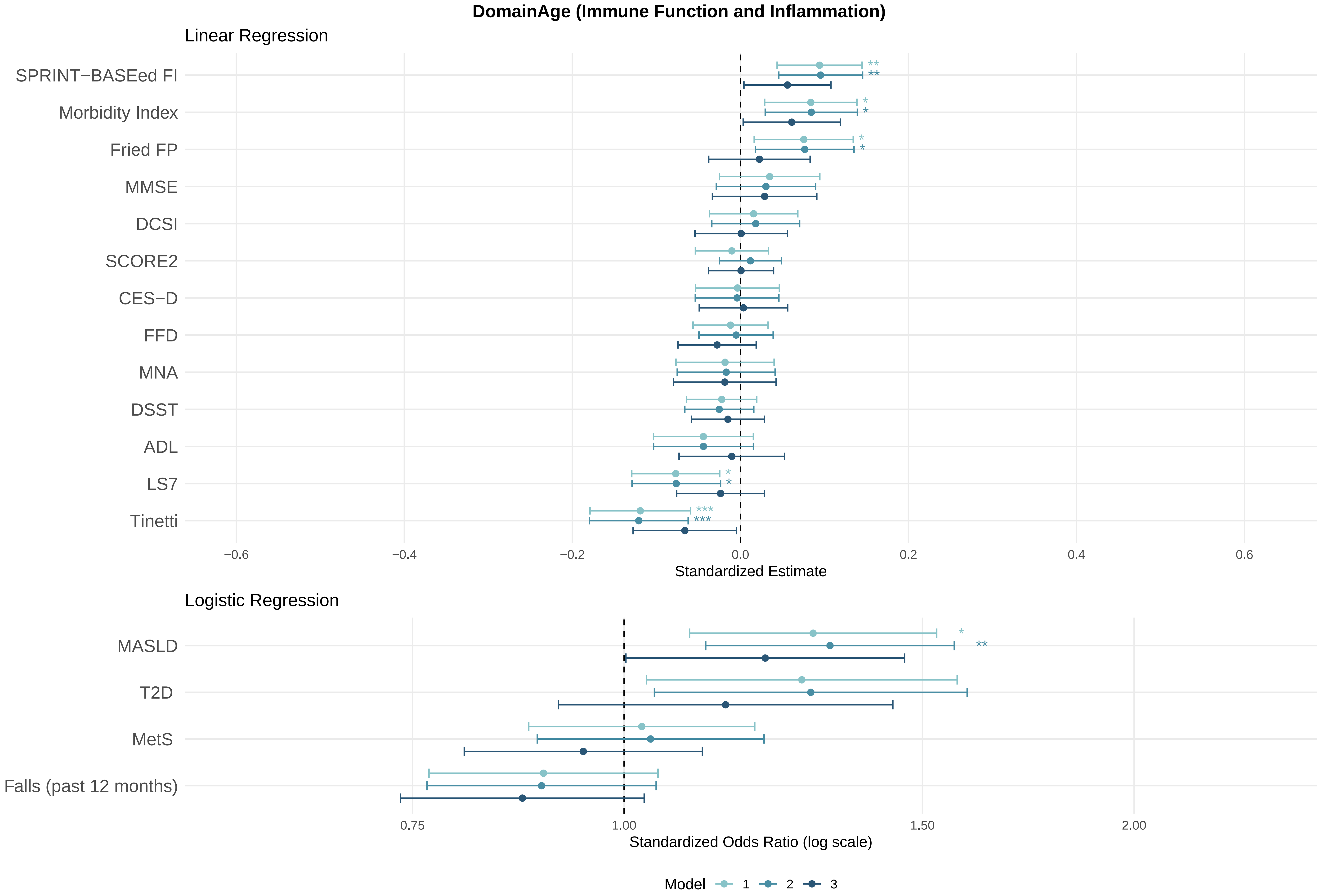

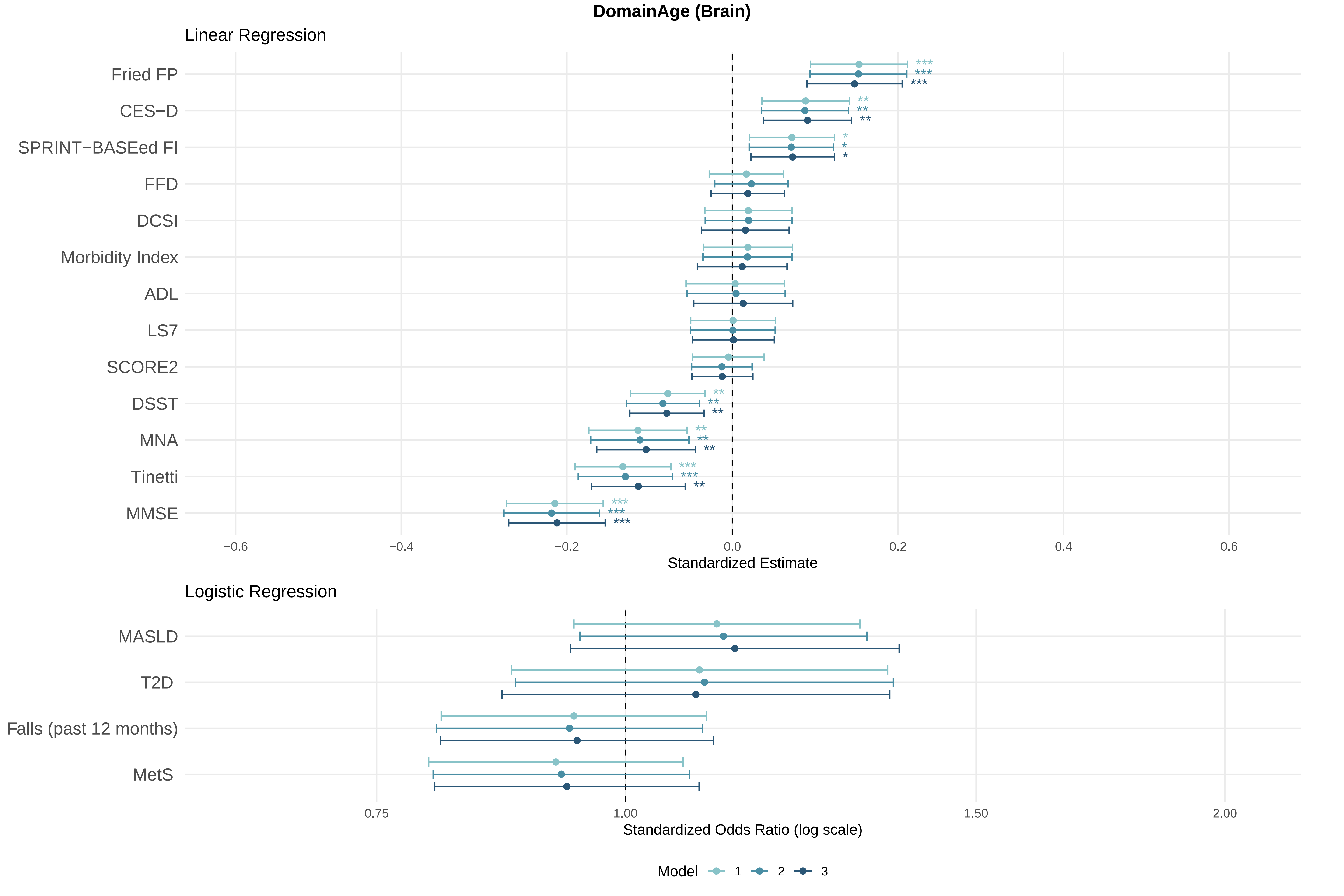

### Supplementary Figure 7

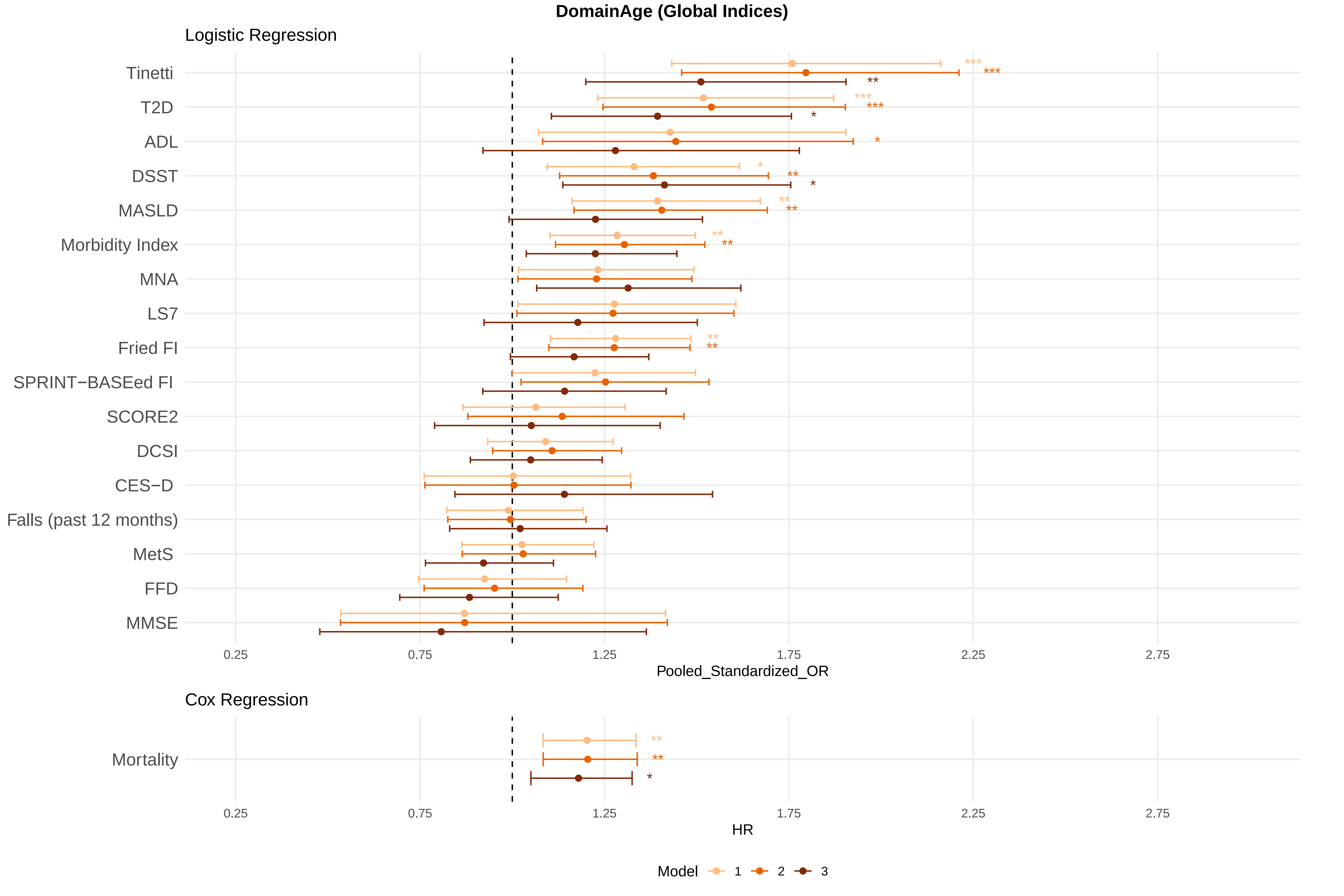

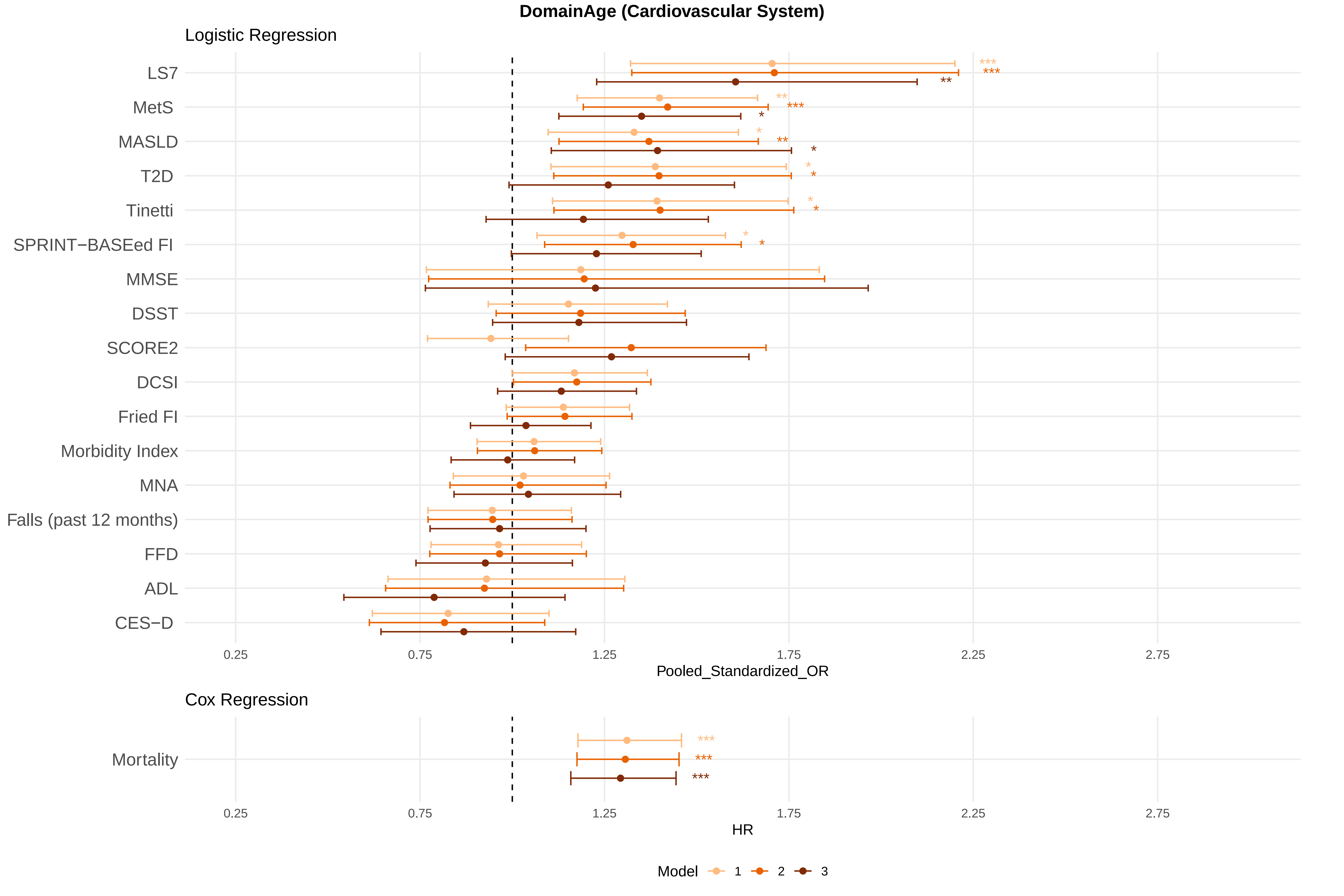

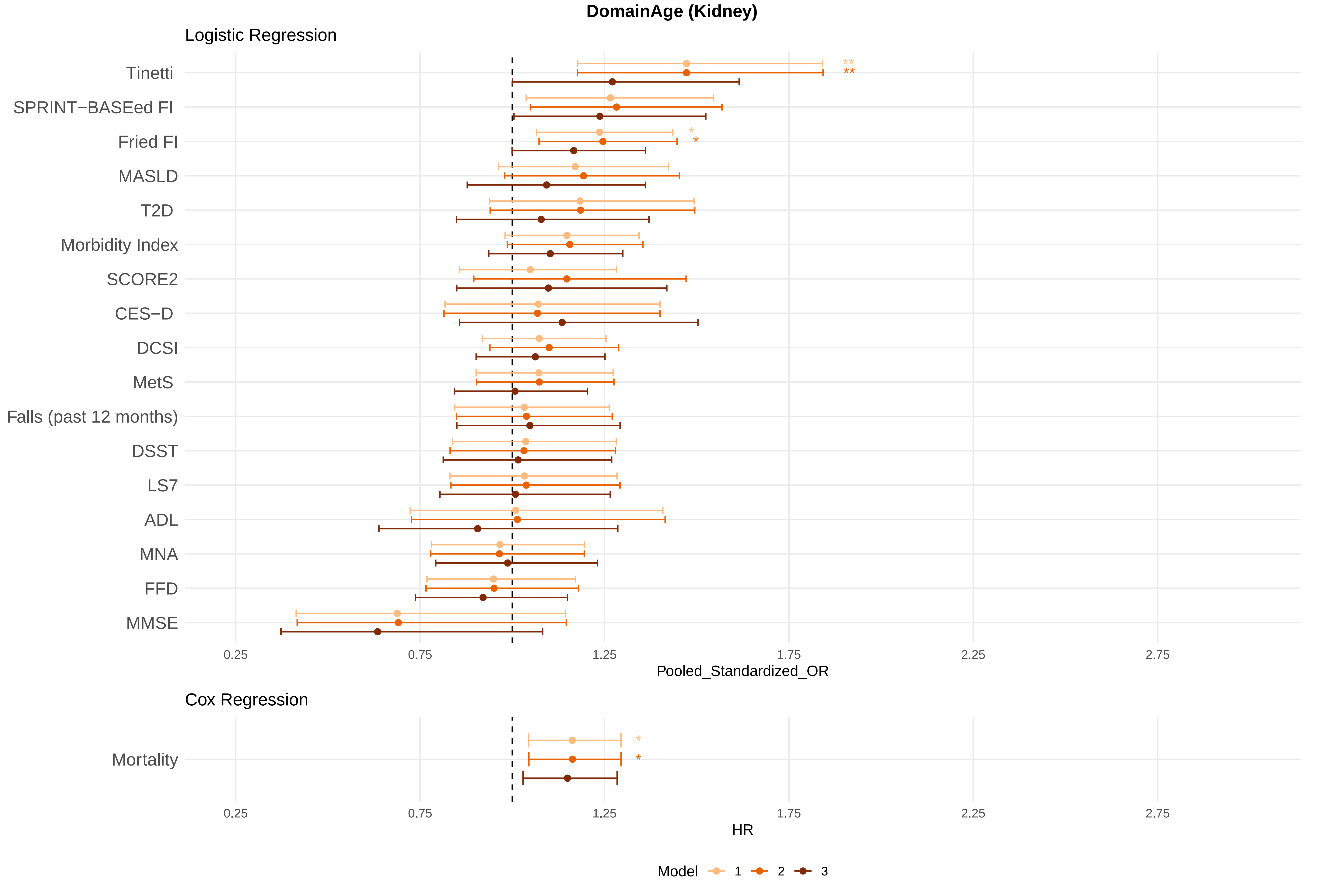

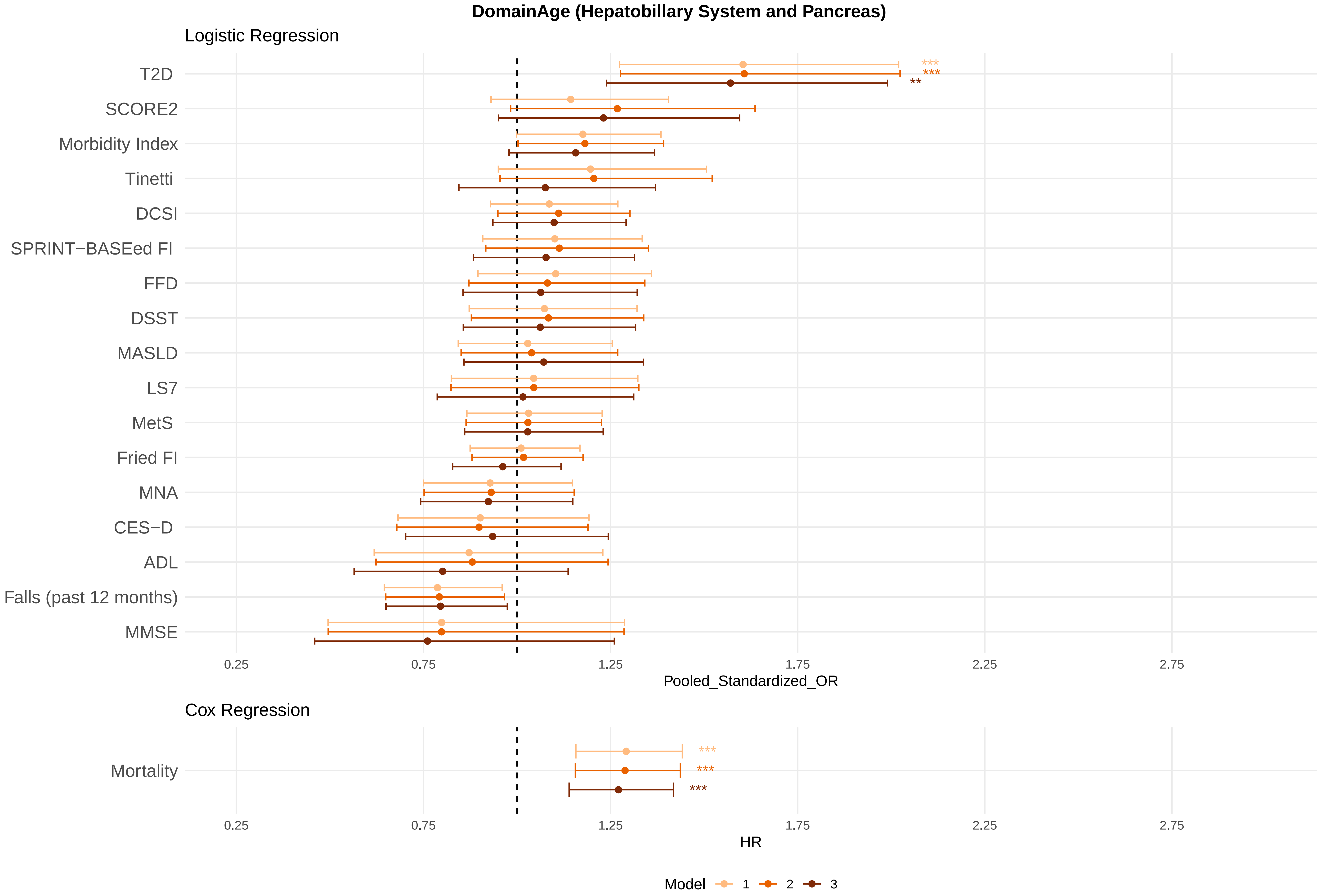

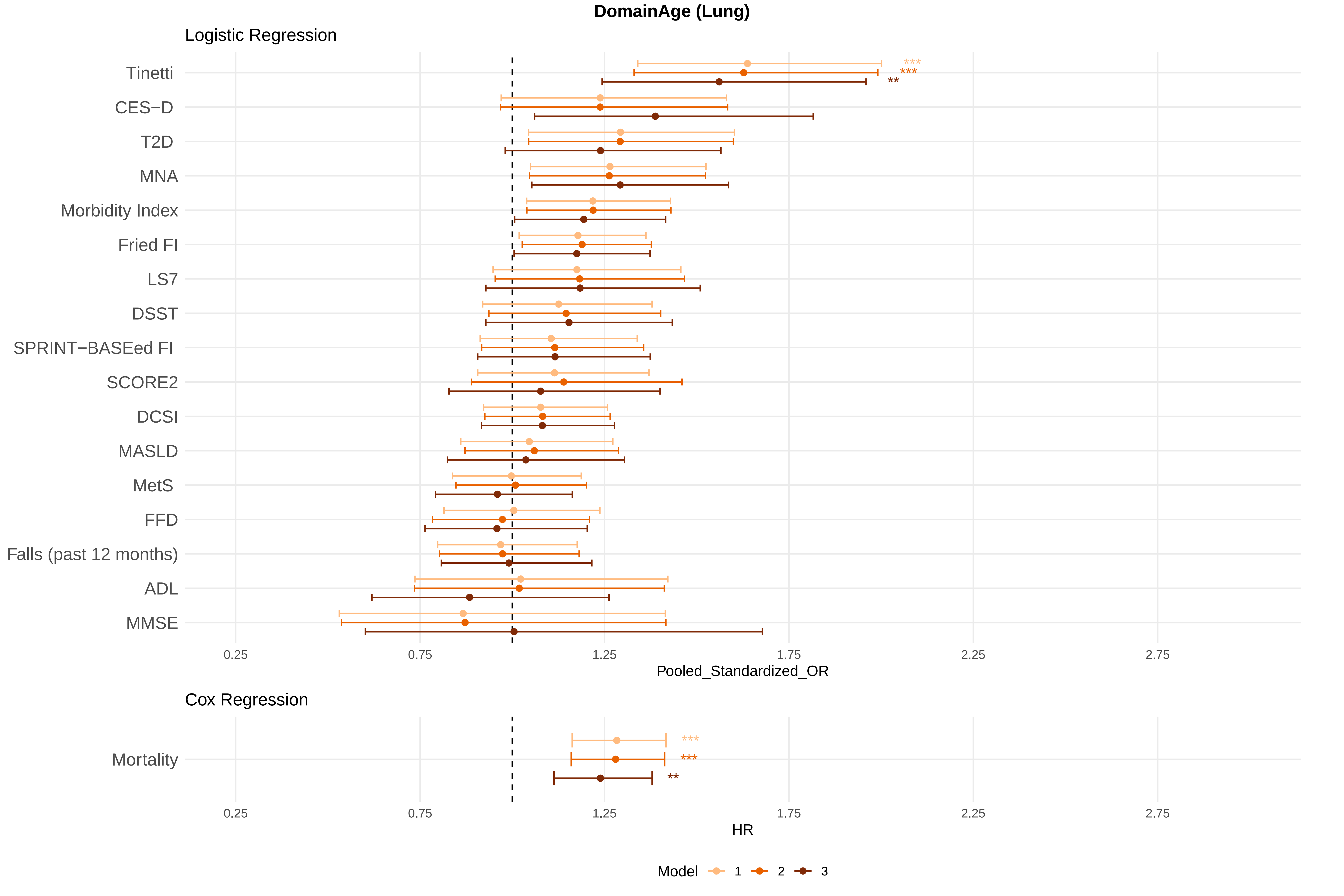

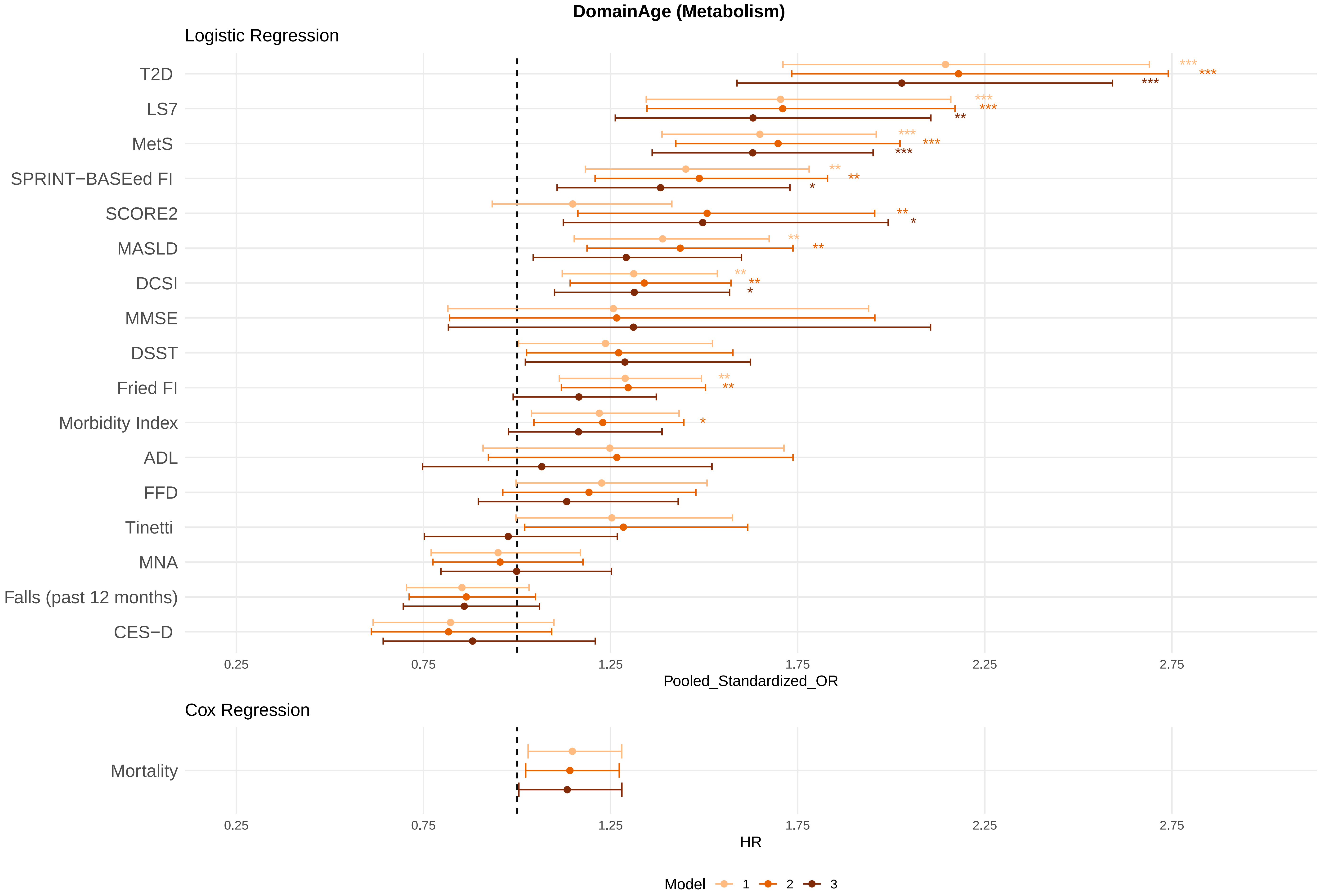

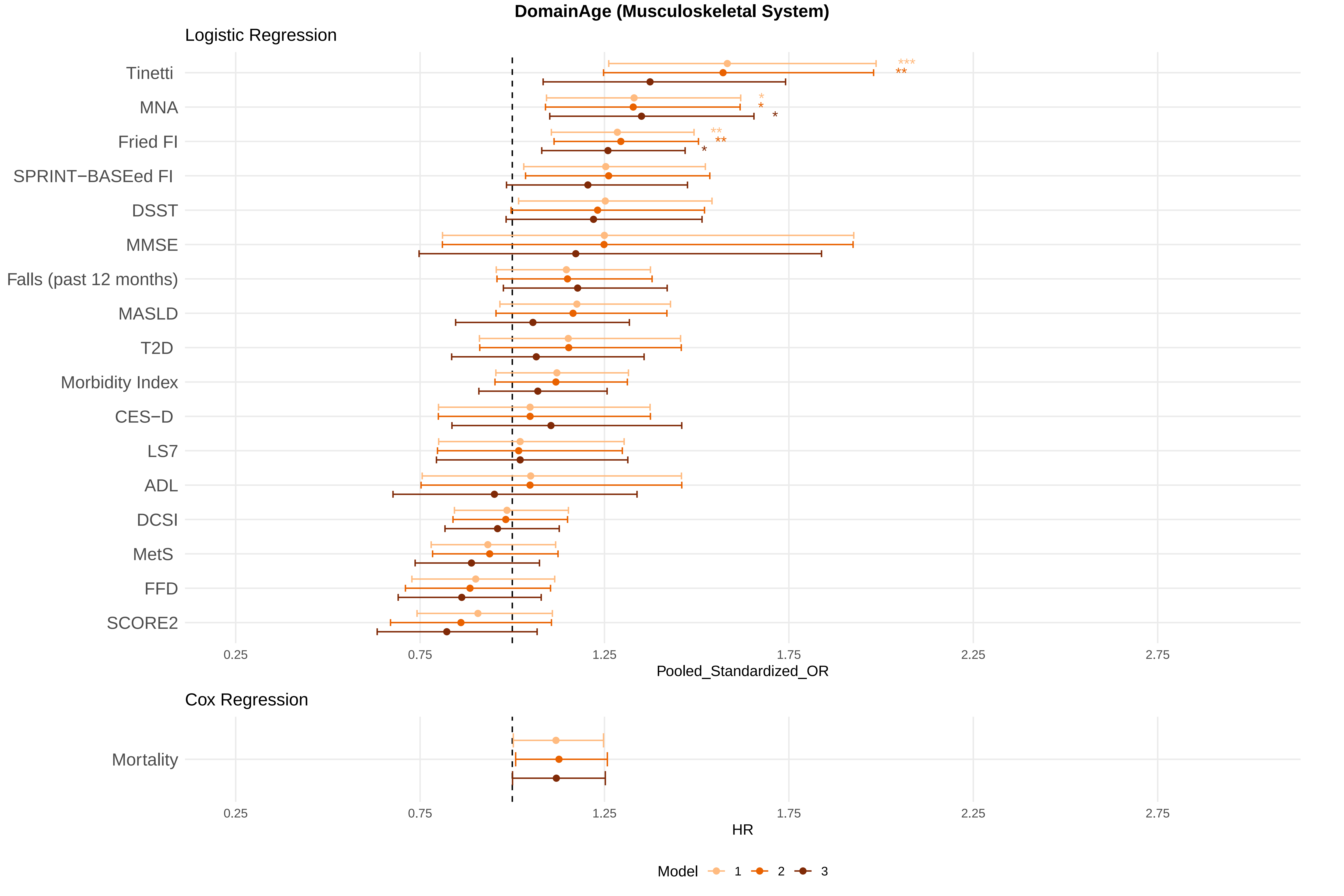

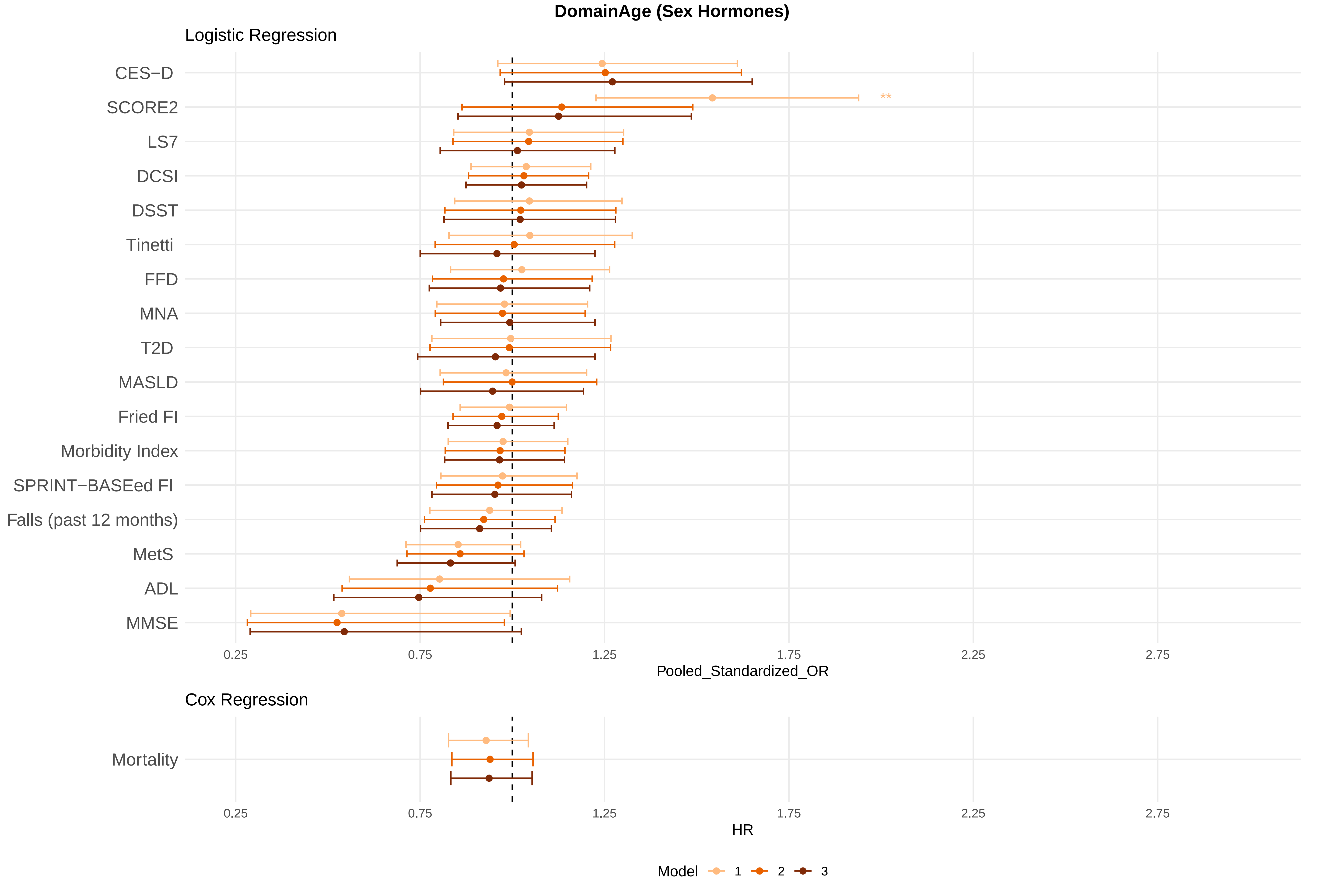

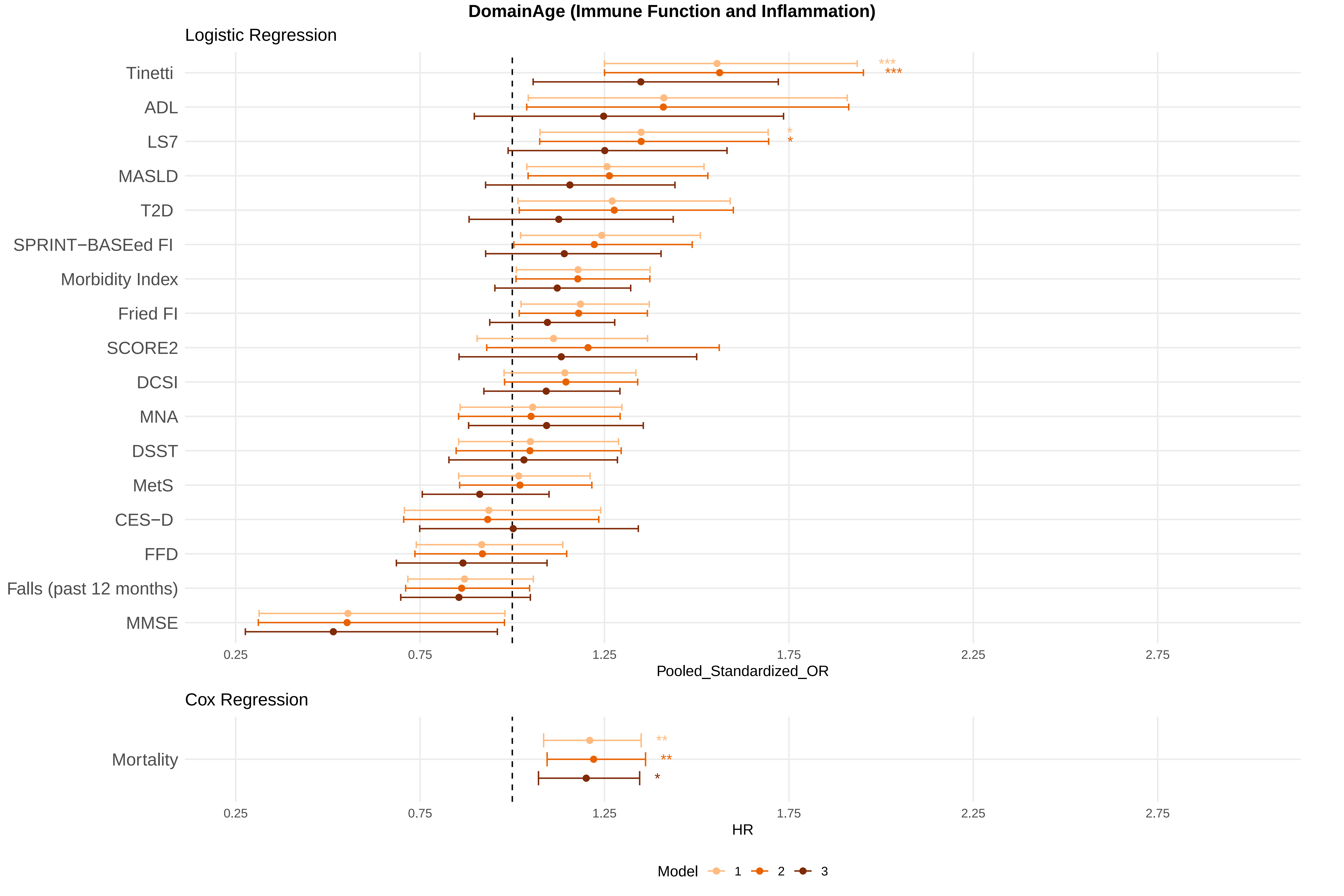

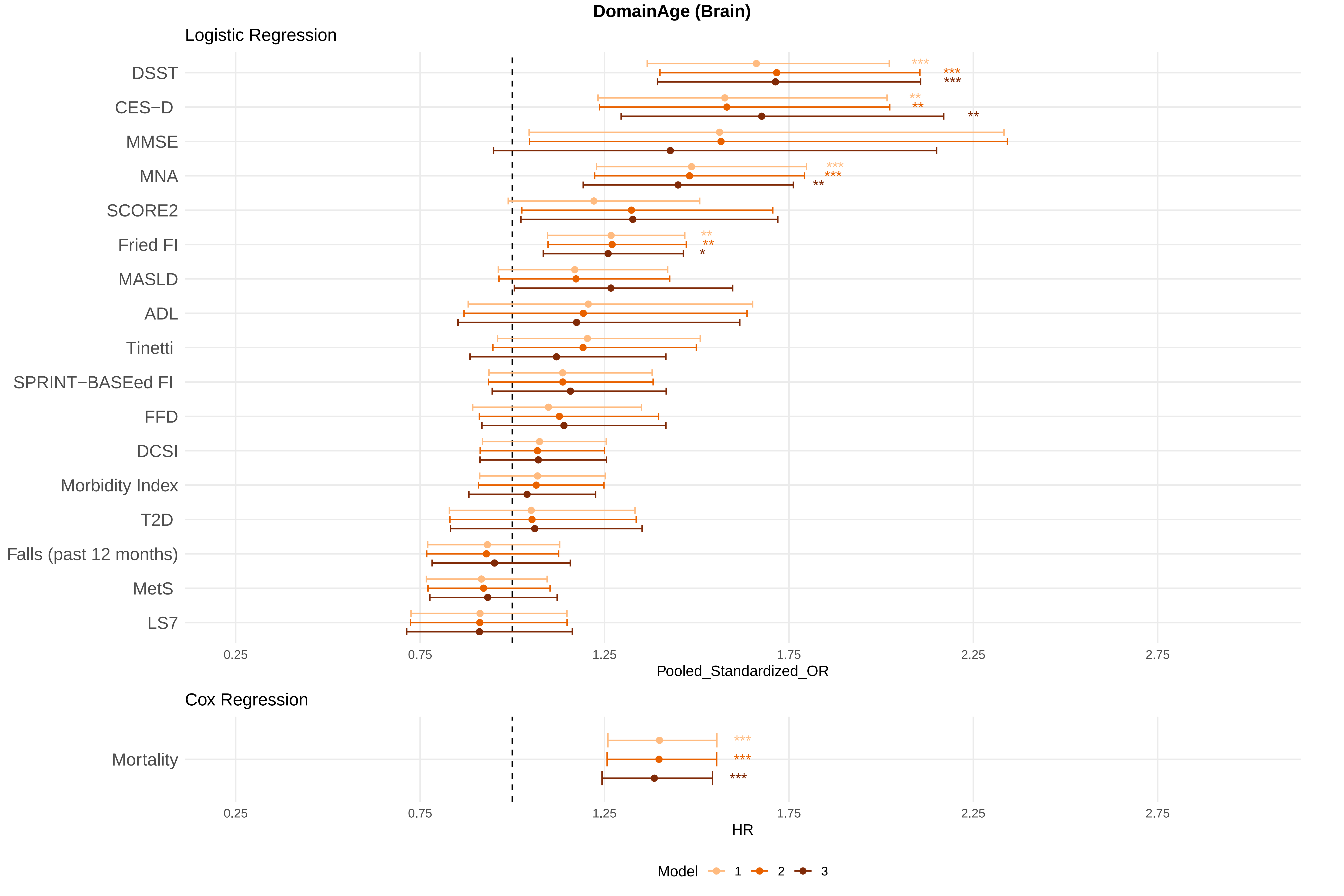
