## Supplementary Figure 2 for "MultiAge: A New Multidimensional Biomarker of Biological Age Derived from Comprehensive Phenotypic and Molecular Profiling"

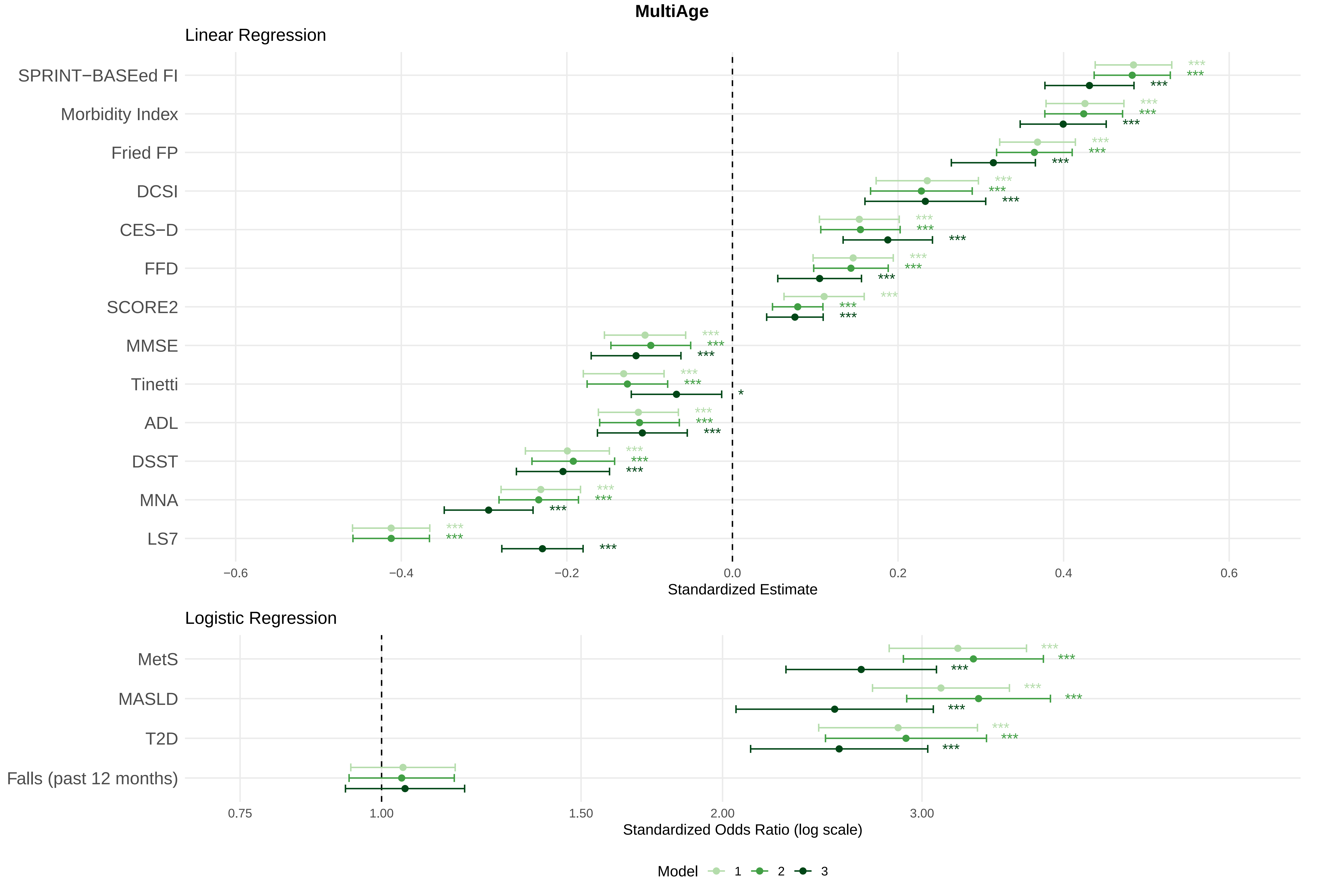
MultiAge
Linear Regression
SPRINT−BASEed FI
Morbidity Index
Fried FP
DCSI
CES−D
FFD
SCORE2
MMSE
Tinetti
ADL
DSST
MNA
LS7
−0.6
−0.4
−0.2
0.0
0.2
0.4
0.6
Standardized Estimate
Logistic Regression
MetS
MASLD
T2D
Falls (past 12 months)
0.75
1.00
1.50
2.00
3.00
Standardized Odds Ratio (log scale)
Model
1
2
3

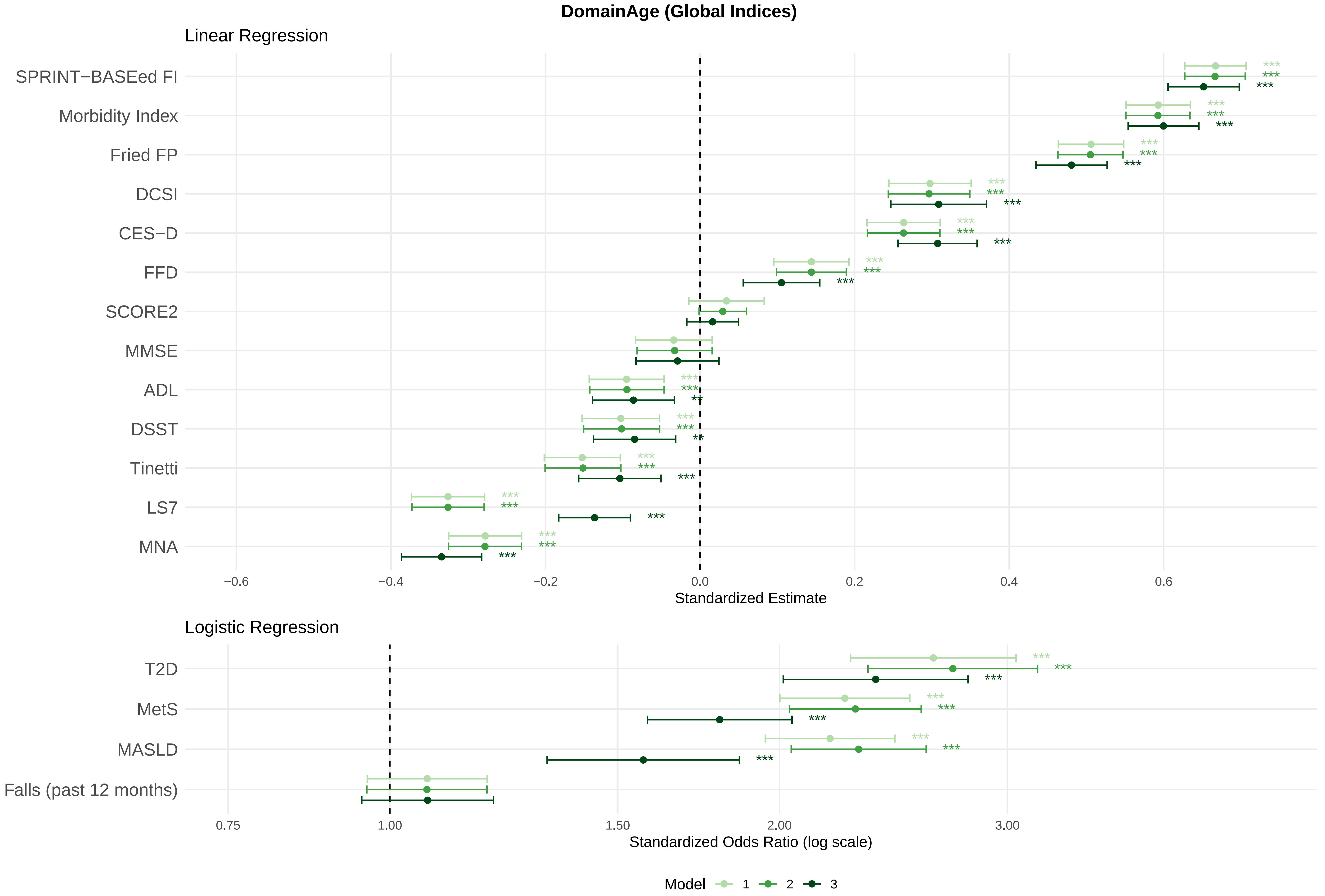

DomainAge (Global Indices)
Linear Regression
SPRINT−BASEed FI
Morbidity Index
Fried FP
DCSI
CES−D
FFD
SCORE2
MMSE
ADL
DSST
Tinetti
LS7
MNA
−0.6
−0.4
−0.2
0.0
0.2
0.4
0.6
Standardized Estimate
Logistic Regression
T2D
MetS
MASLD
Falls (past 12 months)
0.75
1.00
1.50
2.00
3.00
Standardized Odds Ratio (log scale)
Model 1 2 3

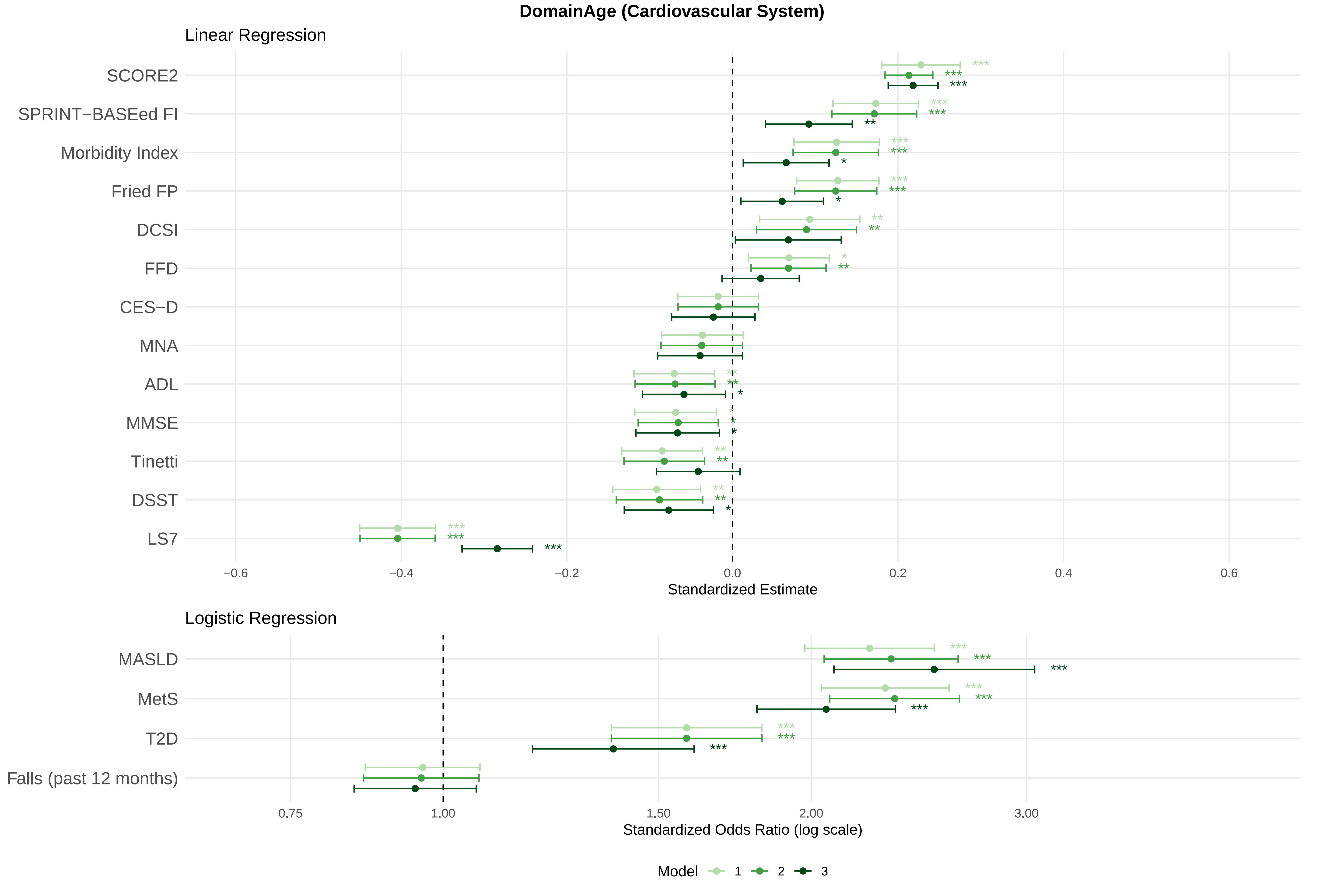

DomainAge (Cardiovascular System)
Linear Regression
SCORE2
SPRINT−BASEed FI
Morbidity Index
Fried FP
DCSI
FFD
CES−D
MNA
ADL
MMSE
Tinetti
DSST
LS7
−0.6
−0.4
−0.2
0.0
0.2
0.4
0.6
Standardized Estimate
Logistic Regression
MASLD
MetS
T2D
Falls (past 12 months)
0.75
1.00
1.50
2.00
3.00
Standardized Odds Ratio (log scale)
Model
1
2
3

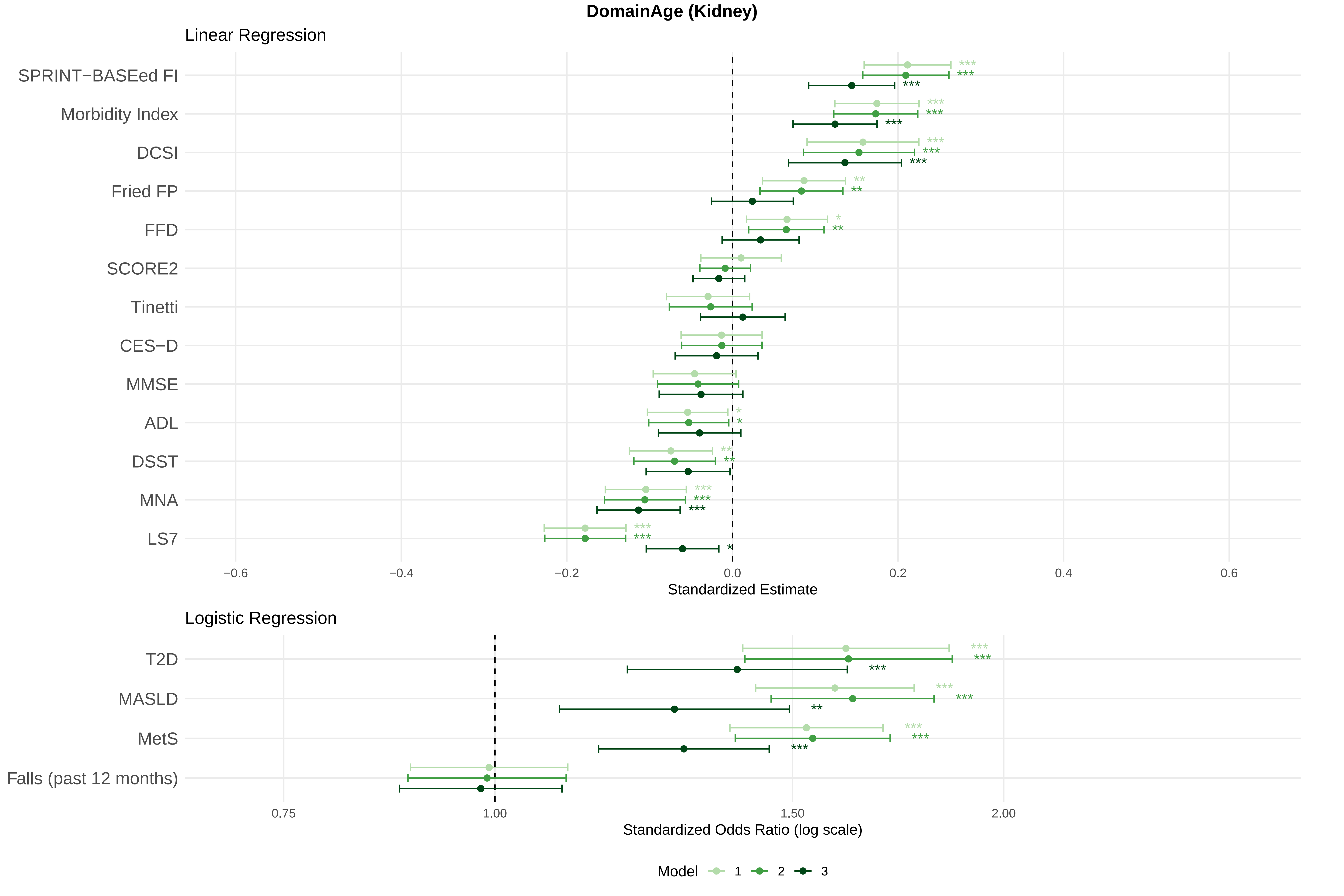
DomainAge (Kidney)
Linear Regression
SPRINT−BASEed FI
Morbidity Index
DCSI
Fried FP
FFD
SCORE2
Tinetti
CES−D
MMSE
ADL
DSST
MNA
LS7
−0.6
−0.4
−0.2
0.0
0.2
0.4
0.6
Standardized Estimate
Logistic Regression
T2D
MASLD
MetS
Falls (past 12 months)
0.75
1.00
1.50
2.00
Standardized Odds Ratio (log scale)
Model
1
2
3

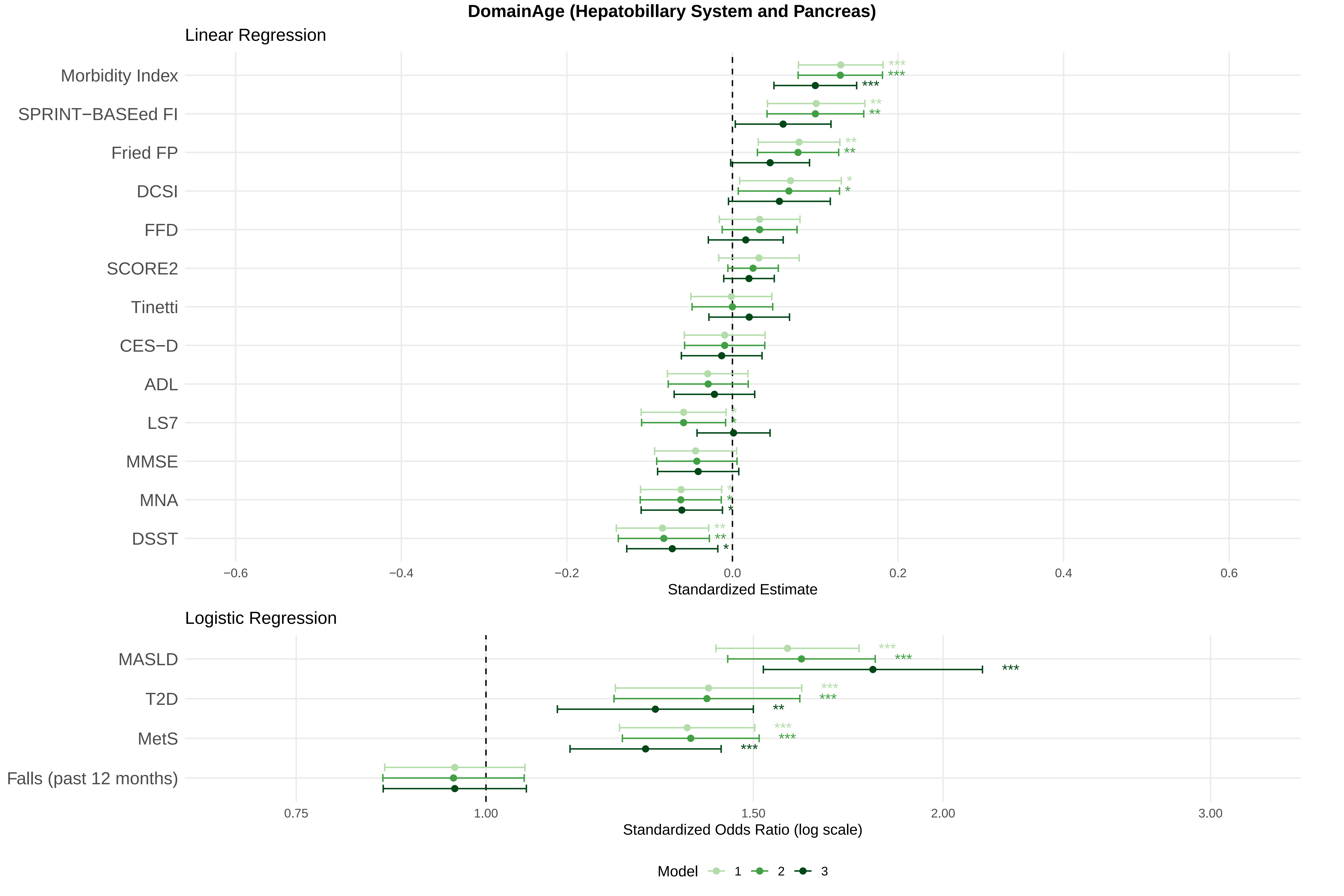

DomainAge (Hepatobillary System and Pancreas)
Linear Regression
Morbidity Index
SPRINT−BASEed FI
Fried FP
DCSI
FFD
SCORE2
Tinetti
CES−D
ADL
LS7
MMSE
MNA
DSST
−0.6
−0.4
−0.2
0.0
0.2
0.4
0.6
Standardized Estimate
Logistic Regression
MASLD
T2D
MetS
Falls (past 12 months)
0.75
1.00
1.50
2.00
3.00
Standardized Odds Ratio (log scale)
Model
1
2
3

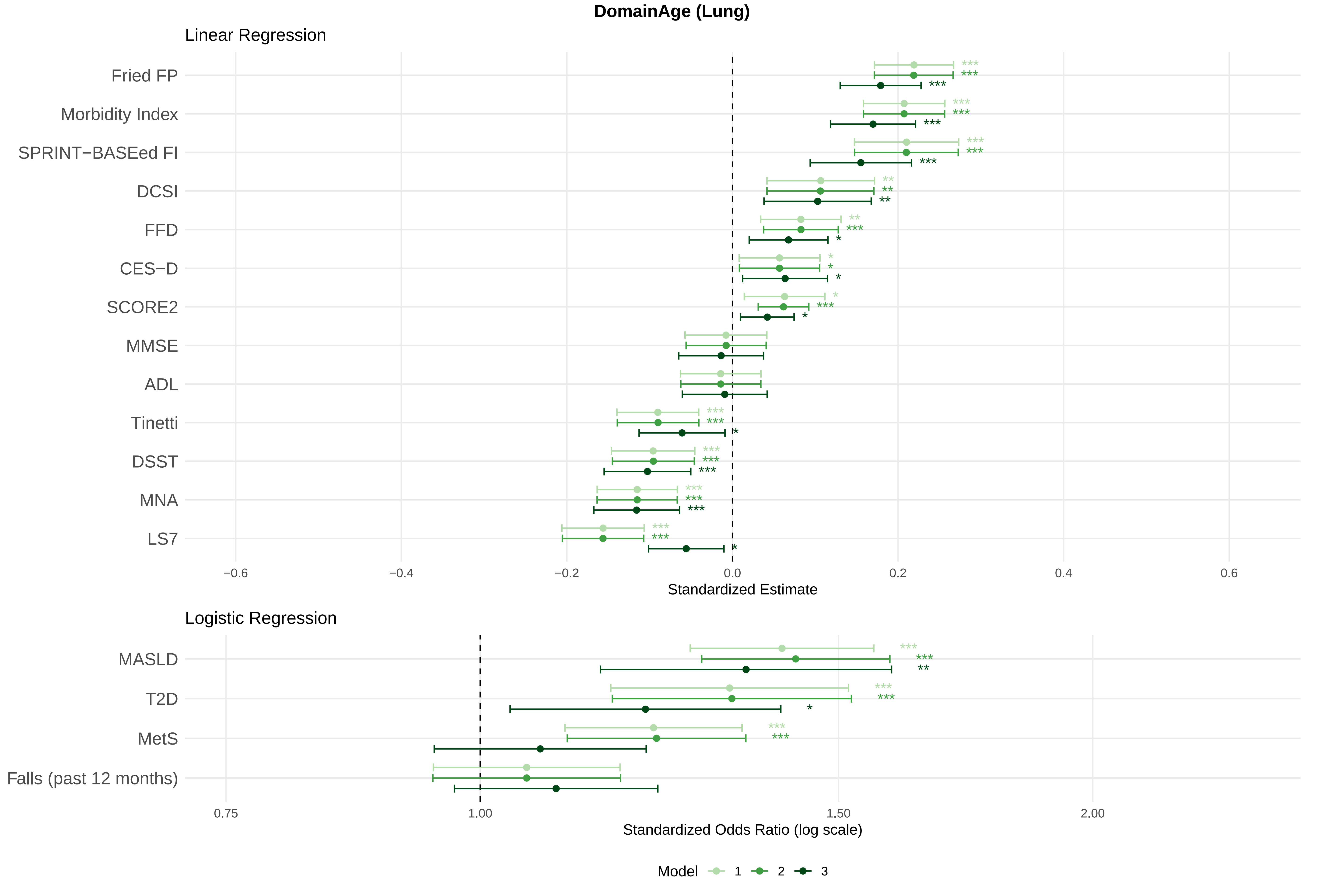
DomainAge (Lung)
Linear Regression
Fried FP
Morbidity Index
SPRINT−BASEed FI
DCSI
FFD
CES−D
SCORE2
MMSE
ADL
Tinetti
DSST
MNA
LS7
−0.6
−0.4
−0.2
0.0
0.2
0.4
0.6
Standardized Estimate
Logistic Regression
MASLD
T2D
MetS
Falls (past 12 months)
0.75
1.00
1.50
2.00
Standardized Odds Ratio (log scale)
Model 1 2 3

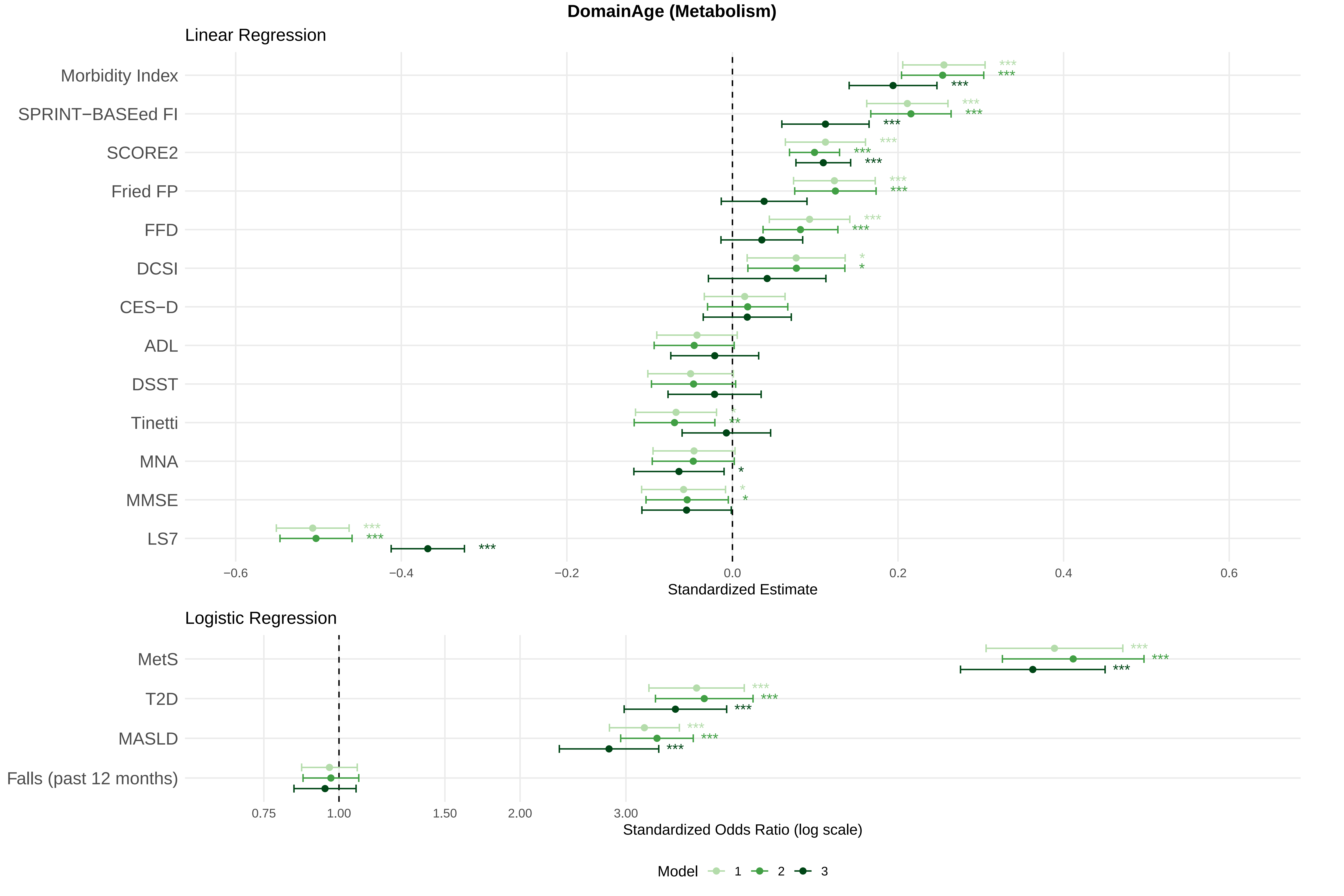

DomainAge (Metabolism)
Linear Regression
Morbidity Index
SPRINT−BASEed FI
SCORE2
Fried FP
FFD
DCSI
CES−D
ADL
DSST
Tinetti
MNA
MMSE
LS7
−0.6
−0.4
−0.2
0.0
0.2
0.4
0.6
Standardized Estimate
Logistic Regression
MetS
T2D
MASLD
Falls (past 12 months)
0.75
1.00
1.50
2.00
3.00
Standardized Odds Ratio (log scale)
Model
1
2
3

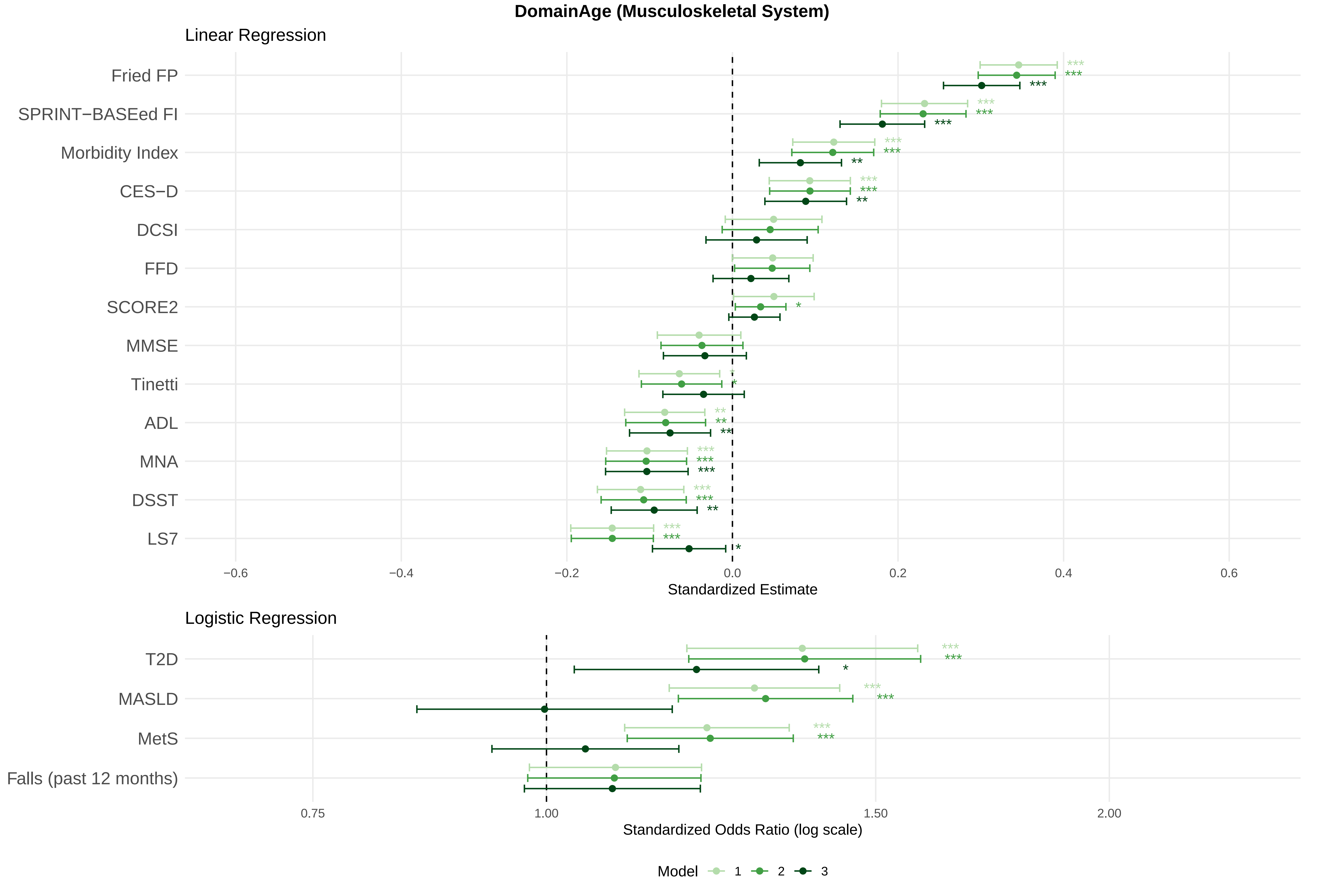
DomainAge (Musculoskeletal System)
Linear Regression
Fried FP
SPRINT−BASEed FI
Morbidity Index
CES−D
DCSI
FFD
SCORE2
MMSE
Tinetti
ADL
MNA
DSST
LS7
−0.6
−0.4
−0.2
0.0
0.2
0.4
0.6
Standardized Estimate
Logistic Regression
T2D
MASLD
MetS
Falls (past 12 months)
0.75
1.00
1.50
2.00
Standardized Odds Ratio (log scale)
Model
1
2
3

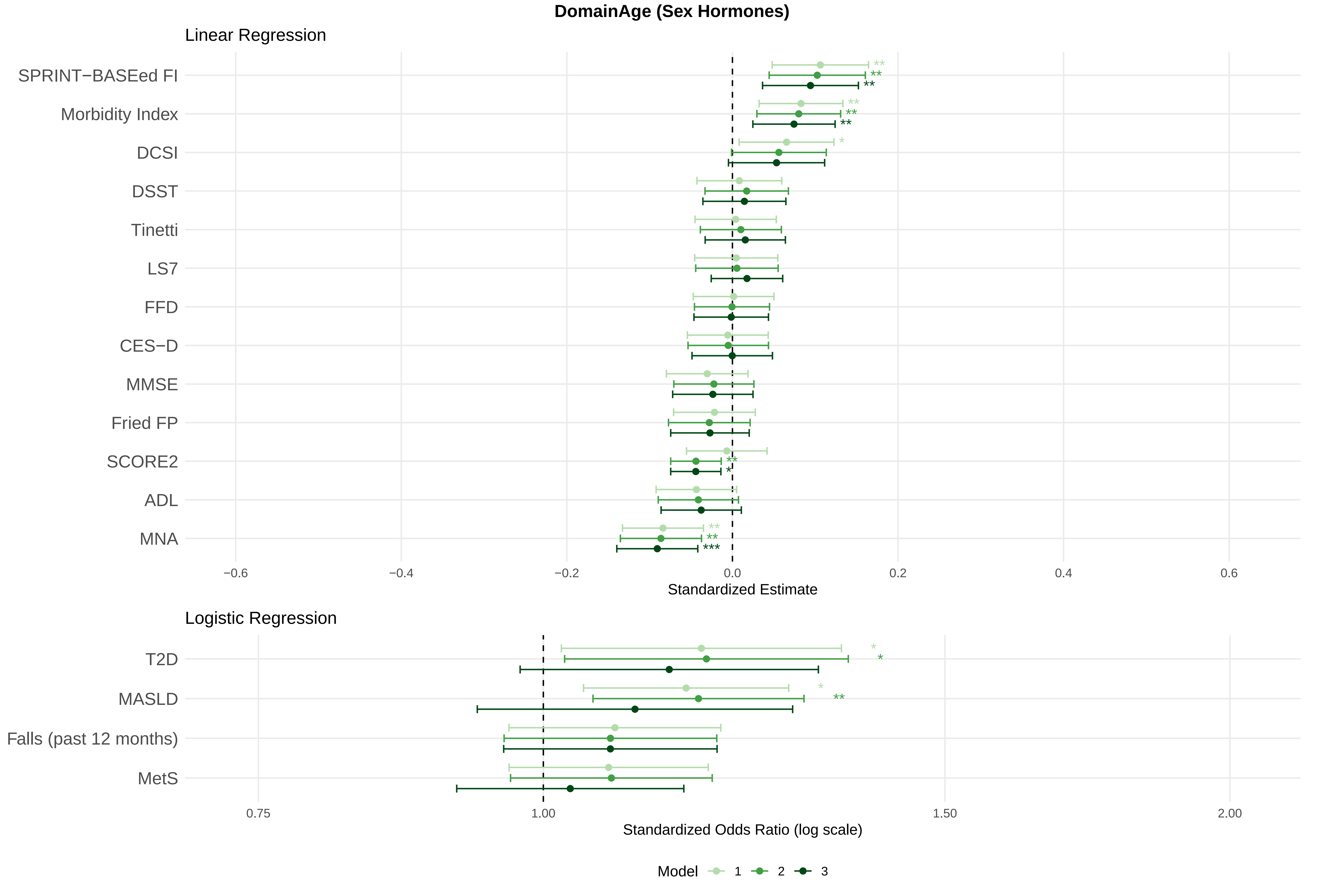
DomainAge (Sex Hormones)
Linear Regression
SPRINT−BASEed FI
Morbidity Index
DCSI
DSST
Tinetti
LS7
FFD
CES−D
MMSE
Fried FP
SCORE2
ADL
MNA
−0.6
−0.4
−0.2
0.0
0.2
0.4
0.6
Standardized Estimate
Logistic Regression
T2D
MASLD
Falls (past 12 months)
MetS
0.75
1.00
1.50
2.00
Standardized Odds Ratio (log scale)
Model 1 2 3

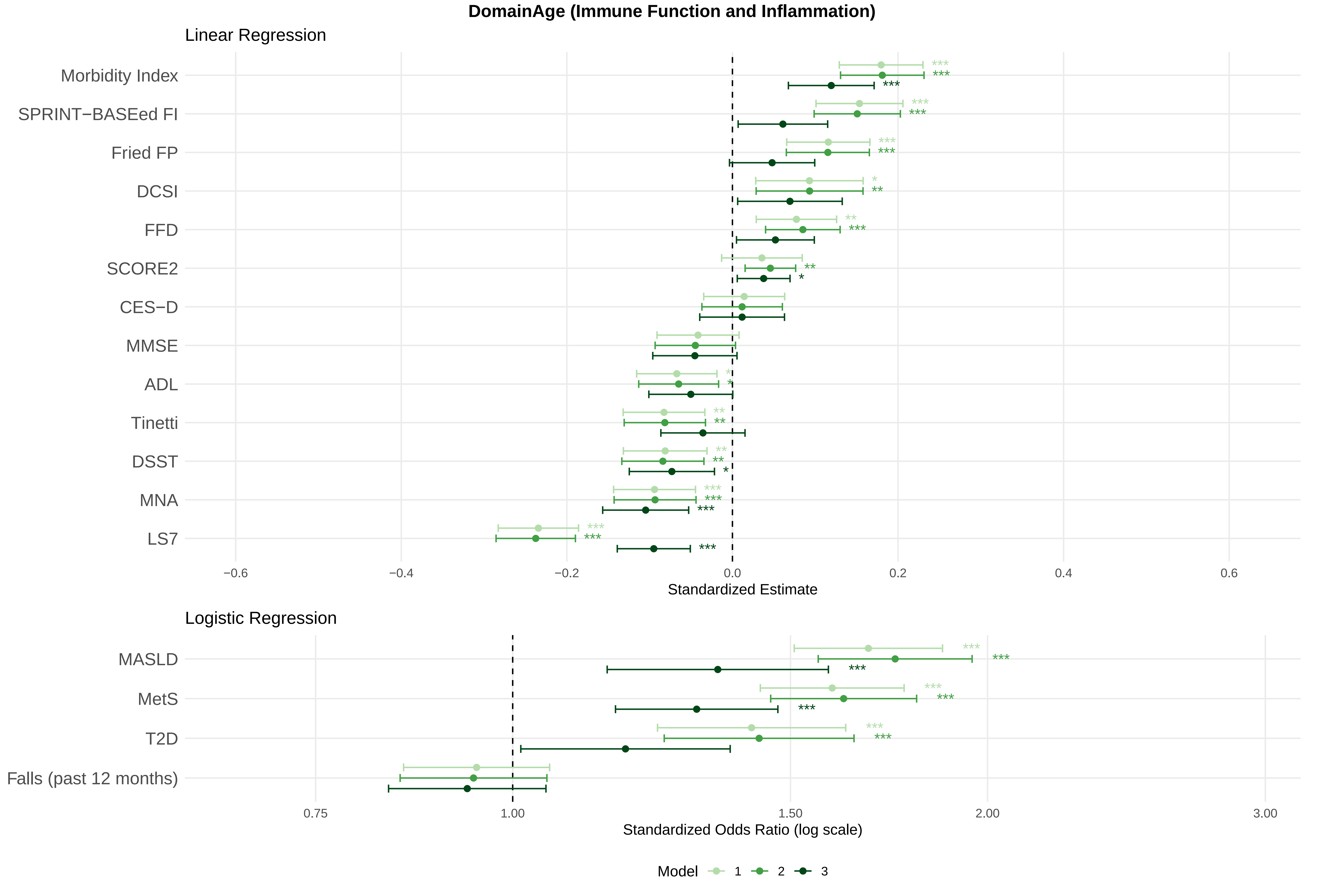

DomainAge (Immune Function and Inflammation)
Linear Regression
Morbidity Index
SPRINT−BASEed FI
Fried FP
DCSI
FFD
SCORE2
CES−D
MMSE
ADL
Tinetti
DSST
MNA
LS7
−0.6
−0.4
−0.2
0.0
0.2
0.4
0.6
Standardized Estimate
Logistic Regression
MASLD
MetS
T2D
Falls (past 12 months)
0.75
1.00
1.50
2.00
3.00
Standardized Odds Ratio (log scale)
Model
1
2
3

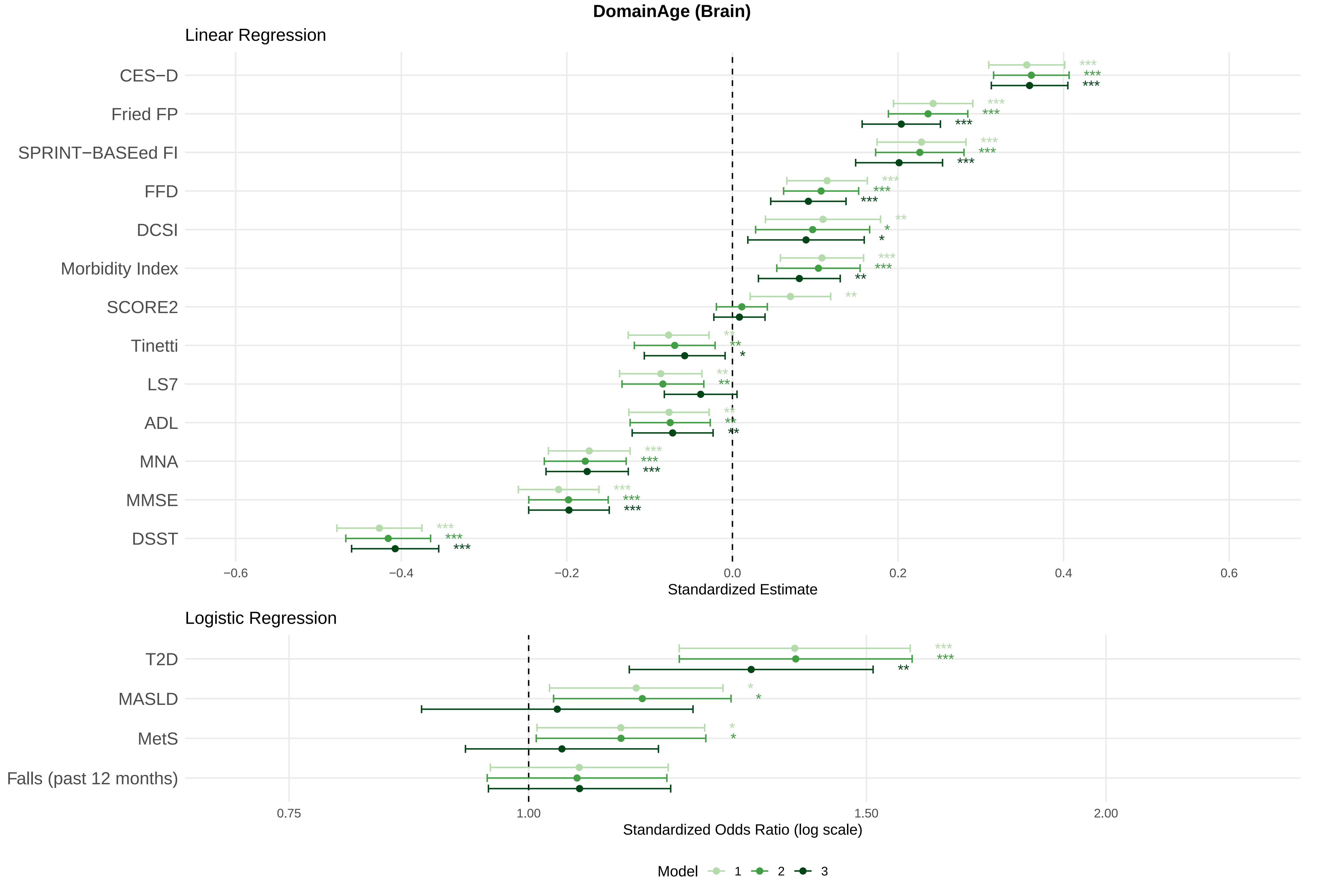

DomainAge (Brain)
Linear Regression
CES−D
Fried FP
SPRINT−BASEed FI
FFD
DCSI
Morbidity Index
SCORE2
Tinetti
LS7
ADL
MNA
MMSE
DSST
−0.6
−0.4
−0.2
0.0
0.2
0.4
0.6
Standardized Estimate
Logistic Regression
T2D
MASLD
MetS
Falls (past 12 months)
0.75
1.00
1.50
2.00
Standardized Odds Ratio (log scale)
Model
1
2
3
