## Supplementary Figure 4 for "MultiAge: A New Multidimensional Biomarker of Biological Age Derived from Comprehensive Phenotypic and Molecular Profiling"

### Slide 1
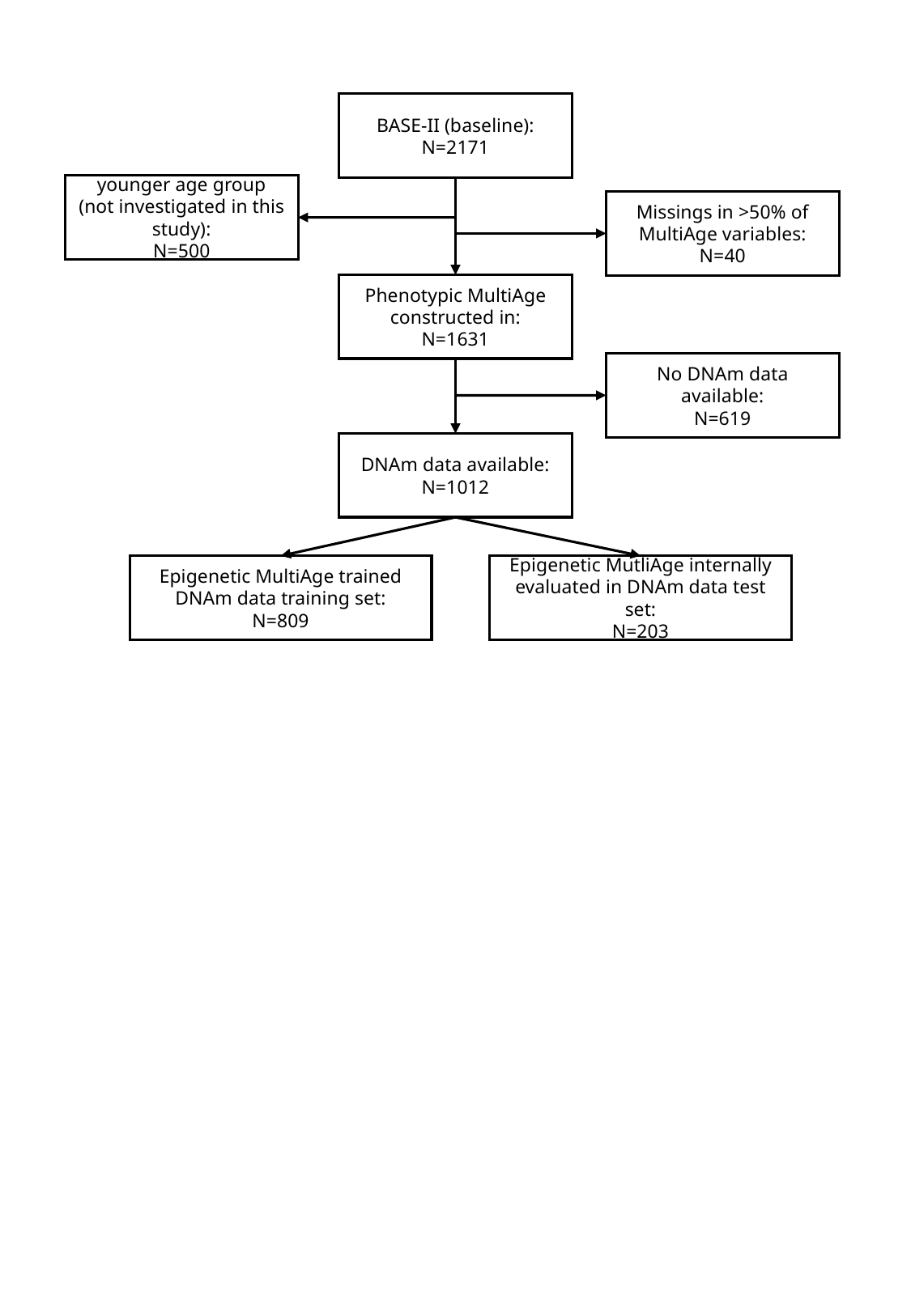

BASE-II (baseline):
N=2171
younger age group (not investigated in this study):
N=500
Missings in >50% of MultiAge variables:
N=40
Phenotypic MultiAge constructed in:
N=1631
No DNAm data available:
N=619
DNAm data available:
N=1012
Epigenetic MultiAge trained DNAm data training set:
N=809
Epigenetic MutliAge internally evaluated in DNAm data test set:
N=203
